# Counting Cases and Deaths by Age Tells Us About COVID-19’s Infectious and Lethal Components

**DOI:** 10.1101/2023.01.05.23284239

**Authors:** James Michaelson

## Abstract

Gauging COVID-19’s *lethality*, and how vaccination can reduce that *lethality*, has been challenging. Here, a new method, *Gompertzian Analysis*, counting cases and deaths, by age, and displaying them on logarithmic graphs, is outlined, and its first findings presented: FIRST, COVID-19 *Gompertzian Lethality* (Deaths/Cases) exhibits an ~10,000-fold exponential increase in the chance of death with age, the *Gompertzian Force of Mortality*, captured by the *Gompertz Mortality Equation*. SECOND, COVID-19 *Pasteurian Infectivity* (Cases/Population) occurs at similar rates across ages. THIRD, the same *Gompertzian Force of Mortality* characterizes other diseases and all-cause mortality, possibly from loss of *Mitotic Dilution* of toxic compounds due to decline in mitosis. FOURTH, resistance to COVID-19 *infectivity* and *lethality* appear to be separate processes. FIFTH, Over the past several years, *Gompertzian Lethality*, has declined, but not *Pasteurian Infectivity*. SIXTH, with each variant, *Gompertzian Lethality* has declined, but not *Pasteurian Infectivity*. SEVENTH, the unvaccinated have seen a decline in *Gompertzian Lethality*, less than the vaccinated, ascribable to infection, at the cost of lives lost. EIGHTH, different vaccines have different reductions in *Gompertzian Lethality* and *Pasteurian Infectivity*. NINTH, vaccination has reduced *Pasteurian Infectivity*, but not enough to suppress the pandemic. TENTH, vaccination has reduced *Gompertzian Lethality*, with sequential vaccination pointing linearly towards zero death after 3 or 4 boosters, without signs of waning. CONCLUSION: *Gompertzian Analysis* provides new, practical, actionable, information for understanding, and minimizing, the lethal burden of COVID-19 and other diseases.

## INTRODUCTION

### The *Gompertz Mortality Equation*

In 1825, Gompertz reported that the chance of death in adulthood increases exponentially with age^1^, and in 2020, Levin et al reported the risk of COVID-19 death (i.e., Deaths/Cases) also has such a ***Gompertzian*** appearance.^2^ This quality, the ***Gompertzian Force of Mortality***, the exponential increase in that chance of death, ***D***, with age, ***t***, appears on a log graph, a ***Gompertz Plot***, as a straight ***Gompertz Line***, the ***Gompertz Mortality Equation***, whose rate of increase with age we call the ***Gompertzian Slope, G***_***s***_, and whose location on the graph, up or down, we call the ***Gompertzian Height, G***_***H***_. ***Gompertz Plots*** of the overall chance of death, and the chance of COVID-19 death, i.e., Deaths/Cases, a quality we shall call ***Gompertzian Lethality***, can be seen in FIGURE 1, APPENDIX-FIGURES 1-3, and many other figures here and in the APPENDIX.

**FIGURE 1.**
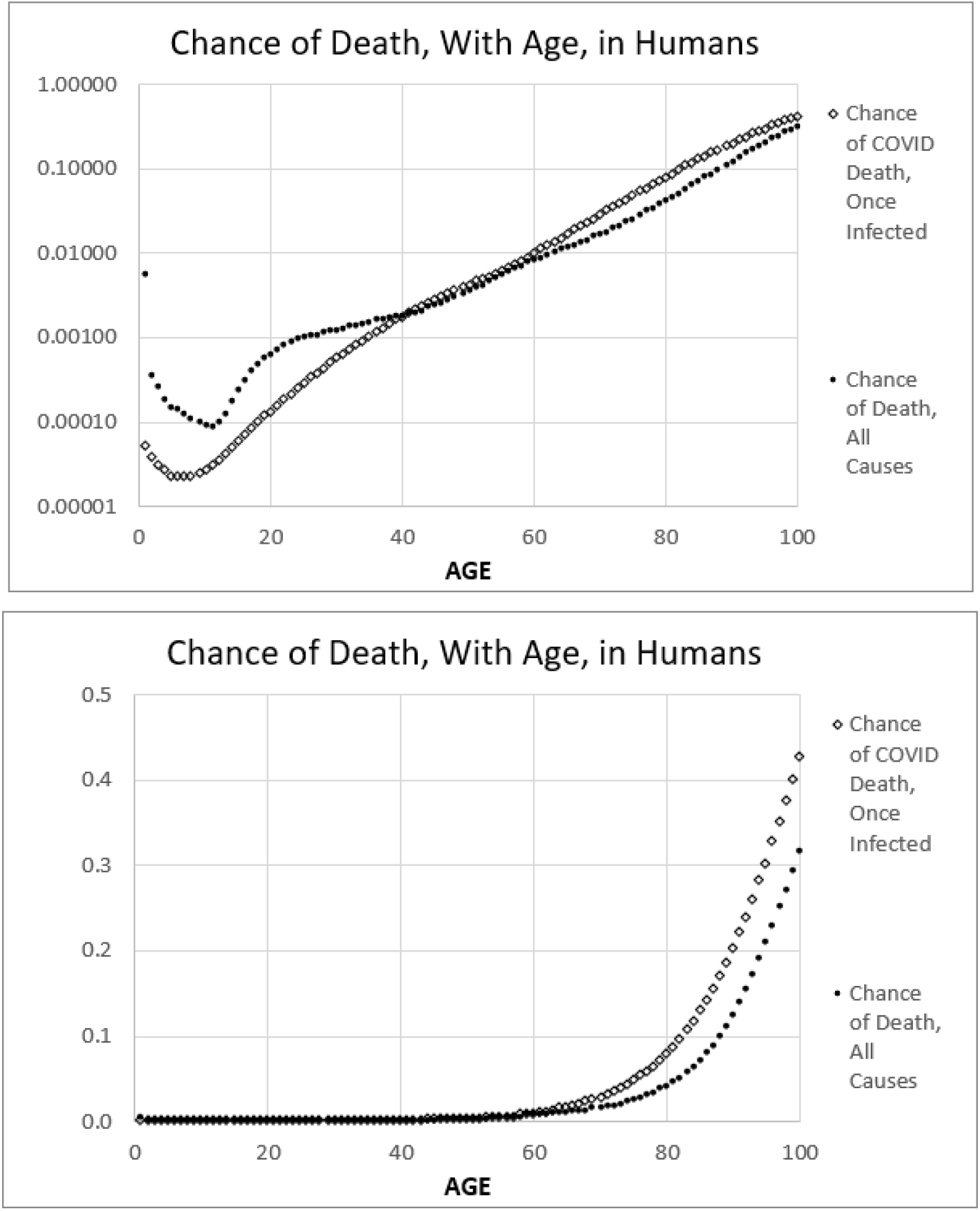
**The *Gompertzian Force of Mortality*^3^** captured by the ***Gompertz Mortality Equation***: ***log*(*D*) =(*G***_***s***_ **· *t*) + log (*G***_***H***_)

### The cellular basis of the ***Gompertz Mortality Equation***

In the accompanying paper, my colleagues and I show that such a ***Gompertz Mortality Equation***, affecting both ageing’s impact on mortality generally, and COVID-19 lethality specifically, can be traced to the slowing of growth that marks maturity.^4^ This could be determined by counting cells, ***N***, as animals grow, a method we have called ***Cellular Phylodynamics***. We used such a ***Cellular Phylodynamic*** analysis to measure the fraction of cells dividing as we age, revealing that that rate at which we age is related to the decline in this ***Mitotic Fraction***. This finding provides a quantitative comprehension for the long-appreciated observation that cell division makes us grow, but it also keeps us well, since biological materials decay with time, a toxic problem that is reversed when cells divide and replace at least half of their components anew. This ***Mitotic Fraction*** mechanism of aging, leading to the decline in ***Mitotic Dilution***, leading to the exponential ***Gompertzian Force of Mortality***, captured by the ***Gompertz Mortality Equation***, lets us link death to cells, in the immune system for COVID-19, and throughout the body for mortality generally, a topic we shall return to later.

### Gompertzian Analysis

Here, measures of COVID-19 ***infectivity*** and ***lethality*** have been teased out of clinical data by sorting patients by age, counting cases and deaths, and displaying these numbers on logarithmic graphs. This new method has been named ***Gompertzian Analysis***.

## METHODS

### *Gompertzian Analysis* gives 3 new measures: *Pasteurian Infectivity, Gompertzian Lethality, Malthusian Lethality*

***Gompertzian Analysis***, that is, sorting patients by age, counting cases and deaths, and displaying these numbers on logarithmic graphs, has made it possible to generate three new practical measures of COVID-19 impact:

* ***Pasteurian Infectivity***: Cases/Population.
* ***Gompertzian Lethality***: Deaths/Cases.
* ***Malthusian Lethality***: Deaths/Population.

All three measures, when compared against age, appear as ***Gompertz Lines*** on logarithmic ***Gompertz Plots***, shown in FIGURES 1 and 2, and many other figures throughout this communication and in the APPENDIX. Thus, each of these three measures can be distilled down to two numbers: the ***Gompertzian Height, G***_***H***_, and ***Gompertzian Slope, G***_***S***_ of the ***Gompertz Mortality Equation***:

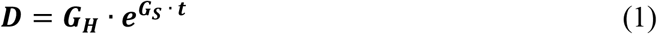

which is equivalent to

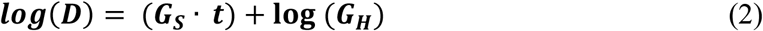

**FIGURE 2.**
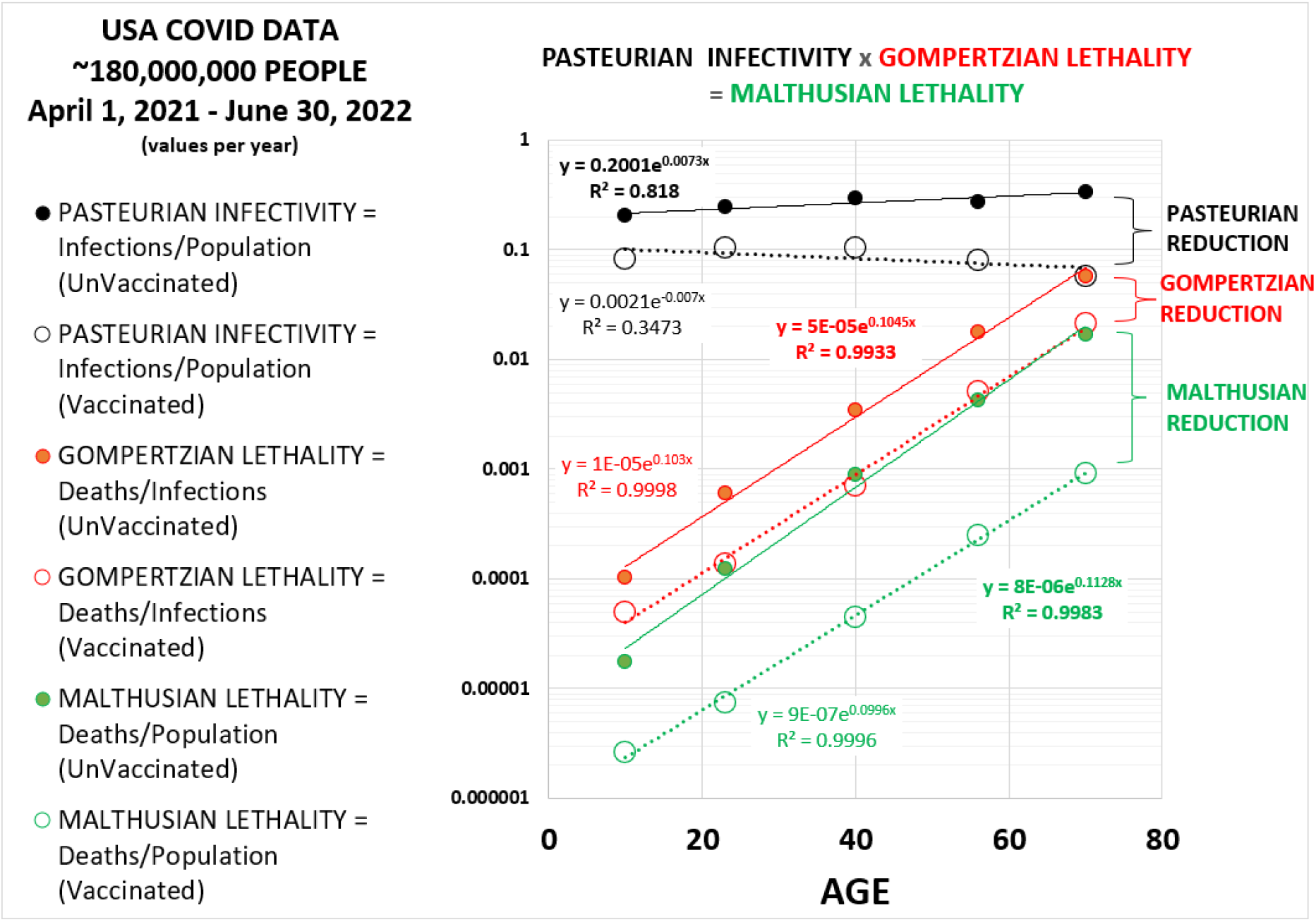
***Gompertzian Lethality, Pasteurian Infectivity***, and ***Malthusian Lethality***, and their ***Reductions*** by Vaccination, USA^14^

Where ***D*** is the chance of death and ***t*** is age. Analysis of data, and calculation of the values of the parameters ***G***_***H***_, and ***G***_***S***_, is determined by examining fit to the exponential equation, which we shall see is usually quite close.

The data will show for ***Gompertzian Lethality*** (Deaths/Cases), and ***Malthusian Lethality*** (Deaths/Population), that there is an ~10,000-fold rise in lethality with age. This critical quality of COVID-19, and ageing, will be examined in detail below.

### *Gompertzian Analysis’s* 3 new measures are each captured by a single number: the *Gompertzian Height, G*_*H*_

As we shall see, ***Gompertzian Analysis*** of a variety of datasets has revealed that vaccines, and other forces, change COVID-19 ***infectivity*** and ***lethality*** by changing the ***Gompertzian Height, G***_***H***_, but not the ***Gompertzian Slope, G***_***S***_. That is to say, vaccines change ***infectivity*** and ***lethality*** by moving down the ***Gompertz Lines*** of ***Pasteurian Infectivity, Gompertzian Lethality***, and ***Malthusian Lethality***, without changing their slopes. This mathematically subtle point is a topic of continuing analysis by my colleagues and I, but, for now, we need only appreciate that this simplifies our analysis of COVID-19 ***infectivity*** and ***lethality***.

Numerically, the data presented here reveal, over and over, that the whatever might be the change in ***Gompertzian Height, G***_***H***_, induced by vaccinations and other forces, ***G***_***SG***_ (the ***Gompertzian Slope*** of ***Gompertzian Lethality***) and ***G***_***SM***_ (the ***Gompertzian Slope*** of ***Malthusian Lethality***), both of which capture the ~10,000-fold rise in lethality with age, has repeatedly been found to have a value of **~0.1**. Similarly, since ***Pasteurian Infectivity*** tends to have similar values among patients of various ages, ***G***_***SP***_ (the ***Gompertzian Slope*** of ***Pasteurian Infectivity***), has repeatedly been found to have a value of **~1.0**.

Thus, ***G***_***HP***_ (the ***Gompertzian Height*** of ***Pasteurian Infectivity***), ***G***_***HG***_ (the ***Gompertzian Height*** of ***Gompertzian Lethality***), and ***G***_***HM***_ (the ***Gompertzian Height*** of ***Malthusian Lethality***), each provide a single measure of COVID-19 ***infectivity*** or ***lethality***. For all three qualities, higher is bad, lower is good.

### *Gompertzian Analysis’s* 3 new measures give population-wide values from partial age samples

***Gompertzian Lethality, Pasteurian Infectivity***, and ***Malthusian Lethality*** are all calculable across the age spectrum, even from incomplete age samples (APPENDIX-FIGURE 6, APPENDIX-FIGURE 7). That is to say, calculations of ***Gompertzian Lethality, Pasteurian Infectivity***, and ***Malthusian Lethality*** will give accurate values for COVID-19 ***infectivity*** or ***lethality*** for patients of all ages, even when these values are calculated from a sample of patients that are young, or old, or contain any mix of ages. With information on the age structure of a population, calculations of ***Gompertzian Lethality Pasteurian Infectivity***, and ***Malthusian Lethality***, even from small samples of patients that don’t contain a full sample of all ages, will give values of the population-wide burden of ***infectivity*** and ***lethality***, that is to say, population-wide values for number of cases and deaths, when the actual population-wide number of cases and deaths are unknown.

### *Gompertzian Lethality* and *Case Fatality*

Both ***Gompertzian Lethality*** and ***Case Fatality*** measure (Deaths/Cases). However, because (Deaths/Cases) displays an ~10,000-fold exponential rise in lethality with age, even slight differences in ages of the individuals in a group will skew the measure of ***Case Fatality***. In contrast, this does not affect ***Gompertzian Lethality***, which provides a number, ***G***_***HG***_, the ***Gompertzian Height*** of ***Gompertzian Lethality*** of COVID-19 ***lethality***, which is independent of patient age. Such a calculation was not available before.

### *Gompertzian Analysis* gives 3 additional new measures for measuring impact of vaccination and other forces: *Reductions*

***Gompertzian Analysis*** gives 3 additional new measures for measuring impact of vaccination and other forces on COVID-19 called ***Reductions***: For vaccination, the ***Pasteurian Infectivity Reduction, Gompertzian Lethality Reduction***, and ***Malthusian Lethality Reduction***, was calculated for each age group as “ 1-(Vaccinated/Unvaccinated)”. All of the age groups were then averaged. ***Reductions*** are typically expressed as “ ***percentages***” but may also be expressed in terms of “ ***folds***” or “ ***chances***” when this improves the clarity of the meaning of the ***Reductions*** (see APPENDIX for details). These three new measures generated by ***Gompertzian Analysis*** - ***Pasteurian Infectivity Reduction, Gompertzian Lethality Reduction***, and ***Malthusian Lethality Reduction***-add additional information not available before from measures such as ***Vaccine Effectiveness***.

### *Pasteurian Infectivity Reduction*: The Effect of Vaccines on the Risk of Infection

***Pasteurian Infectivity Reduction*** and ***Vaccine Effectiveness Against Infection*** are both calculated in terms of 1-vaccinated/unvaccinated (Cases/Population). Because ***Pasteurian Infectivity*** occurs at similar rates across ages, the two measures are roughly equivalent.

### *Malthusian Lethality Reduction*: The Effect of Vaccines on the Risk of Infection and Death

***Malthusian Lethality Reduction*** and ***Vaccine Effectiveness Against Death*** are both calculated in terms of 1-vaccinated/unvaccinated (Deaths/Population). However, because (Deaths/Population) displays an ~10,000-fold exponential rise in lethality with age, even slight differences in ages of the patients in the vaccinated and unvaccinated groups may lead to imprecision in the measure of ***Vaccine Effectiveness Against Death***. In contrast, this does not affect ***Malthusian Lethality Reduction***, which provides a measure of vaccine impact that is independent of patient age.

### *Gompertzian Lethality Reduction*: The Effect of Vaccines on Risk of Death Once Infected

***Gompertzian Lethality Reduction*** provides the measure for how vaccines, and other forces, affect just COVID-19 ***lethality***. Such a calculation was not available before.

### *Gompertzian Analysis* with Full Age Information

The public data used here are provided in age-group categories, usually open-ended for the youngest and oldest groups, which reduces the potential precision of ***Gompertzian Analysis*** to characterize the parameters of ***Gompertzian Lethality, Pasteurian Infectivity, Malthusian Lethality***, and their ***Reductions*** by vaccination, and other forces. Fortunately, more accurate measures of the parameters of the ***Gompertz Mortality Equation*** can be made when the age of each patient is known, rather than placed in an age-group, as has been described by Meuller et al^5^ and others. Those with direct access to larger datasets, and thus with knowledge of the age of each case and death, should benefit from this opportunity.

## RESULTS

### COVID-19 *Lethality*

#### COVID-19 displays an ~10,000-fold exponential rise in lethality with age: the *Gompertzian Force of Mortality*

***Gompertzian Analysis*** of a variety of datasets (see APPENDIX for references) has revealed that COVID-19 ***Gompertzian Lethality*** (deaths/cases) captures an ~10,000-fold exponential increase in the chance of death from infancy to old age (FIGURES 1, 2, many other figures here and in the APPENDIX). Such an exponential increase in the chance of death with age, discovered for all-cause mortality by ***Gompertz***^1^ ***in 1835, and for COVID-19 by Levin et al***^***2***^ ***in 2020, is called the Gompertzian Force of Mortality***. The ***Gompertzian Force of Mortality*** is captured by the ***Gompertz Mortality Equation*** (#1 and #2) and can be seen as straight ***Gompertz Lines*** on logarithmic ***Gompertz Plots*** throughout this report.

#### The *Gompertzian Force of Mortality* describes COVID-19, and other diseases, and mortality generally

COVID-19’s ~10,000-fold exponential rise in lethality with age is but one example of the overall increase in the chance of death that occurs as we age, as well as for most of the individual diseases which cause death as we age. ***Gompertz Plots*** for COVID-19, infectious hepatitis, and influenza^6^ can be seen in APPENDIX-FIGURE 4, while ***Gompertz Plots*** for deaths to cirrhosis, nephrosis, hypertension, and heart disease,^6^ can be seen in APPENDIX-FIGURE 5.

#### COVID-19’s Fearful *Gompertzian Force of Mortality* Comes Into View When Displayed Linearly

The ~10,000-fold exponential increase in ***Gompertzian Lethality*** (deaths/cases) from infancy to old age becomes shocking when visualized on linear graphs rather than on logarithmic ***Gompertz Plots***. Data from the beginning of the pandemic, shown in FIGURE 1 and APPENDIX-FIGURE 3, visualizes that from infancy to age 60, we stealthily accumulate less than 1% chance of death if infected, but from that age on, we see the progressively more catastrophic, exponential, increase in COVID-19 lethality, with risk of death rising to about 3% at age 70, to almost 10% at age 80, to almost 20% at age 90, and to almost 50% at age 100.

**FIGURE 3.**
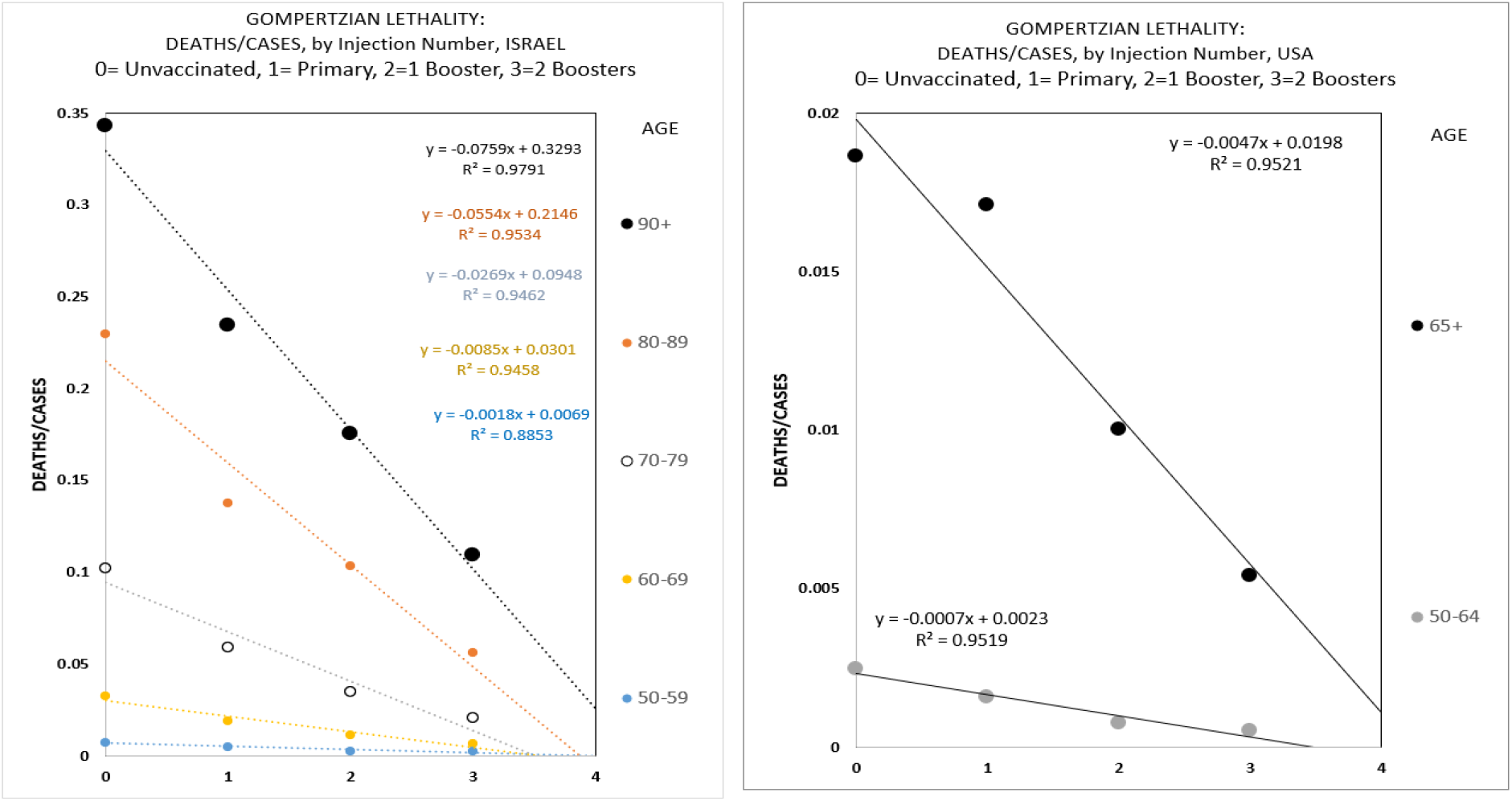
Decline in ***Gompertzian Lethality*** (Deaths/Cases) By Number of Vaccine Doses. Data from Israel^10^ and USA^9^

### COVID-19 *Infectivity*

#### COVID-19 infection occurs at similar rates across ages

***Gompertzian Analysis*** of a variety of datasets (see APPENDIX for references) has revealed that while the chance of COVID-19 death (***Gompertzian Lethality***, Deaths/Cases) is ~10,000-fold higher in old age than in youth, the chance of COVID-19 infection (***Pasteurian Infectivity***, Cases/Population) varies little from age to age (FIGURE 2 and many other figures in the APPENDIX). Old people aren’t noticeably more likely to get infected by COVID-19, but they are catastrophically more likely to die of COVID-19 if infected.

### Non-immunological influences on COVID-19 *Lethality* and *Infectivity*

#### Men have higher levels of *Gompertzian Lethality* than women but similar levels of *Pasteurian Infectivity*

***Gompertzian Analysis*** reveals that men have a higher level of COVID-19 ***Gompertzian Lethality*** (Deaths/Cases), in each age group, than women, taking the form of a higher ***Gompertzian Height, G***_***H***_, with both sexes having a somewhat similar ***Gompertzian Slope, G***_***S***_ (APPENDIX-FIGURE 42).^7^ There is no evident male/female difference in ***Pasteurian Infectivity*** (Cases/Population).

#### Diabetics have higher levels of *Gompertzian Lethality* and *Pasteurian Infectivity*

***Gompertzian Analysis*** reveals that diabetics have higher COVID-19 ***Gompertzian Lethality*** and ***Pasteurian Infectivity***, in each age group.^8^ These higher levels take the form of higher ***Gompertzian Height, G***_***H***_, with no evident change in ***Gompertzian Slope, G***_***S***_. (APPENDIX-FIGURE 43).

### COVID-19 Vaccination

#### Vaccination reduces the chance of death once infected

Vaccination reduces the chance of COVID-19 death once infected. With data from many datasets (see APPENDIX for references), ***Gompertzian Analysis*** reveals that this decline takes the form of ***Gompertz Lines*** of ***Gompertzian Lethality*** (deaths/cases) being markedly lower for vaccinated individuals than for unvaccinated individuals, while these progressively lower ***Gompertz Lines*** remain roughly parallel (FIGURE 2, and others throughout the APPENDIX). Thus, vaccination’s reduction in ***Gompertzian Lethality*** takes the form of a decline in ***Gompertzian Height, G***_***H***_, with little evident change in ***Gompertzian Slope, G***_***S***_.

#### Vaccination reduces the chance of death cumulatively by dose, linearly pointing to zero death after 3 or 4 boosters

When displayed linearly, COVID-19 ***Gompertzian Lethality***, the chance of death if infected, declines in each age group cumulatively with each dose, linearly pointing to zero death after 3 or 4 boosters (FIGURE 3). We have seen this linear decline in COVID-19 ***Gompertzian Lethality*** with each additional dose from ***Gompertzian Analysis*** of data from the CDC’s USA population-wide data collection effort^9^ (FIGURE 3), of data from Hungary (APPENDIX-FIGURES 33 and 34), and of data from Israel^10^ (FIGURE 3; see also APPENDIX-FIGURES 29, 31, 32). A variety of other more recent studies, although with less complete data, have also shown the same effect (see APPENDIX for details).

#### Vaccination reduces the chance of death cumulatively, without signs of waning

The vaccine-induced reduction in COVID-19 ***Gompertzian Lethality*** shows no sign of waning, although the data on this point are limited. A telling indication of this can be seen among the patients in the CDC data from March 2022 to July 2022, who were identified as having 1 booster (APPENDIX-FIGURE 27). This group’s ***Gompertzian Lethality***, which was lowered by primary vaccination followed by a booster, showed no change over this 4-month period, indicating that the vaccine’s ability to reduce the chance of death doesn’t wane noticeably over time (See APPENDIX for details).

#### We will soon know whether sequential dosing pointing to zero death after 3 or 4 boosters actually gets to zero death

While COVID-19 ***Gompertzian Lethality*** declines cumulatively with each dose, for each age group, linearly ***pointing*** to zero death after 3 or 4 boosters, we have yet to see whether it actually ***gets*** there. Fortunately, since 3^rd^ boosters have now been given widely, we shall soon have the data to answer this question.^11,12^

#### Vaccination reduces the chance of infection, but not enough

***Gompertzian Analysis*** of a variety of COVID-19 datasets has revealed that the ***Gompertz Line*** of ***Pasteurian Infectivity*** (Cases/Population) is lower for vaccinated individuals than for unvaccinated individuals (FIGURE 2, and others throughout the APPENDIX). However, while primary vaccination reduces the ***Gompertz Line*** of ***Pasteurian Infectivity*** (Cases/Population), boosters administered subsequently were not accompanied by any additional reduction in ***Pasteurian Infectivity*** (APPENDIX-FIGURE 30). That is to say, while sequential vaccination reduces COVID-19 ***Gompertzian Lethality***, and does so cumulatively with each additional dose, vaccines also reduce ***Pasteurian Infectivity***, but do not accumulate further reductions with additional doses, and do not do so without waning. As a result, the reduction in COVID-19 ***infectivity*** achieved by vaccination has not been enough to eradicate transmission in the population.

#### Age has no evident impact on vaccine’s abilities to reduce infection or death

When we examine the ***Gompertz Lines*** of ***Pasteurian Infectivity*** (Cases/Population) or ***Gompertzian Lethality*** (Deaths/Cases) of vaccinated individuals compared to unvaccinated individuals, on logarithmic ***Gompertz Plots***, over and over, we see that such lines are roughly parallel (FIGURE 2, and numerous figures in the APPENDIX). This indicates that vaccines reduce infection and death in the young and the old equally, although doing so from a ***Gompertzian Lethality*** starting point that is more challenging for the old than the young.

#### Different vaccines have different reductions in COVID-19 *infectivity* and *lethality*

***Gompertzian Analysis*** of a variety of datasets has found that different vaccines have different reductions in ***Gompertzian Lethality*** and ***Pasteurian Infectivity***. Perhaps the most striking data comes from the studies of Hungarian patients.^13^ As can be seen in APPENDIX-TABLE I, the vaccines with the highest and lowest ***Pasteurian Reductions*** differed more than ***5-fold*** in their capacity to reduce infection, and vaccines with the highest and lowest ***Gompertzian Reductions*** differed ***2-fold*** in their capacity to reduce the occurrence of death once infected. The combined difference of the highest and lowest ***Malthusian Reductions*** differed ***8-fold*** in their capacity to reduce the occurrence of death in the populations as a whole.

#### Boosters: Who’s Dying Now?

The CDC vaccination and booster status data on people with COVID-19 infections from March 2022 through July 2022^9^ give us a picture of how deaths are occurring among the unvaccinated, among those with only primary vaccinations, among those who also have had 1 booster, and among those that have had an additional, 2^nd^ booster (APPENDIX-TABLE V). Those with 2 boosters cut their chance of COVID-19 death in half, in comparison to those with only 1 booster, and had only ~1/10^th^ of the chance of dying of COVID-19 in comparison to those without vaccination. Of course, 3^rd^ boosters have been administered now, and the projected values shown here suggest the possibility of negligible lethality; we await the data to see whether this hopeful possibility is realized.

### COVID-19 2021-2022

CDC data on approximately 70% of all COVID-19 cases in the USA from April 2021 to June 2022, makes it possible to break down patients by age, and count cases and deaths, to give long-term ***Gompertzian Analysis*** assessments of ***Pasteurian Infectivity*** (Cases/Population), ***Gompertzian Lethality*** (Deaths/Cases), and ***Malthusian Lethality*** (Deaths/Population).^14^

#### *Gompertzian Lethality* went up and down mildly, ending 10-fold lower

For both vaccinated and unvaccinated individuals, ***Gompertzian Analysis*** has found that ***Gompertzian Lethality*** (Deaths/Cases), in each age group, went up and down mildly with time, with small waves that varied several-fold in height (APPENDIX-FIGURES 22-25). Over the long term, ***Gompertzian Lethality*** declined roughly 10-fold from April 2021 to June 2022(FIGURE 4).

**FIGURE 4.**
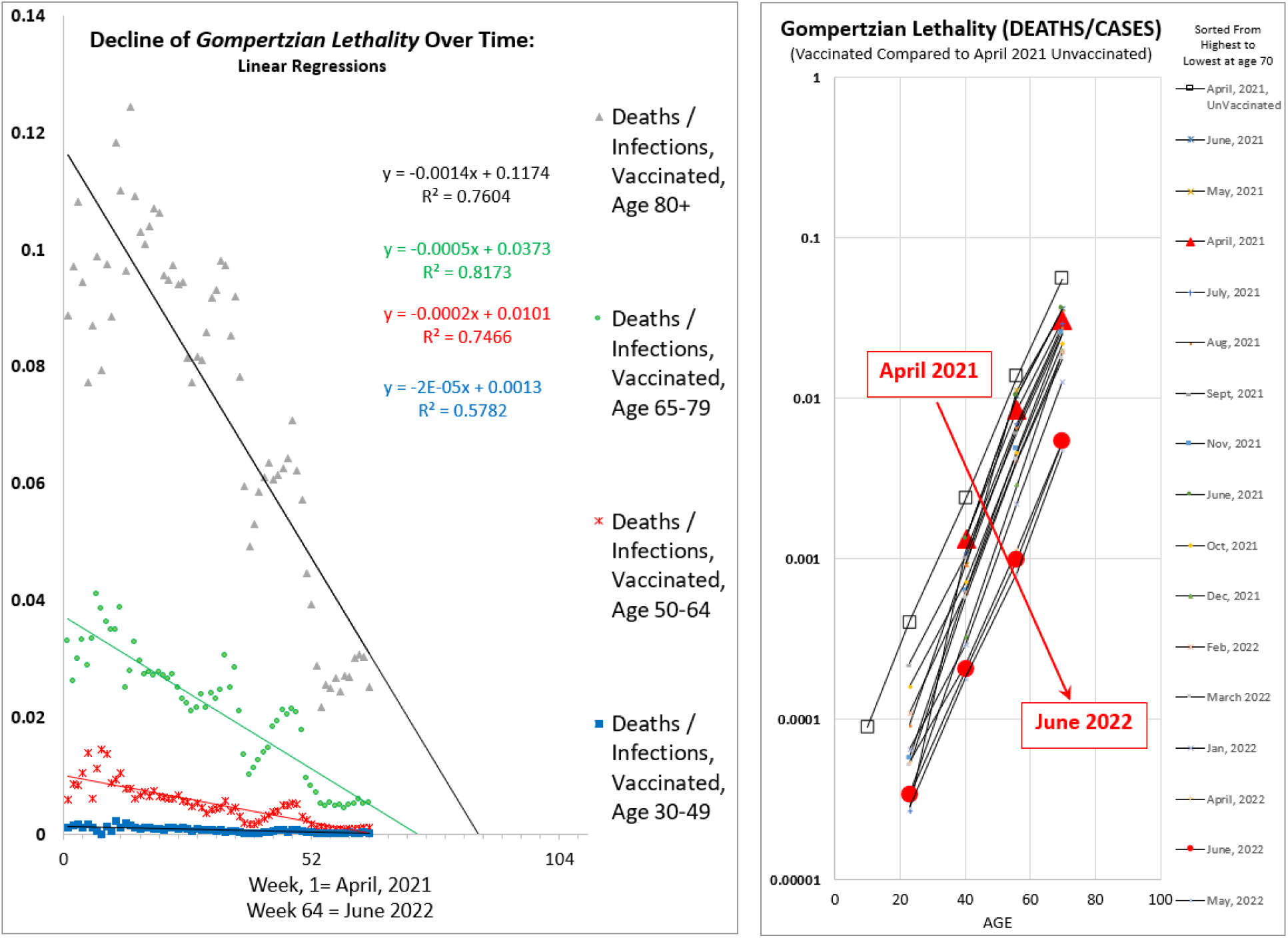
Decline in ***Gompertzian Lethality*** April 2021 to June 2022, USA, by age.^14^ (Left) Linearly, date on X-axis; (Right) Logarithmically, age on X-axis

#### *Gompertzian Lethality* of vaccinated is heading towards zero deaths

For the vaccinated, when examined on linear graphs, ***Gompertzian Lethality*** (Deaths/Cases), in all age groups, declined linearly over time, pointing to zero deaths over the next few months (FIGURE 4 and APPENDIX-FIGURE 23). To what degree this decline in ***Gompertzian Lethality*** was driven by boosters, increased immunity by breakthough infections, new variants, or other factors, isn’t evident from this dataset. Whether we actually get to negligable lethality among the vaccinated, and especially among those fully vaccinated with all possible boosters, will soon become clear.

#### *Gompertzian Lethality* among the vaccinated declined by *Gompertzian Height*

Graphs of the log of ***Gompertzian Lethality*** (Deaths/Cases) vs (age) for monthly groups of vaccinated COVID-19 patients revealed ***Gompertz Lines*** of ***Gompertzian Lethality*** declining progressively in ***Gompertzian Height*** from April 2021 to June 2022, with little evident change in the ***Gompertzian Slope*** (FIGURE 4 and APPENDIX-FIGURE 25). That is to say, ***Gompertzian Lethality*** declined by roughly equivalently improvements in each age group.

#### *Pasteurian Infectivity* went up and down wildly, ending 10-fold higher

For both vaccinated and unvaccinated individuals, ***Gompertzian Analysis*** has found that ***Pasteurian Infectivity*** (Cases/Population) went up and down wildly with the huge waves, varying over a hundred-fold in height from April 2021 to June 2022 (APPENDIX-FIGURES 22-24). Over the long term, ***Pasteurian Infectivity*** displayed a roughly a 10-fold increase in the chance of infection. Vaccinated people had lower rates of ***Pasteurian Infectivity*** (Cases/Population) than unvaccinated people, but the virus kept defeating the vaccines.

Comparison of ***Gompertzian Lethality*** with ***Pasteurian Infectivity*** reveals how much progress has occurred in reducing COVID-19 lethality, and how little in progress has occurred in reducing COVID-19 infection. This is illustrated in FIGURE 5 for Age 65-79 individuals, but it is seen in all age groups (APPENDIX-FIGURE 24).

**FIGURE 5.**
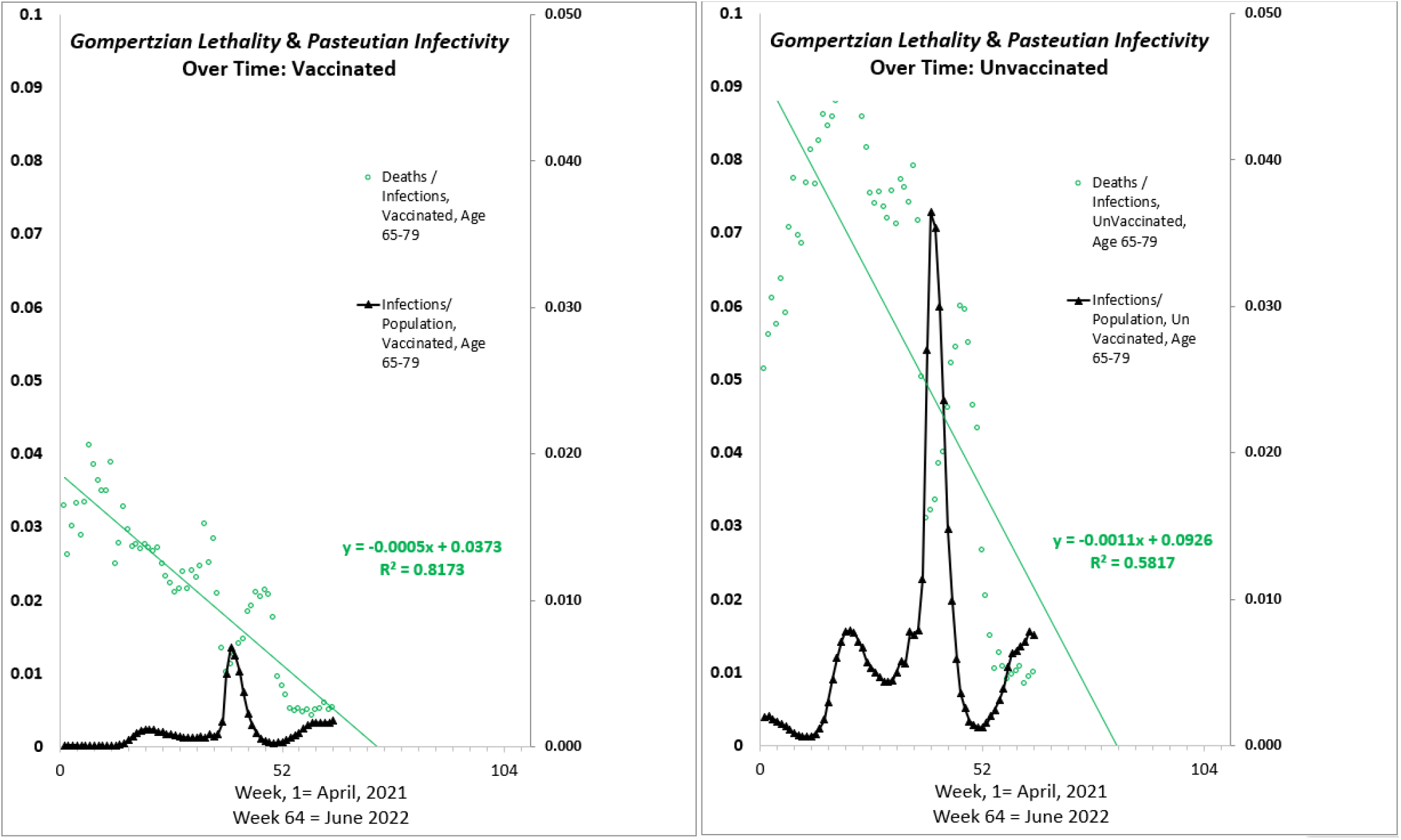
Evolution of ***Gompertzian Lethality*** (Deaths/Cases) and ***Pasteurian Infectivity*** (Cases/Population), April 2021 to June 2022, USA, Age 65-79. (Left) vaccinated. (Right) unvaccinated.^14^

### The unvaccinated also underwent a decline in *Gompertzian Lethality*

***Gompertzian Analysis*** has shown that the unvaccinated also saw a reduction in ***Gompertzian Lethality*** (Deaths/Cases) from April 2021 to June 2022 (APPENDIX-FIGURES 22-24). The decline was less than occurred among the vaccinated, and in each age group, in each week from April 2021 to June 2022, the vaccinated always had a lower level of ***Gompertzian Lethality*** and ***Pasteurian Infectivity*** than the unvaccinated

This decline in ***Gompertzian Lethality*** among the unvaccinated was likely the result of infection, as could also be seen in an Israel cohort of recovered COVID-19 patients shown in APPENDIX-TABLE III and APPENDIX-FIGURE IX. If so, the sad price for the reduction in the ***Gompertzian Lethality*** among the unvaccinated would be the loss of life occurring after some of these infections; by June 2022, unvaccinated Americans were roughly 25% of the populations, but had 75% of the COVID-19 deaths (APPENDIX-FIGURES 18, 19, and 21).

### With new variants, *Gompertzian Lethality* declined while *Pasteurian Infectivity* increased

***Gompertzian Analysis*** of two datasets of patients (UK^15^ and USA^16^) infected by different variants has revealed a lower level of ***Gompertzian Lethality*** (Deaths/Cases), in each age group, for ***Delta***, and a lower level yet for ***Omicron*** (FIGURE 6 and APPENDIX-FIGURES 45 AND 46). In contrast, ***Pasteurian Infectivity*** (Cases/Population) has increased with each new variant. Previous experience teaches that viral Darwinism would select for variants to evolve to be more infectious (***Pasteurian Infectivity***), but not necessarily more deadly (***Gompertzian Lethality***), and that is what appears to have been the case so far.

**FIGURE 6.**
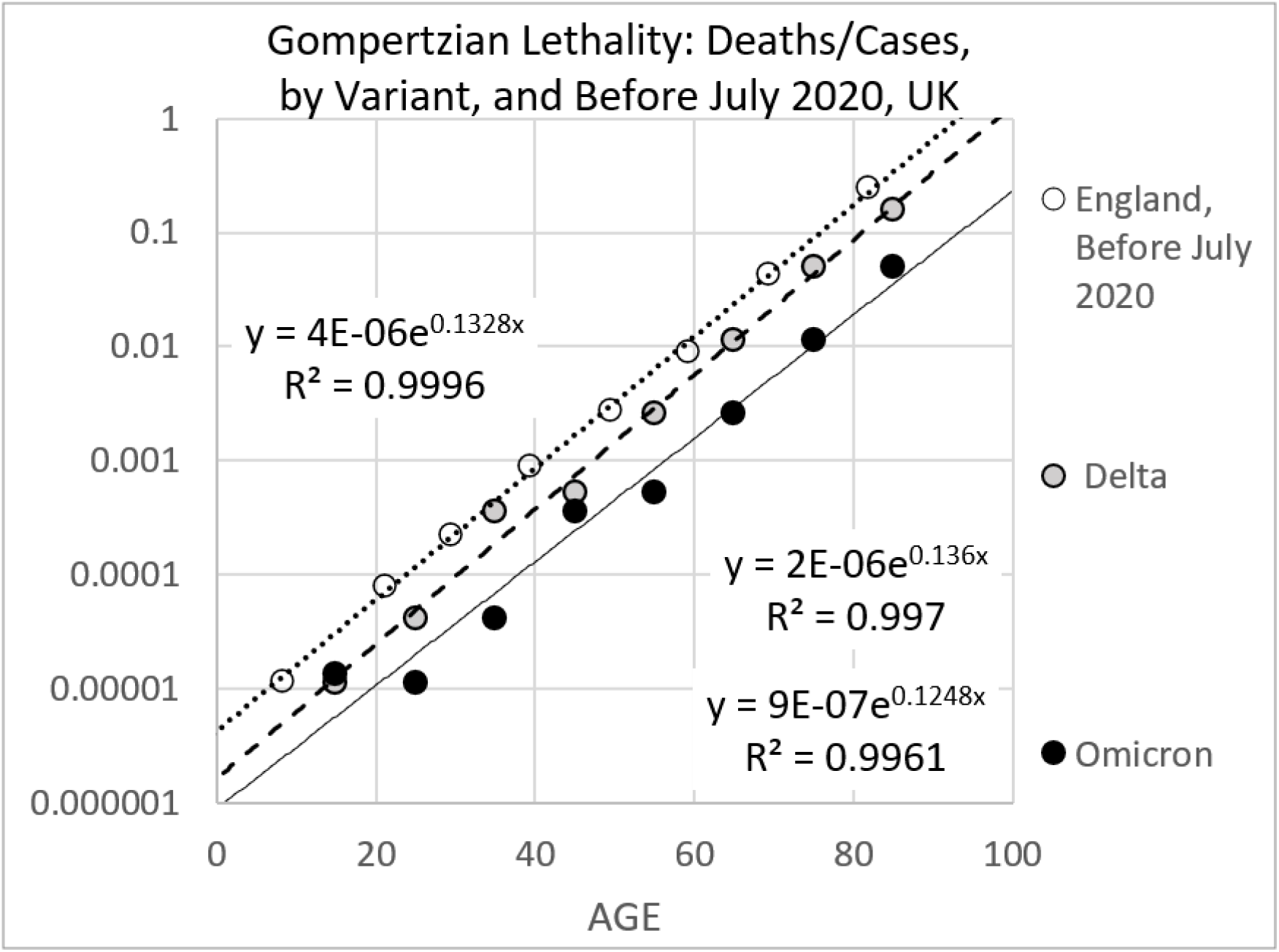
***Gompertzian Lethality*** (Deaths/Cases) By Variant, UK^15^

### LONG COVID and long-term pandemic associated excess mortality

Unlike ***Gompertzian Lethality, Long COVID*** manifests in multiple symptoms, which appear and last after infection,^17,18^ as does long term pandemic associated excess mortality.^19,20,21,22^ ***Gompertzian Analysis*** has found that unlike ***Gompertzian Lethality, Long COVID*** occurs at relatively equivalent rates across ages (APPENDIX-FIGURE 40).^23,24^ Vaccination has been found to protect against ***Long COVID***, (APPENDIX-FIGURE 41),^25,26,27,28^ with protection increasing with each additional dose,^29^ although the data so are far not sufficient to determine which symptoms are prevented, and most importantly, to what degree pandemic associated excess mortality is reduced, if at all, but vaccination.^30,31^ Clearly, this remains a problem of high importance, which should be decipherable by ***Gompertzian Analysis***.

## DISCUSSION

### COVID-19 vaccination reduces *Gompertzian Lethality* (Deaths/Cases), pointing to zero death after 3 or 4 boosters

Here we have seen, simply by counting cases and deaths, by age, and displaying these numbers on logarithmic graphs, that is, ***Gompertzian Analysis***, that COVID-19 sequential vaccination reduces ***Gompertzian Lethality*** (Deaths/Cases, sorted by age), pointing linearly towards zero death after 3 or 4 boosters, without signs of waning (FIGURE 4). More counting will tell us whether we actually get there, whether this protection actually doesn’t wane, whether it actually lasts as new variants arrive, whether those who have had infections might require fewer boosters, while those who have gone without infections might require more boosters, whether those who have missed some boosters can get caught up with late boosters, whether we should continue to add more booster rounds for all, and on, and on. Counting cases and deaths is key to expanding our understanding of COVID-19 and defeating its lethal potential.

### Zero deaths and negligible deaths

We can see in FIGURE 4 how, with each booster, ***Gompertzian Lethality*** (Deaths/Cases, sorted by age), points linearly towards zero death, which, mathematically, as shown in APPENDIX-FIGURES 31-32, 34, 35-39, points exponentially to negligible death, asymptotically approaching zero. The distinction is subtle, and worthy of more study, but from a practical standpoint, irrelevant; get 3 or 4 boosters, and the risk of COVID-19 death can be forgotten. But, of course, continuing to count cases and deaths, so as to test this possibility, and quantify its subtleties, as noted above, cannot be forgotten.

### COVID-19 vaccination reduces *Pasteurian Infectivity* (Cases/Population), but not enough

***Gompertzian Analysis*** also revealed that that COVID-19 vaccination reduces ***Pasteurian Infectivity*** but doesn’t defeat the virus (FIGURE 5, and many more in the APPENDIX). However, ***Gompertzian Analysis*** has also shown that some vaccines are better than others, and that the math might be able to help us do better, a point to be returned to below.

### *Gompertzian Analysis* has revealed additional useful findings

**FIRST:** COVID-19 ***Gompertzian Lethality*** (Deaths/Cases) exhibits an ~10,000-fold exponential increase in the chance of death with age, the ***Gompertzian Force of Mortality***, captured by the ***Gompertz Mortality Equation*. SECOND:** The ***Gompertz Mortality Equation*** appears on logarithmic ***Gompertz Plots*** as straight ***Gompertz Lines*** of death, ***D***, vs age, ***t***, captured by two numbers: the ***Gompertzian Slope, G***_***s***_, and the ***Gompertzian Height, G***_***H***_. **THIRD:** The ***Gompertzian Force of Mortality*** characterizes COVID-19, other diseases, and all-cause mortality, possibly from loss of ***Mitotic Dilution*** of toxic compounds due to decline in mitosis that occurs with growth and aging. **FOURTH:** Most lethal diseases, COVID-19, and the changes induced in COVID-19 ***Gompertzian Lethality*** by vaccination and other forces, are each captured by a different ***Gompertzian Height, G***_***H***_ but a similar ***Gompertzian Slope, G***_***s***_. **FIFTH:** COVID-19 ***Pasteurian Infectivity*** (Cases/Population) occurs at similar rates across ages. **SIXTH:** Men have higher levels of COVID-19 ***Gompertzian Lethality*** than women, captured by a higher the ***Gompertzian Height, G***_***H***,_ but similar levels of ***Pasteurian Infectivity*. SEVENTH:** Diabetics have higher levels of COVID-19 ***Gompertzian Lethality*** and ***Pasteurian Infectivity***, captured by a higher the ***Gompertzian Height, G***_***H***_. **EIGHTH:** COVID-19 vaccines reduce ***Gompertzian Lethality*** (Deaths/Cases), by lowering the ***Gompertzian Height, G***_***H***_, but do not chage the ***Gompertzian Slope, G***_***s***_. **NINTH** Over the past several years, COVID-19 ***Gompertzian Lethality***, has declined, but not ***Pasteurian Infectivity*. TENTH:** With each variant, COVID-19 ***Gompertzian Lethality*** has declined, but not ***Pasteurian Infectivity*. ELEVENTH:** The unvaccinated have seen a decline in COVID-19 ***Gompertzian Lethality***, less than the vaccinated, ascribable to infection, at the cost of lives lost.

### Resistance to COVID-19 *infectivity* and *lethality* appear to be separate processes

Multiple findings of ***Gompertzian Analysis*** are suggestive of the hypothesis that resistance to COVID-19 ***Pasteurian Infectivity*** (Cases/Population) and susceptibility to ***Gompertzian Lethality*** (Deaths/Cases) may arise from two separate responses to the virus from two separate parts of the immune system. **FIRST**: Perhaps the most dramatic difference is that COVID-19 ***Pasteurian Infectivity*** (chance of infection) occurs at similar rates of among the young and old, while COVID-19 ***Gompertzian Lethality*** (chance of death once infected) displays its ~10,000-fold increase in the from young to old. **SECOND**: sequential vaccination leads to progressively cumulative resistance to ***Gompertzian Lethality***, linearly pointing to negligible death after 3 or 4 boosters, without signs of waning, ultimately defeating the virus’s capacity to kill, and while vaccination leads to a reduction in ***Pasteurian Infectivity***, it isn’t cumulative, and it doesn’t defeat the virus. **THIRD**: For various vaccines, the reductions in ***Gompertzian Lethality*** and ***Pasteurian Infectivity*** don’t seem to occur in parallel. **FOURTH**: a variety of forces have different impact on ***Gompertzian Lethality*** and ***Pasteurian Infectivity***. Female-to-male comparisons show lower levels of ***Gompertzian Lethality*** among women than men, but similar levels of ***Pasteurian Infectivity***. On the other hand, diabetics have higher rates of both ***Gompertzian Lethality*** and ***Pasteurian Infectivity***.

### The cells that control *infectivity* and *lethality* can be identified by counting them

Might COVID-19 ***Pasteurian Infectivity Reduction*** be accomplished by the immune system’s B-cells, while the ***Gompertzian Lethality Reduction*** be the work our T-cells?^32^ Whether or not my guess proves to be correct, the locations for ***Pasteurian Infectivity Reduction*** and ***Gompertzian Lethality Reduction*** can be identified by counting cells of the immune system, as they change with age and infection, with the math described in the accompanying manuscript.^4^

### How much *Pasteurian Infectivity Reduction* is needed to bring the pandemic under control?

We are now in a position to relate the mathematics of ***Pasteurian Infectivity*** and ***Pasteurian Reduction*** to COVID-19 pandemic transmission, by examining the relationship between the ***Gompertzian Height*** parameter of ***Pasteurian Infectivity, G***_***HP***_, to the ***R***_***o***_ parameter of pandemic mathematics. Such math would appear to be doable, and could be critical to learning what level of ***Pasteurian Reduction*** would be necessary to bring the pandemic under control, if, indeed, it can be brough under control. These calculations should also allow us to see how changes in variant properties, population immunological resistance, and vaccine implementation, would impact achieving such a goal.

### Optimizing vaccine reduction of *Pasteurian Infectivity*

The current failure of vaccines to achieve a satisfactory reduction in ***Pasteurian Infectivity*** is disappointing but not hopeless. In the accompanying manuscript,^4^ my colleagues and I address the question of how counting cells of the immune system, in animals, and in humans, could be used to extract a mathematics of vaccine immunization for identifying, rationally, the optimal dosages, schedules, composition, adjuvantation, pharmaceutical augmentation, and other aspects of vaccine implementation, so as to maximized the vaccine’s ***Reduction*** in ***Pasteurian Infectivity***. These calculations should also allow us to examine vaccine action at single moments in time, and in the presence of evolving variants.

### The cellular basis of age’s impact on COVID-19 *Lethality*

In this manuscript, we have seen how counting cases and deaths reveals that the risk of COVID-19 ***Gompertzian Lethality***, and indeed many other causes of death, increases exponentially with age, the ***Gompertzian Force of Mortality***, captured by the ***Gompertz Mortality Equation***, while in the accompanying manuscript, we have seen how counting our cells as we grow reveals that this ***Gompertzian Force of Mortality*** is correlated with the decline in the fraction of our cells dividing, the ***Mitotic Fraction***, which occurs as we settle into our final size and years.^4^ Indeed, by counting our cells, it has been possible to put into quantitative terms, how, and why, and when, we grow, from single fertilized cells into adults of full size and shape, how the control of cell division harnesses our unruly cells into well behaved multicellular animals, and how this process ultimately makes us old and ill.^33,34,35^ This math reveals that our lifespan is correlated with an age when fewer than one-in-a-thousand cells are dividing, quantifying the long-appreciated mechanism of aging, the failure of cells to be rejuvenated by dilution with new materials made, and DNA repaired, at mitosis.^36,37,38^ This math thus links COVID-19 ***Lethality*** to our cells, and as noted above, gives us a way to find which cells account for COVID-19 ***Pasteurian Infectivity Reduction*** and which cells account for ***Gompertzian Lethality Reduction***, as well as to how make the calculations needed to optimize COVID-19 vaccination.

### COVID can teach us about aging: Aging can teach us about COVID

The ~200,000 research articles published on COVID-19 comprise the largest gerontology data collection project in the history of demography.^39^ We should not miss this opportunity to tease out COVID-19’s various age-related features, and search for their cellular origins. ***Gompertzian Analysis*** seems to be telling that COVID-19 provides the ideal framework for understanding aging’s ***Force of Mortality***, while aging’s ***Force of Mortality*** provides the ideal framework for understanding, and reducing, COVID-19’s lethal burden.

Note: this study has not been peer reviewed, and the findings could change.

## Data Availability

All data were extracted either from supplements to papers cited, or from open access datasets, also cited

## References

1 Gompertz, B. On the nature of the function expressive of the law of human mortality, and on a new mode of determining the value of life contingencies. Philosophical Transactions of the Royal Society of London. 115: 513–585. (1825). “ doi:10.1098/rstl.1825.0026. S2CID 145157003.

2 Levin AT, Hanage WP, Owusu-Boaitey N, Cochran KB, Walsh SP, Meyerowitz-Katz G. Assessing the age specificity of infection fatality rates for COVID-19: systematic review, meta-analysis, and public policy implications. Eur J Epidemiol. 2020 Dec;35(12):1123–1138. doi: 10.1007/s10654-020-00698-1. Epub 2020 Dec 8. PMID: 33289900; PMCID: PMC7721859.

3 COVID-19 Forecasting Team. Variation in the COVID-19 infection-fatality ratio by age, time, and geography during the pre-vaccine era: a systematic analysis. Lancet. 2022 Apr 16;399(10334):1469–1488. doi: 10.1016/S0140-6736(21)02867-1. Epub 2022 Feb 24. Erratum in: Lancet. 2022 Apr 16;399(10334):1468. PMID: 35219376; PMCID: PMC8871594.

4 Citi L, Su, J, Huang, L, Michaelson, J. Counting Cells By Age Tells Us How, and Why, and When, We Grow, and Become Old and Ill. 2022. In Preparation.

5 LD Mueller, T J Nusbaum, M R Rose. The Gompertz equation as a predictive tool in demography. Exp Gerontol. 1995 Nov-Dec;30(6):553–69. doi: 10.1016/0531-5565(95)00029-1.

6 Jones HB, A SPECIAL CONSIDERATION OF THE AGING PROCESS, DISEASE, AND LIFE EXPECTANCY November 1, 1955 CLEVELAND PUBLIC LIBRARY TECHNOLOGY DIVISION MAR 2 1956 SERIAL DOCS OARDS Printed for the U. S. Atomic Energy Commission UNIVERSITY OF CALIFORNIA Radiation Laboratory Berkeley, California Contract No. W - 7405 - eng – 48. https://www.google.com/books/edition/A_Special_Consideration_of_the_Aging_Pro/xLL4pko-4hoC?hl=en

7 Torres-Ibarra L, Basto-Abreu A, Carnalla M, Torres-Alvarez R, Reyes-Sanchez F, Hernández-Ávila JE, Palacio-Mejia LS, Alpuche-Aranda C, Shamah-Levy T, Rivera JA, Barrientos-Gutierrez T. SARS-CoV-2 infection fatality rate after the first epidemic wave in Mexico. Int J Epidemiol. 2022 May 9;51(2):429–439. doi: 10.1093/ije/dyac015. PMID: 35157072; PMCID: PMC8903396.

8 Rawshani A, Kjölhede EA, Rawshani A, Sattar N, Eeg-Olofsson K, Adiels M, Ludvigsson J, Lindh M, Gisslén M, Hagberg E, Lappas G, Eliasson B, Rosengren A. Severe COVID-19 in people with type 1 and type 2 diabetes in Sweden: A nationwide retrospective cohort study. Lancet Reg Health Eur. 2021 May;4:100105. doi: 10.1016/j.lanepe.2021.100105. Epub 2021 Apr 30. PMID: 33969336; PMCID: PMC8086507.

9 CDC. Rates of COVID-19 Cases or Deaths by Age Group and Vaccination Status and Booster Dose. https://covid.cdc.gov/covid-data-tracker/#rates-by-vaccine-status Updated 9/20/22 https://data.cdc.gov/Public-Health-Surveillance/Rates-of-COVID-19-Cases-or-Deaths-by-Age-Group-and/d6p8-wqjm

10 Saban M, Myers V, Wilf-Miron R. Changes in infectivity, severity and vaccine effectiveness against delta COVID-19 variant ten months into the vaccination program: The Israeli case. Prev Med. 2022 Jan;154:106890. doi: 10.1016/j.ypmed.2021.106890. Epub 2021 Nov 17. PMID: 34800471; PMCID: PMC8596646.

11 Dickerman BA, Gerlovin H, Madenci AL, Figueroa Muñiz MJ, Wise JK, Adhikari N, Ferolito BR, Kurgansky KE, Gagnon DR, Cho K, Casas JP, Hernán MA. Comparative effectiveness of third doses of mRNA-based COVID-19 vaccines in US veterans. Nat Microbiol. 2023 Jan 2. doi: 10.1038/s41564-022-01272-z. Epub ahead of print. PMID: 36593297.

12 Rates of COVID-19 Cases or Deaths by Age Group and Updated (Bivalent) Booster Status https://data.cdc.gov/Public-Health-Surveillance/Rates-of-COVID-19-Cases-or-Deaths-by-Age-Group-and/54ys-qyzm

13 Vokó Z, Kiss Z, Surján G, Surján O, Barcza Z, Pályi B, Formanek-Balku E, Molnár GA, Herczeg R, Gyenesei A, Miseta A, Kollár L, Wittmann I, Müller C, Kásler M. Nationwide effectiveness of five SARS-CoV-2 vaccines in Hungary-the HUN-VE study. Clin Microbiol Infect. 2022 Mar;28(3):398–404. doi: 10.1016/j.cmi.2021.11.011. Epub 2021 Nov 25. PMID: 34838783; PMCID: PMC8612758.

14 CDC COVID Data Tracker: Rates of COVID-19 Cases and Deaths by Vaccination Status https://covid.cdc.gov/covid-data-tracker/#rates-by-vaccine-status https://data.cdc.gov/Public-Health-Surveillance/Rates-of-COVID-19-Cases-or-Deaths-by-Age-Group-and/ukww-au2k

15 Nyberg T, Ferguson NM, Nash SG, Webster HH, Flaxman S, Andrews N, Hinsley W, Bernal JL, Kall M, Bhatt S, Blomquist P, Zaidi A, Volz E, Aziz NA, Harman K, Funk S, Abbott S; COVID-19 Genomics UK (COG-UK) consortium, Hope R, Charlett A, Chand M, Ghani AC, Seaman SR, Dabrera G, De Angelis D, Presanis AM, Thelwall S. Comparative analysis of the risks of hospitalisation and death associated with SARS-CoV-2 omicron (B.1.1.529) and delta (B.1.617.2) variants in England: a cohort study. Lancet. 2022 Apr 2;399(10332):1303–1312. doi: 10.1016/S0140-6736(22)00462-7. Epub 2022 Mar 16. PMID: 35305296; PMCID: PMC8926413

16 Skarbinski J, Wood MS, Chervo TC, Schapiro JM, Elkin EP, Valice E, Amsden LB, Hsiao C, Quesenberry C, Corley DA, Kushi LH. Risk of severe clinical outcomes among persons with SARS-CoV-2 infection with differing levels of vaccination during widespread Omicron (B.1.1.529) and Delta (B.1.617.2) variant circulation in Northern California: A retrospective cohort study. Lancet Reg Health Am. 2022 Aug;12:100297. doi: 10.1016/j.lana.2022.100297. Epub 2022 Jun 16. PMID: 35756977; PMCID: PMC9212563.

17 Global Burden of Disease Long COVID Collaborators, Wulf Hanson S, Abbafati C, Aerts JG, Al-Aly Z, Ashbaugh C, Ballouz T, Blyuss O, Bobkova P, Bonsel G, Borzakova S, Buonsenso D, Butnaru D, Carter A, Chu H, De Rose C, Diab MM, Ekbom E, El Tantawi M, Fomin V, Frithiof R, Gamirova A, Glybochko PV, Haagsma JA, Haghjooy Javanmard S, Hamilton EB, Harris G, Heijenbrok-Kal MH, Helbok R, Hellemons ME, Hillus D, Huijts SM, Hultström M, Jassat W, Kurth F, Larsson IM, Lipcsey M, Liu C, Loflin CD, Malinovschi A, Mao W, Mazankova L, McCulloch D, Menges D, Mohammadifard N, Munblit D, Nekliudov NA, Ogbuoji O, Osmanov IM, Peñalvo JL, Petersen MS, Puhan MA, Rahman M, Rass V, Reinig N, Ribbers GM, Ricchiuto A, Rubertsson S, Samitova E, Sarrafzadegan N, Shikhaleva A, Simpson KE, Sinatti D, Soriano JB, Spiridonova E, Steinbeis F, Svistunov AA, Valentini P, van de Water BJ, van den Berg-Emons R, Wallin E, Witzenrath M, Wu Y, Xu H, Zoller T, Adolph C, Albright J, Amlag JO, Aravkin AY, Bang-Jensen BL, Bisignano C, Castellano R, Castro E, Chakrabarti S, Collins JK, Dai X, Daoud F, Dapper C, Deen A, Duncan BB, Erickson M, Ewald SB, Ferrari AJ, Flaxman AD, Fullman N, Gamkrelidze A, Giles JR, Guo G, Hay SI, He J, Helak M, Hulland EN, Kereselidze M, Krohn KJ, Lazzar-Atwood A, Lindstrom A, Lozano R, Malta DC, Månsson J, Mantilla Herrera AM, Mokdad AH, Monasta L, Nomura S, Pasovic M, Pigott DM, Reiner RC Jr, Reinke G, Ribeiro ALP, Santomauro DF, Sholokhov A, Spurlock EE, Walcott R, Walker A, Wiysonge CS, Zheng P, Bettger JP, Murray CJL, Vos T. Estimated Global Proportions of Individuals With Persistent Fatigue, Cognitive, and Respiratory Symptom Clusters Following Symptomatic COVID-19 in 2020 and 2021. JAMA. 2022 Oct 25;328(16):1604–1615. doi: 10.1001/jama.2022.18931. PMID: 36215063; PMCID: PMC9552043..

18 Al-Aly Z, Bowe B, Xie Y. Long COVID after breakthrough SARS-CoV-2 infection. Nat Med. 2022 Jul;28(7):1461–1467. doi: 10.1038/s41591-022-01840-0. Epub 2022 May 25. PMID: 35614233; PMCID: PMC9307472..

19 https://data.gov.scot/coronavirus-covid-19/detail.html#excess_deaths

20 Paglino E, Lundberg DJ, Cho A, Wasserman JA, Raquib R, Luck AN, Hempstead K, Bor J, Elo IT, Preston SH, Stokes AC. Excess all-cause mortality across counties in the United States, March 2020 to December 2021. medRxiv [Preprint]. 2022 May 17:2022.04.23.22274192. doi: 10.1101/2022.04.23.22274192. PMID: 35547848; PMCID: PMC9094106. https://www.ncbi.nlm.nih.gov/pmc/articles/PMC9094106/

21 Excess Deaths Associated with COVID-19 https://www.cdc.gov/nchs/nvss/vsrr/covid19/excess_deaths.htm

22 Excess mortality in England analysis https://app.powerbi.com/view?r=eyJrIjoiYmUwNmFhMjYtNGZhYS00NDk2LWFlMTAtOTg0OGNhNmFiNGM0IiwidCI6ImVlNGUxNDk5LTRhMzUtNGIyZS1hZDQ3LTVmM2NmOWRlODY2NiIsImMiOjh9

23 Emecen AN, Keskin S, Turunc O, Suner AF, Siyve N, Basoglu Sensoy E, Dinc F, Kilinc O, Avkan Oguz V, Bayrak S, Unal B. The presence of symptoms within 6 months after COVID-19: a single-center longitudinal study. Ir J Med Sci. 2022 Jun 17:1–10. doi: 10.1007/s11845-022-03072-0. Epub ahead of print. PMID: 35715663; PMCID: PMC9205653.

24 Subramanian A, Nirantharakumar K, Hughes S, Myles P, Williams T, Gokhale KM, Taverner T, Chandan JS, Brown K, Simms-Williams N, Shah AD, Singh M, Kidy F, Okoth K, Hotham R, Bashir N, Cockburn N, Lee SI, Turner GM, Gkoutos GV, Aiyegbusi OL, McMullan C, Denniston AK, Sapey E, Lord JM, Wraith DC, Leggett E, Iles C, Marshall T, Price MJ, Marwaha S, Davies EH, Jackson LJ, Matthews KL, Camaradou J, Calvert M, Haroon S. Symptoms and risk factors for long COVID in non-hospitalized adults. Nat Med. 2022 Aug;28(8):1706–1714. doi: 10.1038/s41591-022-01909-w. Epub 2022 Jul 25. PMID: 35879616; PMCID: PMC9388369.

25 Gao P, Liu J, Liu M. Effect of COVID-19 Vaccines on Reducing the Risk of Long COVID in the Real World: A Systematic Review and Meta-Analysis. Int J Environ Res Public Health. 2022 Sep 29;19(19):12422. doi: 10.3390/ijerph191912422. PMID: 36231717; PMCID: PMC9566528.

26 Kuodi P, Gorelik Y, Zayyad H, Wertheim O, Wiegler KB, Abu Jabal K, Dror AA, Nazzal S, Glikman D, Edelstein M. Association between BNT162b2 vaccination and reported incidence of post-COVID-19 symptoms: cross-sectional study 2020-21, Israel. NPJ Vaccines. 2022 Aug 26;7(1):101. doi: 10.1038/s41541-022-00526-5. PMID: 36028498; PMCID: PMC9411827.

27 Ayoubkhani D, Bosworth ML, King S, Pouwels KB, Glickman M, Nafilyan V, Zaccardi F, Khunti K, Alwan NA, Walker AS. Risk of Long COVID in People Infected With Severe Acute Respiratory Syndrome Coronavirus 2 After 2 Doses of a Coronavirus Disease 2019 Vaccine: Community-Based, Matched Cohort Study. Open Forum Infect Dis. 2022 Sep 12;9(9):ofac464. doi: 10.1093/ofid/ofac464. PMID: 36168555; PMCID: PMC9494414.

28 Brannock MD, Chew RF, Preiss AJ, Hadley EC, McMurry JA, Leese PJ, Girvin AT, Crosskey M, Zhou AG, Moffitt RA, Funk MJ, Pfaff ER, Haendel MA, Chute CG; N3C and RECOVER Consortia. Long COVID Risk and Pre-COVID Vaccination: An EHR-Based Cohort Study from the RECOVER Program. medRxiv [Preprint]. 2022 Oct 7:2022.10.06.22280795. doi: 10.1101/2022.10.06.22280795. PMID: 36238713; PMCID: PMC9558440.

29 Azzolini E, Levi R, Sarti R, Pozzi C, Mollura M, Mantovani A, Rescigno M. Association Between BNT162b2 Vaccination and Long COVID After Infections Not Requiring Hospitalization in Health Care Workers. JAMA. 2022 Aug 16;328(7):676–678. doi: 10.1001/jama.2022.11691. PMID: 35796131; PMCID: PMC9250078.

30 Hastie CE, Lowe DJ, McAuley A, Winter AJ, Mills NL, Black C, Scott JT, O’Donnell CA, Blane DN, Browne S, Ibbotson TR, Pell JP. Outcomes among confirmed cases and a matched comparison group in the Long-COVID in Scotland study. Nat Commun. 2022 Oct 12;13(1):5663. doi: 10.1038/s41467-022-33415-5. Erratum in: Nat Commun. 2022 Nov 1;13(1):6540. PMID: 36224173; PMCID: PMC9556711.

31 Al-Aly Z, Bowe B, Xie Y. Long COVID after breakthrough SARS-CoV-2 infection. Nat Med. 2022 Jul;28(7):1461–1467. doi: 10.1038/s41591-022-01840-0. Epub 2022 May 25. PMID: 35614233; PMCID: PMC9307472.

32 Wherry EJ, Barouch DH. T cell immunity to COVID-19 vaccines. Science. 2022 Aug 19;377(6608):821–822. doi: 10.1126/science.add2897. Epub 2022 Aug 18. PMID: 35981045.

33 Michaelson J. Cell selection in development. Biol Rev Camb Philos Soc. 1987 May;62(2):115–39. doi: 10.1111/j.1469-185x.1987.tb01264.x. PMID: 3300794.

34 Michaelson J. Cellular selection in the genesis of multicellular organization. Lab Invest. 1993 Aug;69(2):136–51. PMID: 8350596.

35 Michaelson J. The role of molecular discreteness in normal and cancerous growth. Anticancer Res. 1999 Nov-Dec;19(6A):4853–67. PMID: 10697599.

36 Cagan A, Baez-Ortega A, Brzozowska N, Abascal F, Coorens THH, Sanders MA, Lawson ARJ, Harvey LMR, Bhosle S, Jones D, Alcantara RE, Butler TM, Hooks Y, Roberts K, Anderson E, Lunn S, Flach E, Spiro S, Januszczak I, Wrigglesworth E, Jenkins H, Dallas T, Masters N, Perkins MW, Deaville R, Druce M, Bogeska R, Milsom MD, Neumann B, Gorman F, Constantino-Casas F, Peachey L, Bochynska D, Smith ESJ, Gerstung M, Campbell PJ, Murchison EP, Stratton MR, Martincorena I. Somatic mutation rates scale with lifespan across mammals. Nature. 2022 Apr;604(7906):517–524. doi: 10.1038/s41586-022-04618-z. Epub 2022 Apr 13. PMID: 35418684; PMCID: PMC9021023.

37 Gladyshev VN et al. Molecular damage in aging. Nature Aging. volume 1, pages 1096–1106 (2021)

38 Gladyshev VN. On the cause of aging and control of lifespan: heterogeneity leads to inevitable damage accumulation, causing aging; control of damage composition and rate of accumulation define lifespan. Bioessays. 2012 Nov;34(11):925–9. doi: 10.1002/bies.201200092. Epub 2012 Aug 23. PMID: 22915358; PMCID: PMC3804916.

39 Johnson, J. Five things about covid we still don’t understand at our peril. The Washington Post September 26, 2022

