## APPENDIX for "Counting Cases and Deaths by Age Tells Us About COVID-19’s Infectious and Lethal Components"

### FOR

James Michaelson PhD

Department of Pathology, Massachusetts General Hospital, USA
Department of Surgery, Massachusetts General Hospital, USA
Department of Pathology, Harvard Medical School, USA

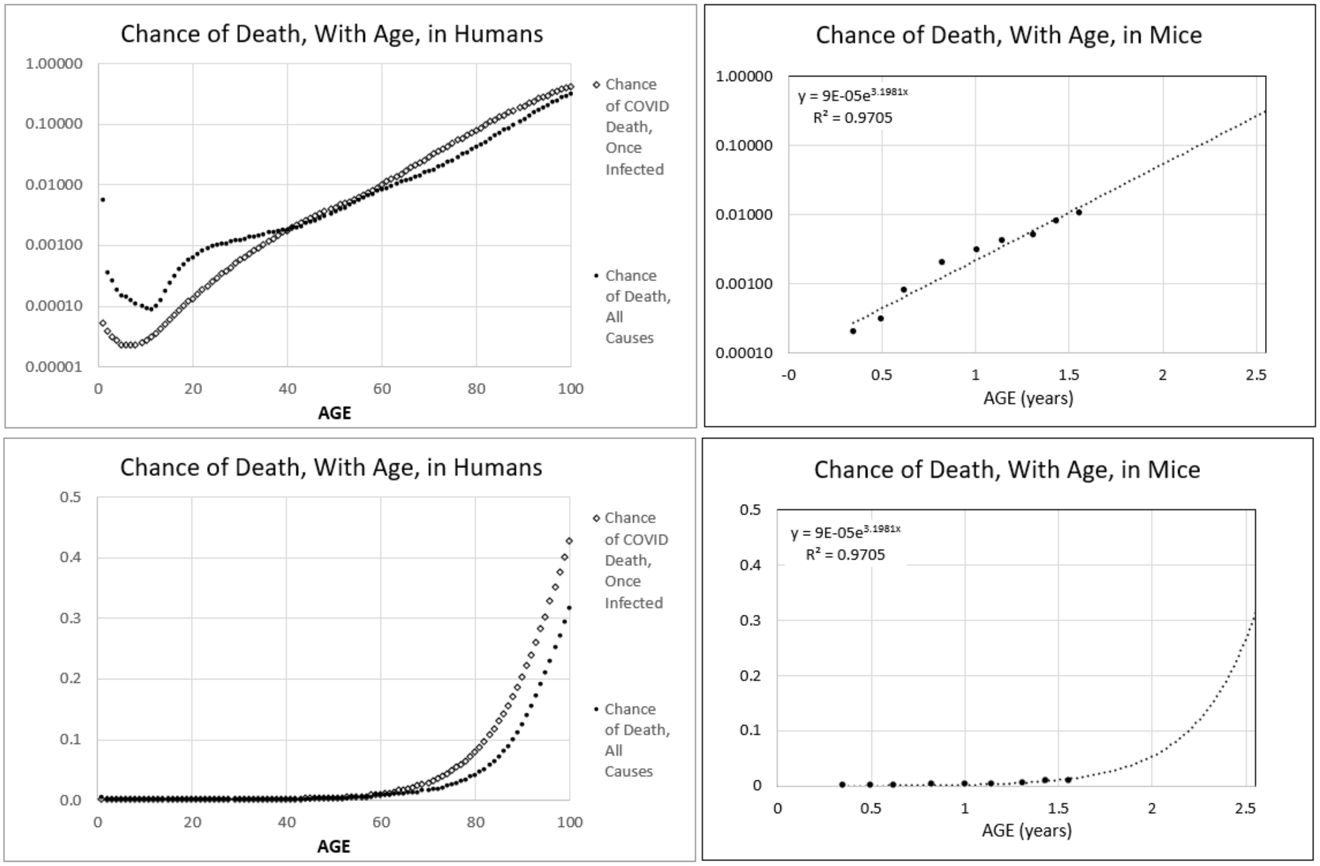


APPENDIX-FIGURE **1:** The ***Gompertzian Force of Mortality*** captured by the ***Gompertz Mortality Equation***.

#### ***Gompertzian Analysis*** of COVID-19 in the Pre-Vaccination Era

Levin *et al*'s initial report of the log-linear, ***Gompertzian***, appearance of the age association of COVID-19 lethality[1] was subsequently confirmed by a number of additional studies summarizing many countries (APPENDIX-FIGURE 3, values kindly provided by Dr. Brazeau[2,3,4,5,,]). Levin followed up with a comparative analysis of the age distribution of COVID-19 lethality in the developed and undeveloped world[6], revealing a somewhat higher occurrence of death by age, thus characterizing the ~10,000-fold exponential difference in COVID-19 lethality from infancy to old age (APPENDIX-FIGURE 2, APPENDIX-FIGURE 3).

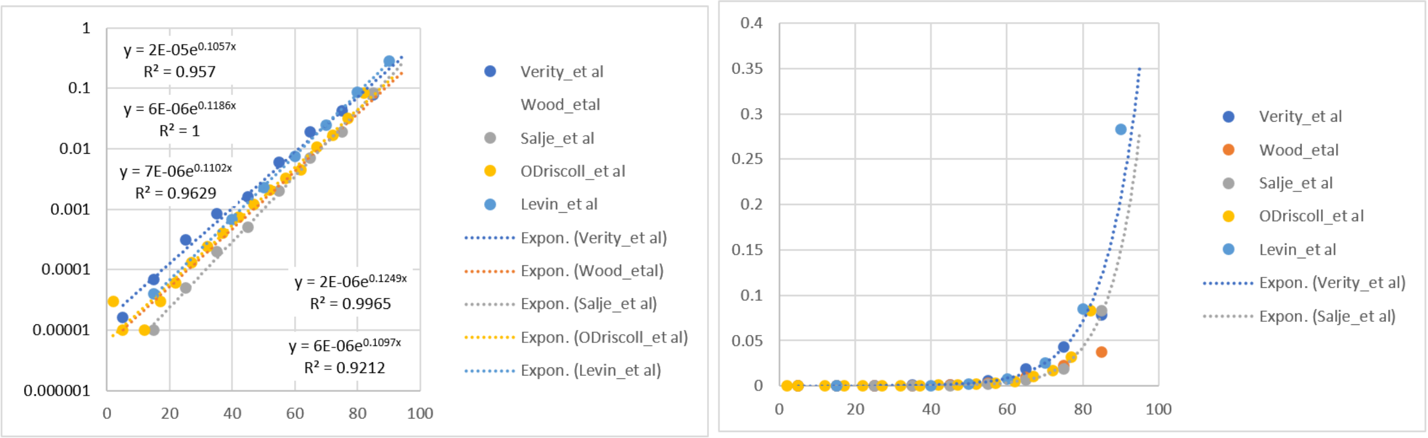


APPENDIX-FIGURE 2

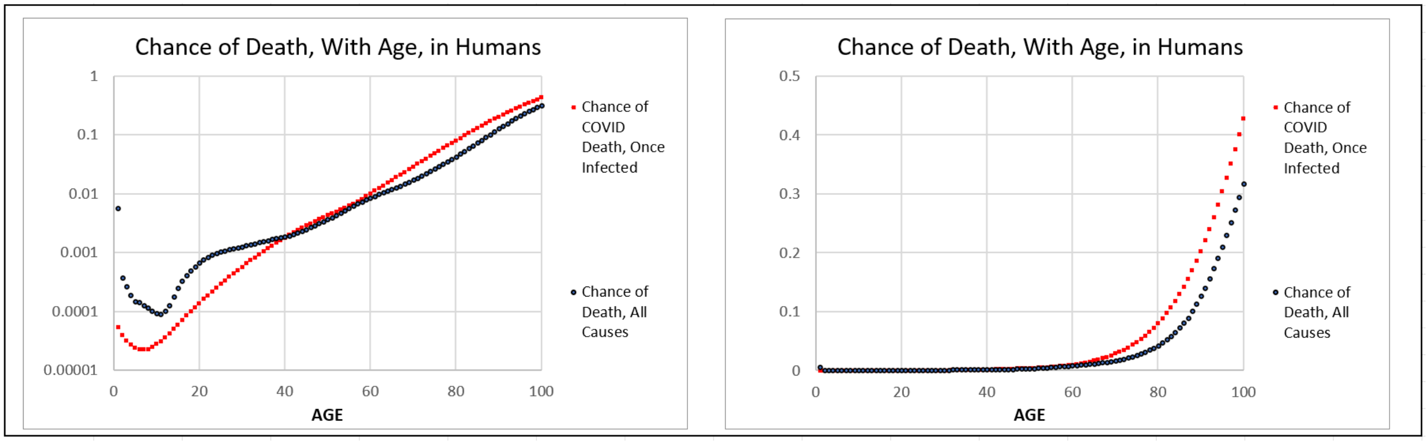


APPENDIX-FIGURE 3[7]

#### COVID's Fearful ***Gompertzian Force of Mortality*** Comes Into View When Displayed Linearly

As we shall see throughout this report, ***Gompertz Plots***, that is, the log plots of age-sorted data, and their mathematical equivalent, the exponential equation, give us powerful insights into COVID-19 ***infectivity*** and ***lethality***, but they do not provide easy visualizations of the things we care about most: how many of us will be affected, what are our individual chances of being affected, and what can be done about it. These more personal and practical understandings come into view by transforming the log ***Gompertz Plots*** into linear graphs (APPENDIX-FIGURE 3, right).

The first thing that strikes us by such a linear revisualization is the shocking, progressively more lethal, outcome of COVID-19 that occurs as we age. For example, on such linear graphs as in APPENDIX-FIGURE 3 (right), we can hardly detect the line of the COVID-19 ***Gompertz Mortality Equation*** (Deaths/Cases) in the first five decades of life, when its ***Gompertzian Force of Mortality*** stealthily accumulates less than 1% chance of death, but from age 60 on, we see the progressively more catastrophic, exponential, increase in COVID-19 lethality, with risk of death rising from about 1% at age 60, to about 3% at age 70, to almost 10% at age 80, to almost 20% at age 90, and to almost 50% at age 100.

***Gompertzian Analysis*** of Other Infectious Diseases

A number of other infectious disease, but by no means all, display the log-linear ***Gompertzian*** case fatality relationship noted above for COVID-19.[8] ***Gompertz Plots*** of COVID-19, Hepatitis, and Influenza can be seen in APPENDIX-FIGURE 4.

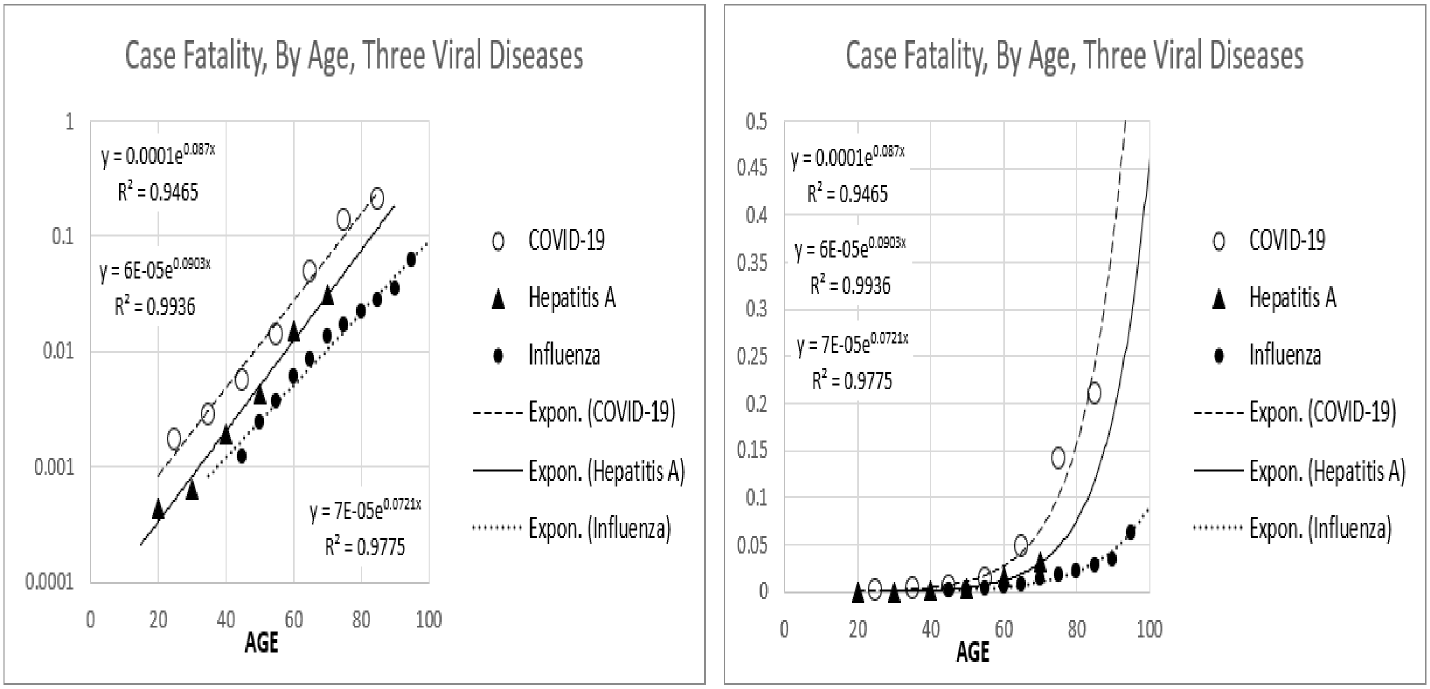


**APPENDIX-FIGURE 4**[8]

***Gompertzian Analysis*** of Non-Infectious Diseases

Many individual causes of death, not just infectious disease, have also been found to form ***Gompertz Lines***, as can be seen in APPENDIX-FIGURE 5 of data assembled by Jones[9]. Thus, the log-linear ***Gompertzian*** appearance of COVID-19 death is not a specific phenomenon of that disease, but a general phenomenon of aging, and most of the maladies of longevity. This wide spectrum of illnesses, all displaying the ***Gompertzian Force of Mortality***, may well reflect the body-wide decline in functional cells caused by the slowing of growth, and thus the decline in the fraction of cells dividing that marks adult life, as shown in the accompanying paper.[10]

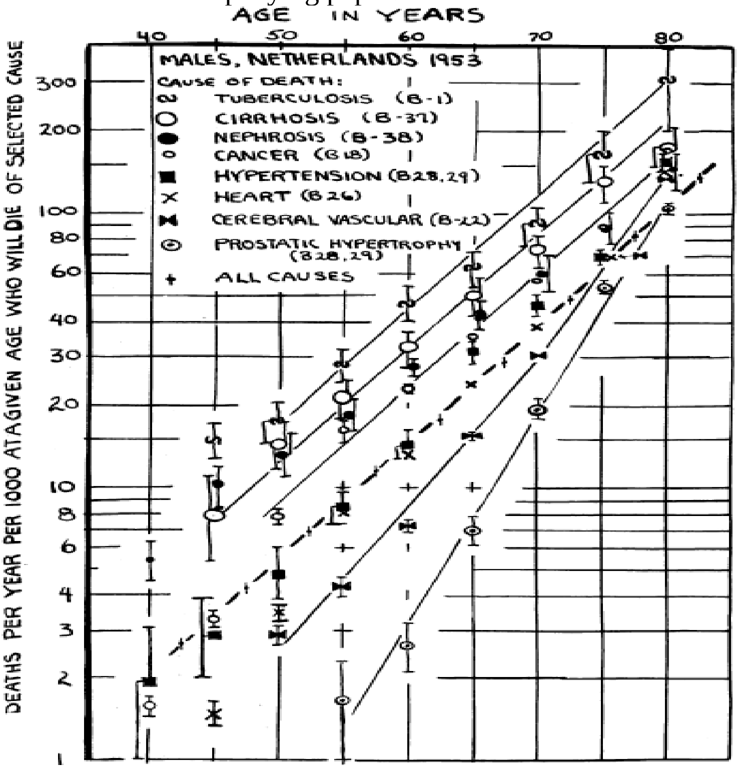


**APPENDIX-FIGURE 5**[9]

***Gompertzian Analysis*** of Young and Old.

Seldom do we have data from people of all ages. However, the exponential quality of the ***Gompertz Line*** allows us to make estimates of lethality over all ages from data on patients of just a few ages, whether young or old. An example of this can be seen in data from the USA[11] shown in APPENDIX-FIGURE 6, where the exponential fits made from patients of age 21-60, and 61-89, and all ages from 21-89, yields similar ***Gompertz Lines***, with similar ***Gompertzian Slopes***, $G_s$, and ***Gompertzian Heights***, $G_H$ (APPENDIX-FIGURE 6). Note in APPENDIX-FIGURE 7, how using data points provided by the COVID-19 Forecasting Team[7] (APPENDIX-FIGURE 4), values from age 40 to 70, or from 80 to 100, yields quite similar ***Gompertz Lines***, over the full life range of life from age 40 to 100. This means that we needn't have a full age sample of a population to make a reasonable picture of COVID-19 lethality over the full adult age range of a population.

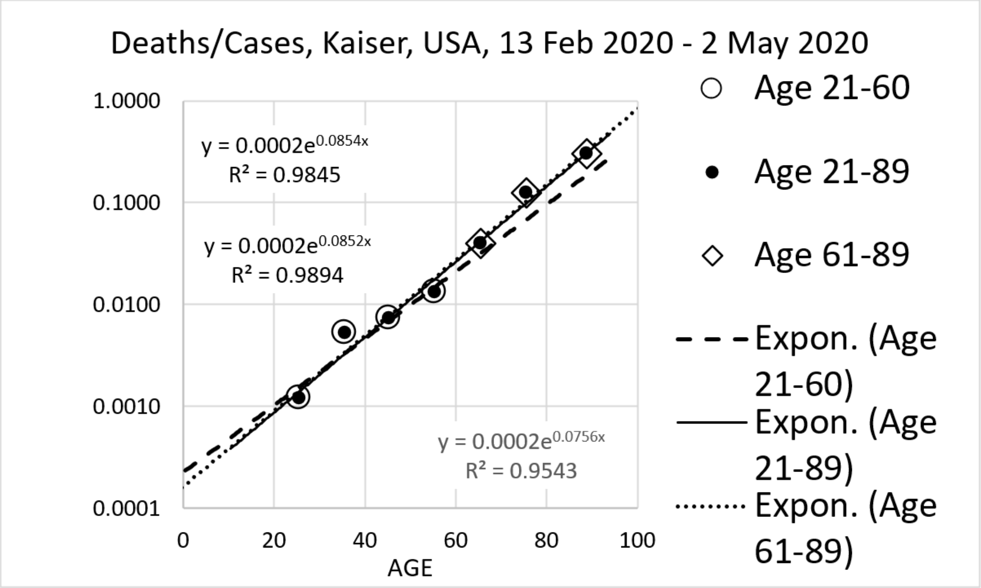


APPENDIX-FIGURE 6

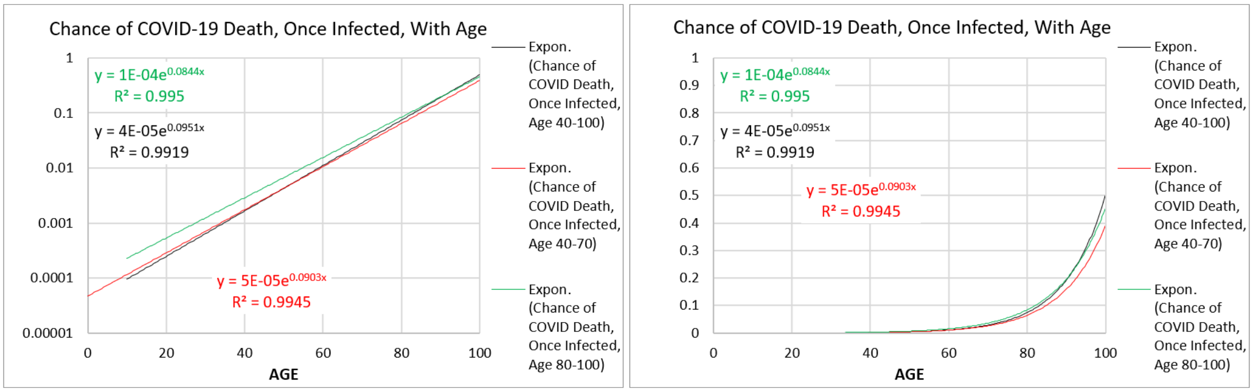


APPENDIX-FIGURE 7.[7]

***Gompertzian Analysis*** of COVID-19 ***infectivity*** and ***lethality***

USA(CDC), Analyzed in Aggregate

Basic Features, on Graphs.

Let us now examine the essential qualities of COVID-19 ***infectivity*** and ***lethality***, revealed by sorting data by age. In APPENDIX-FIGURE 8, are shown data from a rather extraordinary CDC dataset of approximately 70% of all COVID-19 cases in the USA from April 2021 to June 2022.[12] Here, I have sorted, by age, datapoints on unvaccinated patients from this dataset, and graphed them on a ***Gompertz Plot***, that is, a log plot, by age. The data sorted this way reveal three properties, which I have called:

***Pasteurian Infectivity***: the fraction of people in a population who have been infected (Cases/Number of People in the Population).
***Gompertzian Lethality***: the fraction of infected people who have died (Deaths/Cases).
***Malthusian Lethality***: the fraction of people in a population who have died (Deaths/Number of People in the Population).

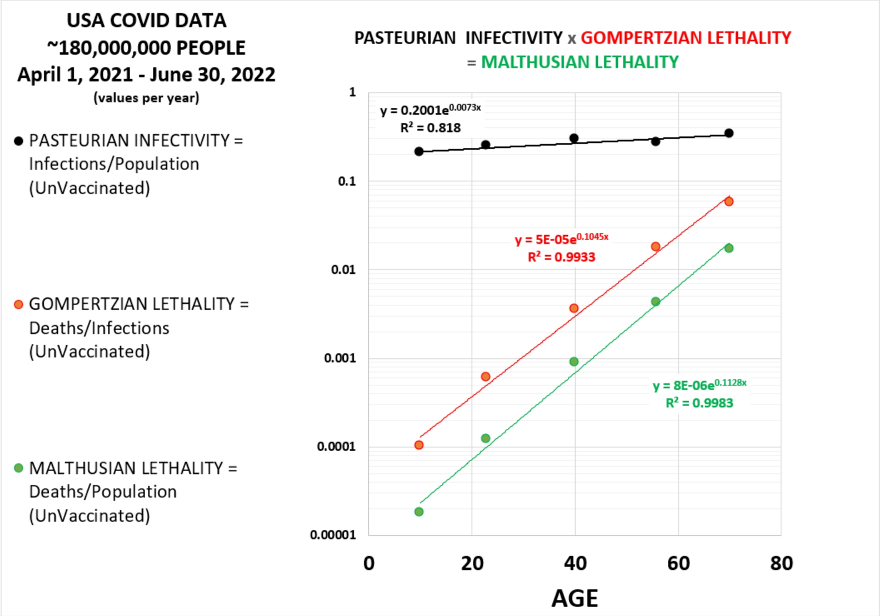


APPENDIX-FIGURE 8

Note, that both ***Gompertzian Lethality*** (Deaths/Cases), and ***Malthusian Lethality*** (Deaths/Population), display the alarming log-linear distribution in chance of death, in agreement with Levin's data, which revealed the ~10,000-fold, log-linear difference in COVID-19 lethality from infancy to old age (APPENDIX-FIGURE 8). On the other hand, the CDC data displays only a small, probably irrelevant, 2-fold, difference between the young and the old in ***Pasteurian Infectivity***, the fraction of people in the population who have been infected (Cases/Population). Thus, these data reveal that age has little, if any, practical impact in the chance of infection, but an enormous impact on the chance of death if infected.

Basic Features, in Numbers.

The associations of ***Pasteurian Infectivity***, ***Gompertzian Lethality***, and ***Malthusian Lethality*** with age could be quantified by fitting each set of points to the exponential equation (APPENDIX-FIGURE 8). For ***Gompertzian Lethality*** (Deaths/Cases) and ***Malthusian Lethality*** (Deaths/Population), these exponential regressions distilled each of these qualities down to just two numbers: the $G_{HG}$ (the ***Gompertzian Height*** of ***Gompertzian Lethality***), and $G_{SG}$ (the ***Gompertzian Slope*** of ***Gompertzian Lethality***), and the $G_{HM}$ (the ***Gompertzian Height*** of ***Malthusian Lethality***), and $G_{SM}$ (the ***Gompertzian Slope*** of ***Malthusian Lethality***).

For ***Pasteurian Infectivity***, these exponential regressions capture the small, probably irrelevant, 2-fold, difference between the youngest and oldest reflected in the $G_{SP}$ (the ***Gompertzian Slope*** of ***Pasteurian Infectivity***), being close to zero. Since any number to the zero power equals 1, as a useful approximation, the $G_{SP}$ can be ignored, with the measure of ***Pasteurian Infectivity*** reflected simply in the $G_{HP}$ (the ***Gompertzian Height*** of ***Pasteurian Infectivity***).

Numerically, the data presented here reveal, over and over, that the whatever might be the change in ***Gompertzian Height***, $G_H$, induced by vaccinations and other forces, $G_{SG}$ (the ***Gompertzian Slope*** of ***Gompertzian Lethality***) and $G_{SM}$ (the ***Gompertzian Slope*** of ***Malthusian Lethality***), both of which capture the ~10,000-fold rise in lethality with age, has repeatedly been found to have a value of **~0.1**. Similarly, since ***Pasteurian Infectivity*** tends to have similar values among patients of various ages, $G_{SP}$ (the ***Gompertzian Slope*** of ***Pasteurian Infectivity***), has repeatedly been found to have a value of **~1.0**.

Thus, $G_{HP}$ (the ***Gompertzian Height*** of ***Pasteurian Infectivity***), $G_{HG}$ (the ***Gompertzian Height*** of ***Gompertzian Lethality***), and $G_{HM}$ (the ***Gompertzian Height*** of ***Malthusian Lethality***), each provide a single measure of COVID-19 ***infectivity*** or ***lethality***. For all three qualities, higher is bad, lower is good.

**_Gompertzian Analysis_** <u>of COVID-19 Vaccination: USA(CDC), Analyzed in Aggregate</u>

<u>Basic Features, on Graphs.</u>

In APPENDIX-FIGURE 9, I have now added data on the people that had been vaccinated. Note how vaccination led to a reduction in ***Pasteurian Infectivity***, the fraction of people in a population who have been infected (Cases/Population), and to a reduction in ***Gompertzian Lethality***, the fraction of infected people who have died (Deaths/Cases). The aggregate effect is seen in a reduction of ***Malthusian Lethality***, the fraction of people in a population who have died (Deaths/Population).

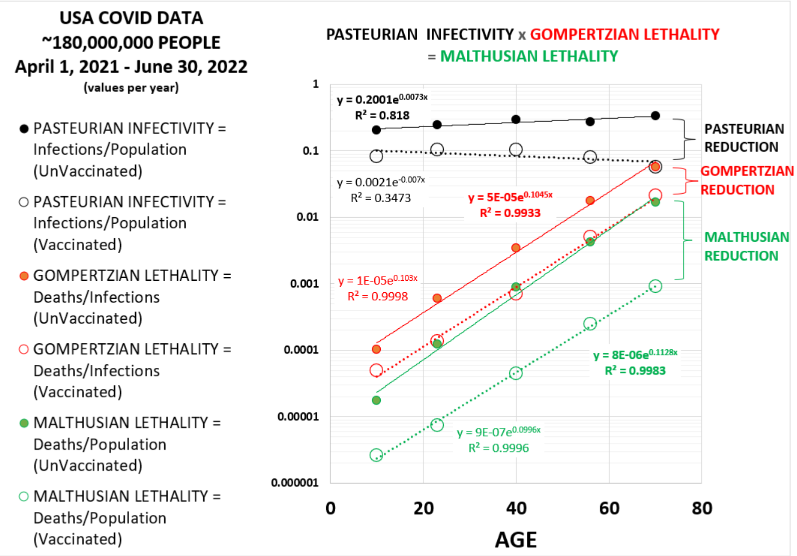


APPENDIX-FIGURE 9

<u>Basic Features, in Numbers</u>

There are several ways to frame these changes induced by vaccination, so as to address specific questions and needs.

<u>***Reductions***</u>

We can capture these measures of vaccine activity with the term I call a "***Reduction***," "1-(Vaccinated/Unvaccinated)", expressed as a percentage. Because the ***Pasteurian Infectivity***, (Cases/Population) occurs at similar rates across the ages, population-wide estimates are straightforward, and roughly in agreement with more common Vaccine Effectiveness measures.

In contrast, because the ***Gompertzian*** and ***Malthusian Lethality*** (Deaths/Cases; Deaths/ Population) differs ~10,000-fold from infancy to old age, aggregate measures, specifically Vaccine Effectiveness, are inherently prone to inaccuracy. However, because the relationship of ***Gompertzian Lethality*** to age fits the log-linear exponential ***Gompertz Mortality Equation***, with similar ***Gompertzian*** and ***Malthusian Slopes***, $\boldsymbol{G_{SG}}$, and $\boldsymbol{G_{SM}}$, that is to say, because the ***Gompertz Lines*** of the vaccinated and unvaccinated are roughly parallel, ***Gompertzian*** and ***Malthusian Reduction*** measures can be made by taking an average of the differences between vaccinated and unvaccinated in each of the various age groups.

Calculations with the data shown in APPENDIX-FIGURE 9 show that the ***Gompertzian Reduction***, that is the reduction in ***Gompertzian Lethality*** (Deaths/Cases) of vaccinated compared to unvaccinated individuals, averaged from all of the age groups, was 73%. The ***Pasteurian Reduction***, that is the reduction in ***Pasteurian Infectivity*** (Cases/Population) of vaccinated compared to unvaccinated individuals, averaged from all of the age groups, was 83%. The ***Malthusian Reduction***, that is the reduction in ***Malthusian Lethality*** (Deaths/Population) of vaccinated compared to unvaccinated individuals, averaged from all of the age groups, was 94%.

As noted above, these ***Reduction*** measures capture a vaccine's impact in manner that is similar, if not precisely the same, as the more familiar metric of Vaccine Effectiveness. I shall return to the similarities, and differences, of these measures in the main text. Perhaps the most striking feature is that ***Gompertzian Lethality*** (Deaths/Cases), and its ***Gompertzian Reduction***, provides a measure of the intrinsic lethality of COVID-19, independent of age, a measure that was not available before.

<u>***Fold Changes***</u>

Sometimes it can be useful to frame a vaccine's impact in terms of "***how many-fold***", as calculated simply by: (Vaccinated/Unvaccinated). For example, a vaccine that led to 1/3rd of the numbers of infections gave a ***3-fold*** reduction in ***Pasteurian Infectivity***. Thus, as shown in APPENDIX-FIGURE 9 the average ***Gompertzian Reduction*** of 73% caused by vaccination in the USA population reflects a ***4-fold*** reduction in in ***Gompertzian Lethality*** (Deaths/Cases). Similarly, the average ***Pasteurian Reduction*** was 83% reflects a ***6-fold*** reduction in in ***Pasteurian Infectivity*** (Cases/Population), and the average ***Malthusian Reduction*** 94% in this dataset reflects a ***17-fold*** reduction in in ***Malthusian Lethality*** (Deaths/Population).

<u>***Chances***</u>

One can also frame values for ***Gompertzian Lethality***, ***Pasteurian Infectivity***, and ***Malthusian Lethality***, and the impact which vaccination and other forces have on the measures of COVID-19 ***infectivity*** and ***lethality***, in terms of their ***chances***, often calculated simply as (Unvaccinated/Vaccinated).

Practical Features of Vaccination Impact

As we examined above, the ***Gompertz Plots***, that is, the log plots, give us insight into the forces of ***infectivity*** and ***lethality***, but when we transform them into linear graphs, we get the more practical, everyday, appreciations of how many of us will be affected and what our individual chances of being affected are (APPENDIX-FIGURE 10). Indeed, linear graphs vividly display how dramatically ***Gompertzian Lethality*** (Deaths/Cases) changes with age, and leads to its major burden among the elderly. Thus, we see easily on a linear graph of CDC data how individuals below age 60 still had less than 1% chance of death after infection, and thus vaccination leads to very small absolute ***Gompertzian Reductions*** among this group. However, from age 60 onward, theses absolute differences between vaccinated an unvaccinated, captured by their ***Gompertzian Reductions***, take on visually striking differences on linear graphs, with 20% of unvaccinated 80-years-olds in the CDC dataset dying after infection, in comparison to 5% of the vaccinated, and 60% of unvaccinated 90-years-olds dying after infection, in comparison to 15% of the vaccinated.

On the other hand, vaccine-driven ***Pasteurian Infectivity*** (Cases/Population) occurs at similar rates in individuals of all ages. Thus, ***Pasteurian Reductions*** in infection, are equally apparent on linear and log graphs.

***Gompertzian Lethality*** (Deaths/Cases) and ***Pasteurian Infectivity*** (Cases/Population) add up to deliver dramatic population-based ***Malthusian Lethality*** (Deaths/ Population), made most visible on linear graphs. Thus, ***Malthusian Reductions***, in this dataset display a 20-fold population-wide reductions in death by vaccination in all age groups, but the greatest absolute reduction in deaths become visually apparent beginning at age 70, with ever more striking savings in life in the 80's and 90's.

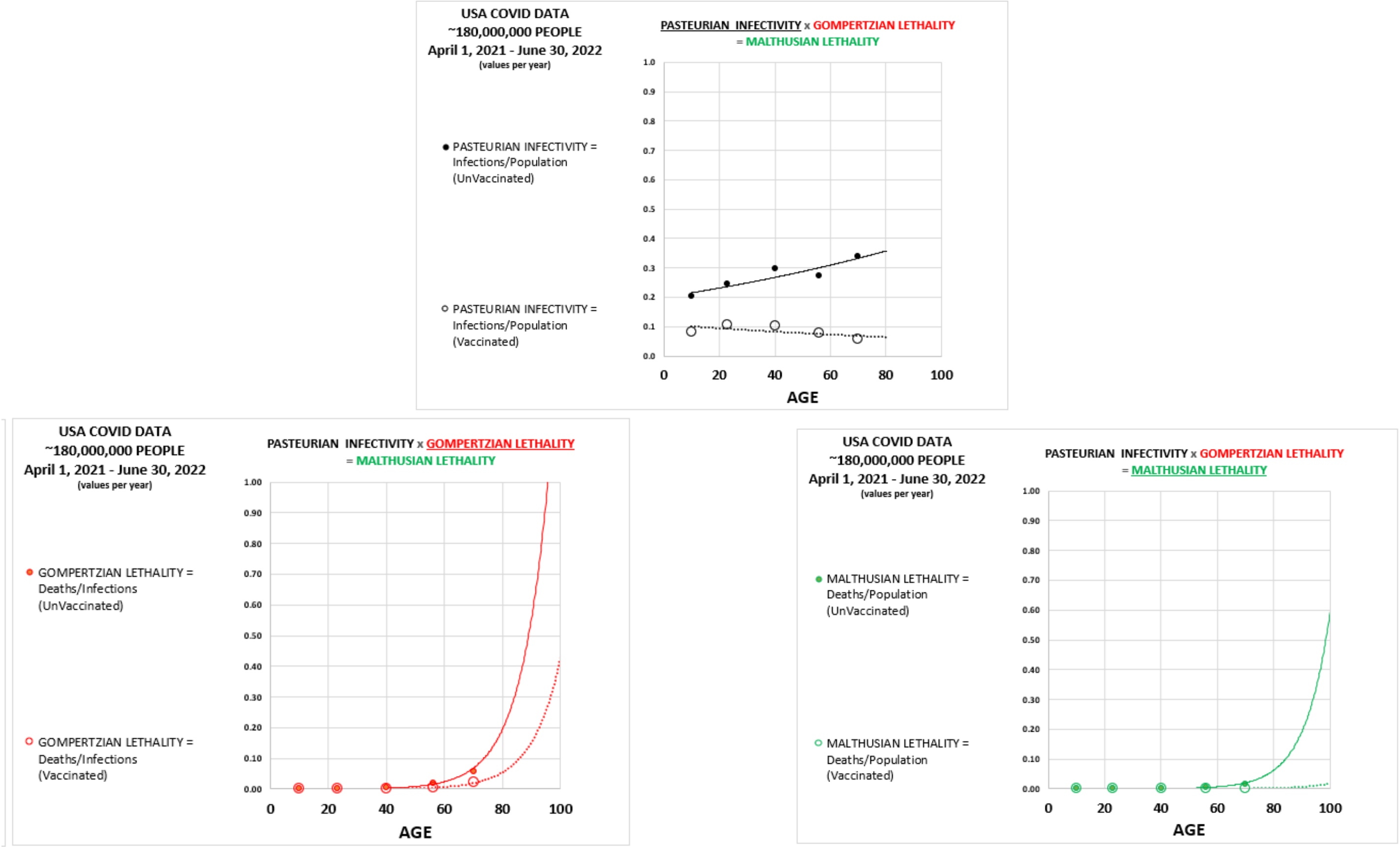


APPENDIX-FIGURE 10

With data on the age-structure of a population, one can now calculate the aggregate lethal and infectious impact across a population. From this, we can also estimate what would have been the impact of different usages of vaccination and other protective measures (not shown).

We shall return again to this marvelous dataset when we examine how the COVID-19 epidemic has evolved. However, first, let us take a look at several studies of smaller populations, which also give us insight in COVID-19 ***infectivity*** and ***lethality***.

***Gompertzian Analysis*** of COVID-19 Vaccination: USA(VA), Vaccine Impact

Two studies carried out at the US Veterans Affairs healthcare system, examined the impact of COVID-19 vaccination. In the first study, outcome for vaccinated individuals was compared with unvaccinated controls, December 11, 2020, to March 25, 2021, that is, about 3½ months, ending a few days before the CDC dataset reviewed above begins, which itself continued for the next 15 months, up until June 30, 2021;[13] the VA group then examined a follow-up to June 30, 2021.[14] Note in APPENDIX-FIGURE 11 how for this VA population, as saw earlier for the CDC dataset, that vaccination led to a reduction in ***Pasteurian Infectivity***, the fraction of people in a population who have been infected (Cases/Population), and in ***Gompertzian Lethality***, the fraction of infected people who have died (Deaths/Cases). The multiple, the aggregate reduction in the ***Malthusian Lethality***, the fraction of people in a population who have died (Deaths/Population), also displayed its reduction, as in the CDC population dataset.

The average ***Gompertzian Reduction***, that is the reduction in ***Gompertzian Lethality*** (Deaths/Cases) of vaccinated compared to unvaccinated individuals, of all of the age groups, was 53%. The average ***Pasteurian Reduction***, that is the reduction in ***Pasteurian Infectivity*** (Cases/Population) of vaccinated compared to unvaccinated individuals, of all of the age groups, was 69%. The average ***Malthusian Reduction***, that is the reduction in ***Malthusian Lethality*** (Deaths/Population) of vaccinated compared to unvaccinated individuals, of all of the age groups, was 87%. Note again how the ***Gompertz Lines*** of the vaccinated and unvaccinated are roughly parallel, indicating roughly equivalent reductions in all age groups.

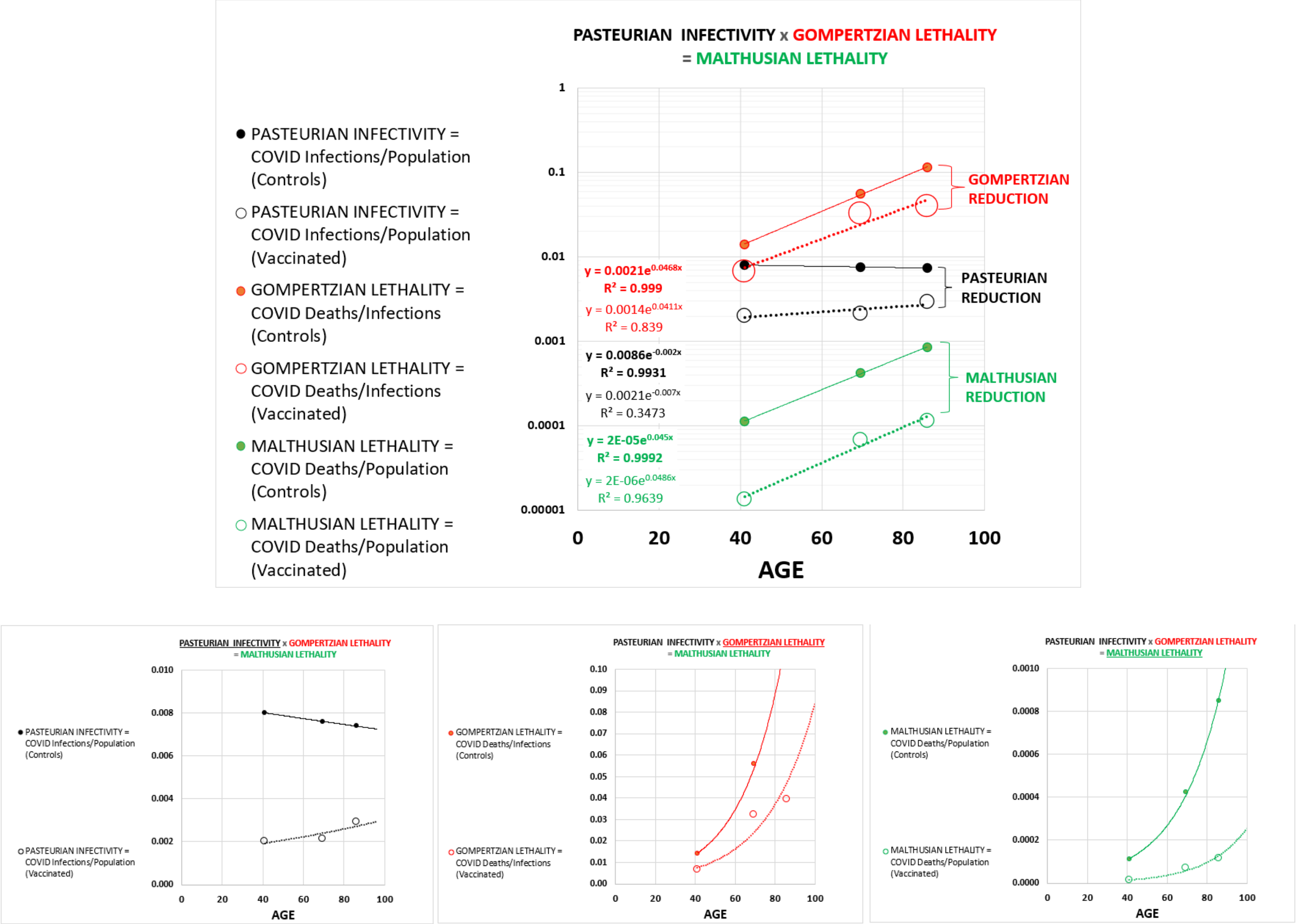


APPENDIX-FIGURE 11

***Gompertzian Analysis*** of COVID-19 Vaccination: USA(VA), Vaccine Comparison

In the second study of this population by this research group, during the same period, one vaccine was compared against another (Moderna Vs Pfizer) (APPENDIX-FIGURE 12). Both vaccines yielded quite similar ***Gompertz Lines*** of ***Gompertzian Lethality*** (Deaths/Cases), while the data on ***Pasteurian Infectivity***, the fraction of people in a population who have been infected (Cases/Population), appears to show a slightly better performance for the Moderna vaccine. The average ***Gompertzian Reduction***, of all of the age groups, that is the degree of lethality in ***Gompertzian Lethality*** (Deaths/Cases) of individuals given Moderna compared to Pfizer vaccines was 7%, essentially the same. On the other hand, the average ***Pasteurian Reduction***, of all of the age groups, that is the reduction in ***Pasteurian Infectivity*** (Cases/Population) of individuals given Moderna compared to Pfizer vaccines was 21%, with this modest reduction evident in each of the three age groups. The average resulting ***Malthusian Reduction***, of all of the age groups, that is the reduction in ***Malthusian Lethality*** (Deaths/Population) of individuals given Moderna compared to Pfizer vaccines was 12%.

Thus, the two vaccines examined by this study yielded almost the same ***Gompertzian Lethality*** (Deaths/Cases), but slightly different ***Pasteurian Infectivity***. This observation gives us our first hint, which we shall see appearing again and again below, that ***Gompertzian Lethality*** and ***Pasteurian Infectivity*** may well be separate, unlinked, outcomes of vaccination.

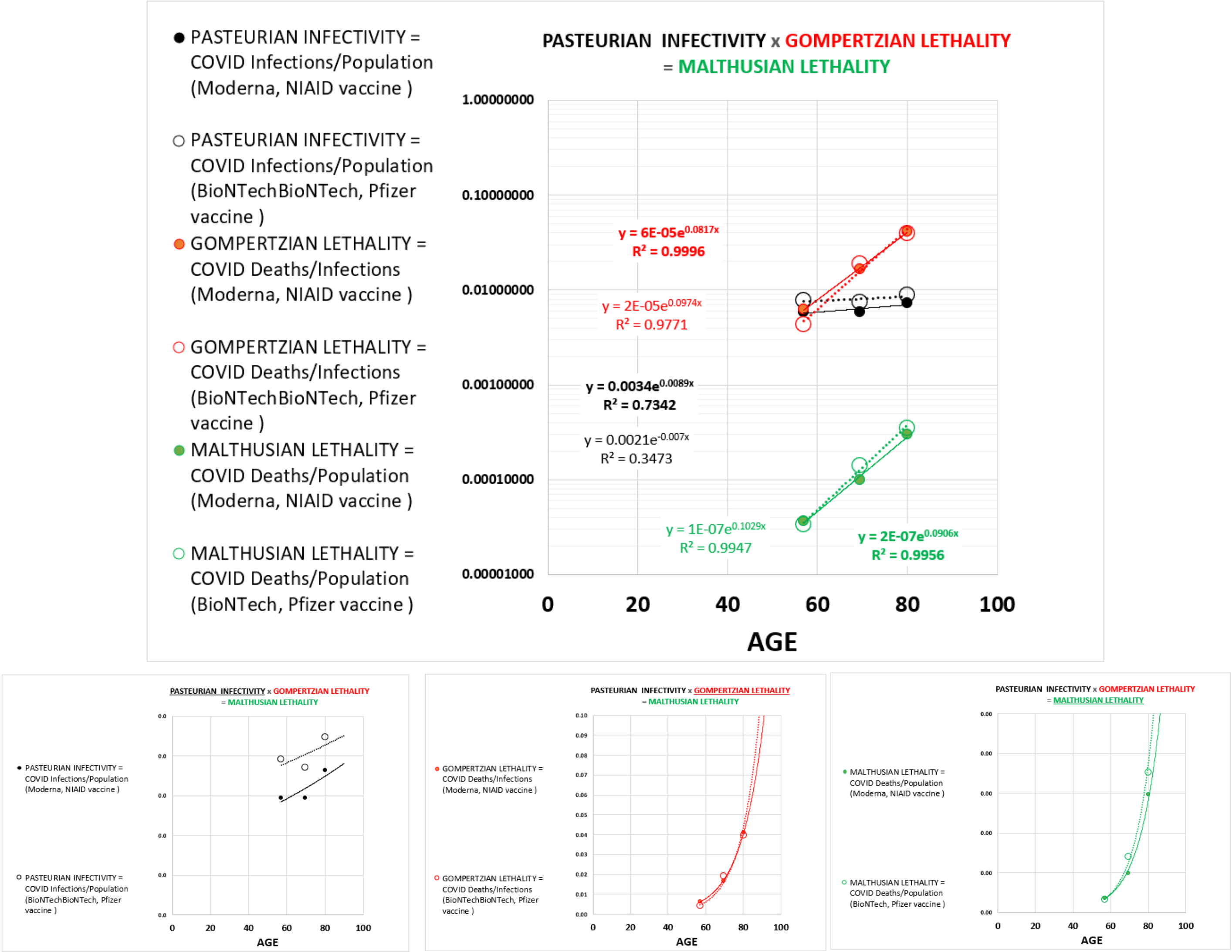


APPENDIX-FIGURE 12

***Gompertzian Analysis*** of COVID-19 Vaccination: HUNGARY and ARGENTINA, Vaccine Impact & Comparisons

The Hungarian national vaccination campaign, with 3.7 million vaccinated individuals, examined the impact of five vaccines (Pfizer, Moderna, Sputnik-V, AstraZeneca, Sinopharm), from January 22, 2021, to June 10, 2021, with data reported for finely grained age groups, for both the primary vaccine, and the two boosters, all described in three reports.[15,16,17] I also include in this section a second study, from Argentina, as it also measured the impact of the Sputnik-V vaccine.[18] The Sputnik-V vaccine has had a troubled history, with rightly criticized analysis, but the results from the Hungary and Argentina studies were so surprising to me, that I thought I should lay them out, despite my uneasiness with putting too much hope in the certainty of what the data might be telling us.

From the Hungary data, calculations of the average ***Gompertzian Reduction***, that is the reduction in ***Gompertzian Lethality*** (Deaths/Cases) of vaccinated compared to unvaccinated individuals of all of the age groups, was 56%. The average ***Pasteurian Reduction***, that is the reduction in ***Pasteurian Infectivity*** (Cases/Population) of vaccinated compared to unvaccinated individuals, of all of the age groups, was 87%. The average ***Malthusian Reduction***, that is the reduction in ***Malthusian Lethality*** (Deaths/Population) of vaccinated compared to unvaccinated individuals, of all of the age groups, was 93%. Note again how the ***Gompertz Lines*** of the vaccinated and unvaccinated are roughly parallel, indicating roughly equivalent reductions in all age groups (APPENDIX-FIGURE 13, APPENDIX-FIGURE 14).

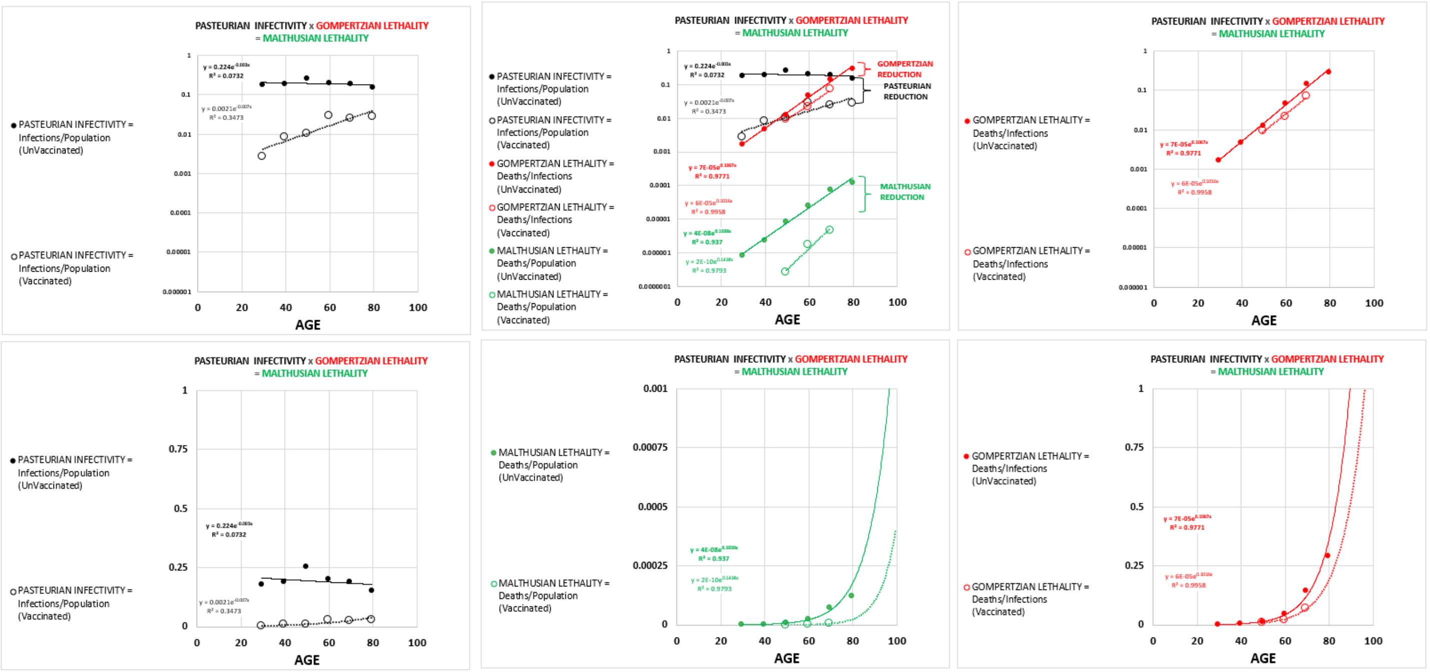


APPENDIX-FIGURE 13

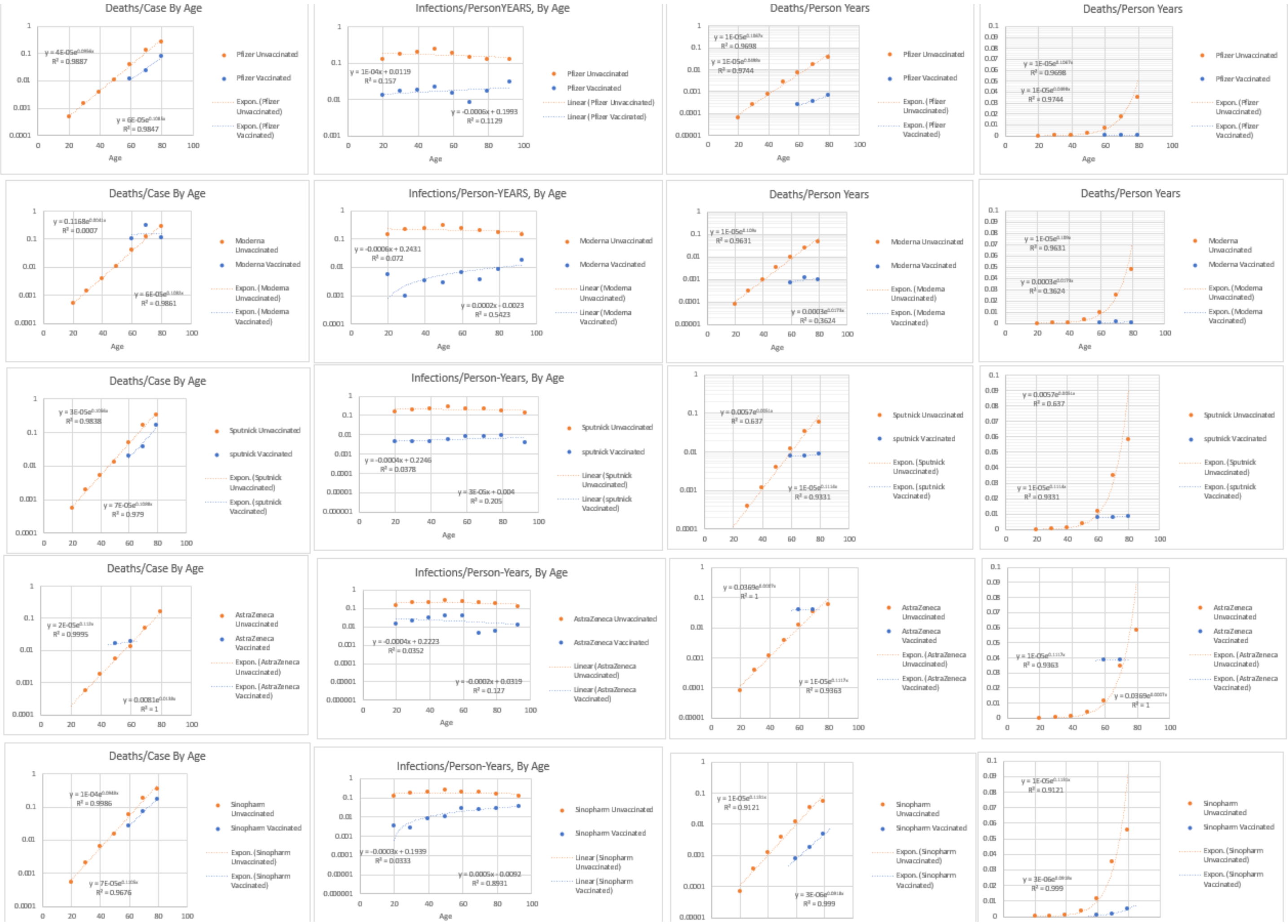


APPENDIX-FIGURE 14

***Gompertz Plots***, revealing log-linear ***Gompertz Lines***, of ***Gompertzian Lethality*** (Deaths/Cases) ***Pasteurian Infectivity*** (Cases/Population) and ***Malthusian Lethality*** (Deaths/Population) for the unvaccinated and vaccinated for each of the vaccines, can be seen in the figures above. Again, the ***Gompertz Lines*** of the vaccinated and unvaccinated are roughly parallel, indicating roughly equivalent reductions in all age groups.

Some of the age groups for individual vaccines had no deaths, which can't be visualized graphically, since the log of zero has no meaning, and thus has no place to go on a log graph. However, we can use the exponential equation, fit to the data from unvaccinated individuals, to calculate how many deaths would have occurred in each age group IF they had not been vaccinated. We can then add the numbers up, and see, over all ages, how much the reductions were. These calculations can be seen in the APPENDIX-TABLES I and II below.

Perhaps the most striking finding of these calculations was the remarkable benefits of the Sputnik vaccine, revealed by these data, especially on ***Gompertzian Lethality***. Thus, while the most widely used vaccine in Hungary, Pfizer, displayed a ***Gompertzian Reduction***, of 45%, a 1.81-fold reduction in the lethality, the Sputnik vaccine displayed a ***Gompertzian Reduction***, of 62%, a 2.63-fold reduction in the lethality. Pfizer displayed a ***Pasteurian Reduction***, of 90%, a 10.25-fold reduction in ***infectivity***, while the Sputnik vaccine displayed a ***Pasteurian Reduction***, of 99%, a 34.12-fold reduction in the ***infectivity***. This added up to Pfizer vaccine having caused a ***Malthusian Reduction***, of 95%, a 10.59-fold reduction in the deaths, while the Sputnik vaccine displayed a ***Malthusian Reduction***, of 99%, an 89.67-fold reduction in deaths.

| **Gompertzian Reduction** | Pfizer- | Moderna | Sputnik-V | AstraZeneca | Sinopharm | ALL |
|---|---|---|---|---|---|---|
| Observed CFR/Expected CFR-> | 0.55107 | 0.75426 | 0.3805 | 0.5857 | 0.56567 | 0.55573 |
| +/- 95% -> | 0.0185 | 0.0905 | 0.0423 | 0.0368 | 0.0205 | 0.0123 |
| Reduction, as a % (1-v/un) | 45% | 25% | 62% | 41% | 43% | 56% |
| **#-Fold Reduction in Death, once infected ->** | **1.81** | **1.33** | **2.63** | **1.71** | **1.77** | **1.80** |
| **Pasteurian Reduction** | | | | | | |
| Incidence Vacinated / Incidence UnVacinated -> | 0.0976 | 0.0308 | 0.02931 | 0.15854 | 0.13065 | 0.13065 |
| Reduction, as a % (1-v/un) | 90% | 97% | 97% | 84% | 87% | 87% |
| **#-Fold Reduction in Infection ->** | **10.25** | **32.46** | **34.12** | **6.31** | **7.65** | **7.65** |
| **Malthusian Reduction** (ie Combined) | 0.05378 | 0.02323 | 0.01115 | 0.09286 | 0.0739 | 0.0726 |
| Reduction, as a % (1-v/un) | 95% | 98% | 99% | 91% | 93% | 93% |
| **#-Fold Reduction in Death, Overall ->** | **18.59** | **43.04** | **89.67** | **10.77** | **13.53** | **13.77** |
| **Pasteurian Reduction / Gompertzian Reduction** | **2.01** | **3.94** | **1.57** | **2.03** | **2.00** | **1.56** |

**APPENDIX-TABLE I**

Not only did the Hungary data indicate striking differences between one vaccine and another in ***Gompertzian Lethality*** (Deaths/Cases) and ***Pasteurian Infectivity*** (Cases/Population), the two qualities appear to be independent manifestations of COVID-19 outcome, and COVID-19 vaccine impact, as we saw earlier in the USA(VA) vaccine comparison study outlined above, and will see in other instances below. This can be seen in the ratio of ***Pasteurian Reduction***/***Gompertzian Reduction***, in the comparison of one vaccine to another, varying from vaccine to vaccine more than two-fold.

The Hungary data revealing a remarkably superior outcome for the Sputnik vaccine, was also seen in a second study of this Sputnik vaccine, carried out in Argentina. With data from this Argentinian study, I could again calculate the value of ***Gompertzian Lethality***, that is, Deaths/Cases, by age. These calculations yielded a ***Gompertzian Reduction***, of 62%, a 2.64-fold reduction in the lethality, after the first dose, and a ***Gompertzian Reduction***, of 94%, a 15.41-fold reduction in the lethality, after the second dose.

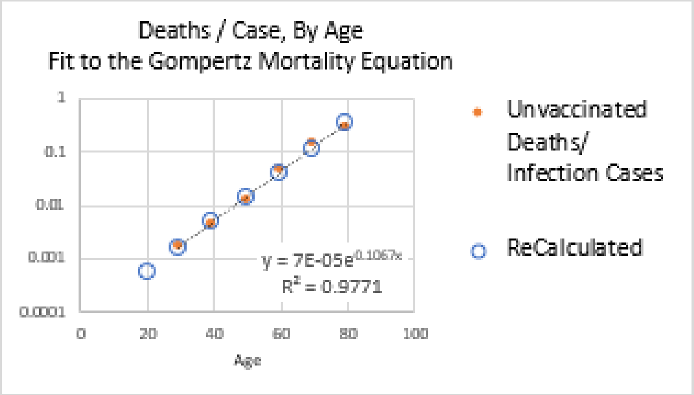


$$y = 7E\text{-}05e^{0.1067x}$$

| | Age | Age | | Pfizer- | Moderna | Sputnik-V | AstraZenec | Sinopharm | ALL |
|---|---|---|---|---|---|---|---|---|---|
| Cases, Among Vaccinated -> | 16–24 | 20 | Cases-> | 89 | 3 | 13 | 6 | 5 | 116 |
| Cases, Among Vaccinated -> | 25–34 | 29.5 | | 253 | 1 | 29 | 16 | 6 | 305 |
| Cases, Among Vaccinated -> | 35–44 | 39.5 | | 391 | 6 | 59 | 79 | 24 | 559 |
| Cases, Among Vaccinated -> | 45–54 | 49.5 | | 610 | 7 | 95 | 310 | 29 | 1,051 |
| Cases, Among Vaccinated -> | 55–64 | 59.5 | | 432 | 19 | 155 | 270 | 456 | 1,332 |
| Cases, Among Vaccinated -> | 65–74 | 69.5 | | 335 | 16 | 131 | 5 | 1,124 | 1,611 |
| Cases, Among Vaccinated -> | 75–84 | 79.5 | | 666 | 35 | 25 | 3 | 593 | 1,322 |
| Total Cases, Among Vaccinated -> | | | | 2,776 | 87 | 507 | 689 | 2,237 | 6,296 |
| | Age | Age | | Pfizer- | Moderna | Sputnik-V | AstraZenec | Sinopharm | ALL |
| Deaths, Among Vaccinated -> | 16–24 | 20 | Deaths -> | 0 | 0 | 0 | 0 | 0 | 0 |
| Deaths, Among Vaccinated -> | 25–34 | 29.5 | | 0 | 0 | 0 | 0 | 0 | 0 |
| Deaths, Among Vaccinated -> | 35–44 | 39.5 | | 0 | 0 | 0 | 0 | 0 | 0 |
| Deaths, Among Vaccinated -> | 45–54 | 49.5 | | 7 | 0 | 0 | 5 | 0 | 12 |
| Deaths, Among Vaccinated -> | 55–64 | 59.5 | | 10 | 2 | 3 | 5 | 12 | 32 |
| Deaths, Among Vaccinated -> | 65–74 | 69.5 | | 26 | 5 | 5 | 0 | 82 | 118 |
| Deaths, Among Vaccinated -> | 75–84 | 79.5 | | 118 | 4 | 4 | 0 | 104 | 230 |
| Total Deaths, Among Vaccinated -> | | | | 161 | 11 | 12 | 10 | 198 | 392 |
| | Age | Age | | Pfizer- | Moderna | Sputnik-V | AstraZenec | Sinopharm | ALL |
| | | | | Expected Cases, in matched indiduals, unvaccinated | | | | | |
| Incidence Vacinated / Incidence UnVacinated -> | | | | 10% | 3% | 3% | 16% | 13% | 13% |
| Expected Cases, if unvaccinated -> | 16–24 | 20 | | 912 | 97 | 444 | 38 | 38 | 1,529 |
| Expected Cases, if unvaccinated -> | 25–34 | 29.5 | | 2,592 | 32 | 990 | 101 | 46 | 3,761 |
| Expected Cases, if unvaccinated -> | 35–44 | 39.5 | | 4,006 | 195 | 2,013 | 498 | 184 | 6,896 |
| Expected Cases, if unvaccinated -> | 45–54 | 49.5 | | 6,250 | 227 | 3,242 | 1,955 | 222 | 11,896 |
| Expected Cases, if unvaccinated -> | 55–64 | 59.5 | | 4,426 | 617 | 5,289 | 1,703 | 3,490 | 15,525 |
| Expected Cases, if unvaccinated -> | 65–74 | 69.5 | | 3,433 | 519 | 4,470 | 32 | 8,603 | 17,057 |
| Expected Cases, if unvaccinated -> | 75–84 | 79.5 | | 6,824 | 1,136 | 853 | 19 | 4,539 | 13,371 |
| Total Expected Cases, if unvaccinated -> | | | | 28,444 | 2,824 | 17,300 | 4,346 | 17,122 | 70,036 |
| | Age | Age | | Pfizer- | Moderna | Sputnik-V | AstraZenec | Sinopharm | ALL |
| | | | 7.00E-05 | Expected Deaths, in matched indiduals, unvaccinated | | | | | |
| | | | IFR | Pfizer- | Moderna | Sputnik-V | AstraZenec | Sinopharm | |
| Expected Deaths, if unvaccinated -> | 16–24 | 20 | 0.0006 | 0 | 0 | 0 | 0 | 0 | 0 |
| Expected Deaths, if unvaccinated -> | 25–34 | 29.5 | 0.0016 | 0 | 0 | 0 | 0 | 0 | 0 |
| Expected Deaths, if unvaccinated -> | 35–44 | 39.5 | 0.0047 | 2 | 0 | 0 | 0 | 0 | 3 |
| Expected Deaths, if unvaccinated -> | 45–54 | 49.5 | 0.0138 | 8 | 0 | 1 | 4 | 0 | 14 |
| Expected Deaths, if unvaccinated -> | 55–64 | 59.5 | 0.0400 | 17 | 1 | 6 | 11 | 18 | 53 |
| Expected Deaths, if unvaccinated -> | 65–74 | 69.5 | 0.1163 | 39 | 2 | 15 | 1 | 131 | 187 |
| Expected Deaths, if unvaccinated -> | 75–84 | 79.5 | 0.3381 | 225 | 12 | 8 | 1 | 201 | 447 |
| Total Expected Deaths, if unvaccinated -> | | | | 292 | 15 | 32 | 17 | 350 | 705 |
| **Gompertzian Reduction** | | | | Pfizer- | Moderna | Sputnik-V | AstraZenec | Sinopharm | ALL |
| Observed CFR/Expected CFR-> | | | | 0.55107 | 0.75426 | 0.3805 | 0.5857 | 0.56567 | 0.55573 |
| | | | +/- 95% -> | 0.0185 | 0.0905 | 0.0423 | 0.0368 | 0.0205 | 0.0123 |
| Reduction, as a % (1-v/un) | | | | 45% | 25% | 62% | 41% | 43% | 56% |
| **#-Fold Reduction in Death, once infected ->** | | | | **1.81** | **1.33** | **2.63** | **1.71** | **1.77** | **1.80** |
| **Pasteurian Reduction** | | | | | | | | | |
| Incidence Vacinated / Incidence UnVacinated -> | | | | 0.0976 | 0.0308 | 0.02931 | 0.15854 | 0.13065 | 0.13065 |
| Reduction, as a % (1-v/un) | | | | 90% | 97% | 97% | 84% | 87% | 87% |
| **#-Fold Reduction in Infection -** | | | | **10.25** | **32.46** | **34.12** | **6.31** | **7.65** | **7.65** |
| **Malthusian Reduction** (ie Combined) | | | | 0.05378 | 0.02323 | 0.01115 | 0.09286 | 0.0739 | 0.0726 |
| Reduction, as a % (1-v/un) | | | | 95% | 98% | 99% | 91% | 93% | 93% |
| **#-Fold Reduction in Death, Overall ->** | | | | **18.59** | **43.04** | **89.67** | **10.77** | **13.53** | **13.77** |

APPENDIX-TABLE II

***Gompertzian Analysis*** of COVID-19 Vaccination: ISRAEL, Vaccine Impact

Israel engaged in a nationwide vaccination program (Pfizer) in December 2020, with a number of excellent studies, quite comparable, examining the is impact of the vaccine in the first months of 2021. For our purposes here, Goldberg et al[19] is more relevant, with data from December 20, 2020, to March 20, 2021.

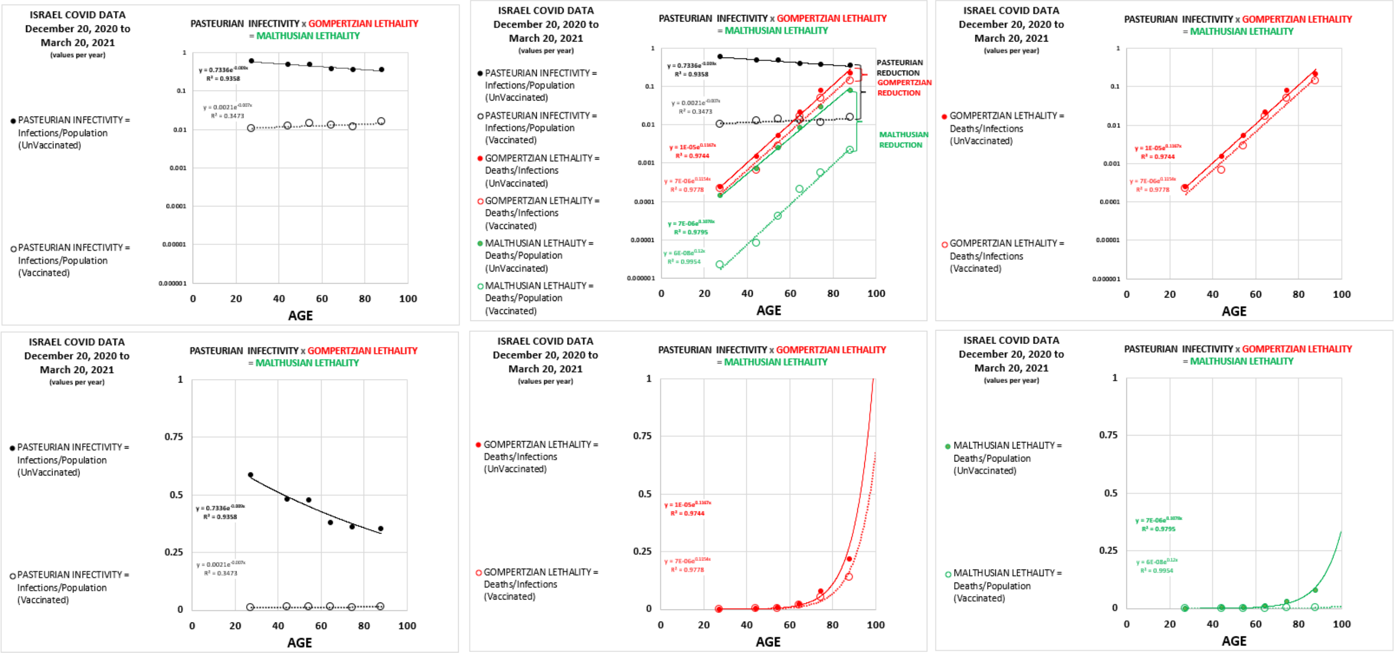


APPENDIX-FIGURE 15

From these data, my calculations of the average ***Gompertzian Reduction***, that is the reduction in ***Gompertzian Lethality*** (Deaths/Cases) of vaccinated compared to unvaccinated individuals, averaged over all of the age groups, was 41%. The average ***Pasteurian Reduction***, that is the reduction in ***Pasteurian Infectivity*** (Cases/Population) of vaccinated compared to unvaccinated individuals, of all of the age groups, was 97%. The average ***Malthusian Reduction***, that is the reduction in ***Malthusian Lethality*** (Deaths/Population) of vaccinated compared to unvaccinated individuals, of all of the age groups, was 98%. Note yet again how the ***Gompertz Lines*** of the vaccinated and unvaccinated are roughly parallel, indicating roughly equivalent reductions in all age groups (APPENDIX-FIGURE 14).

Goldberg et al went further to examine cohorts of patients, differing by the amount of time since the first and second dose, and 384 individuals with COVID-19 infection who had been previous infected, among which there was only one death. These data from these cohorts also reveal that ***Gompertzian Reduction*** (Deaths/Cases) improved for first to second doses, and with time. Remarkably, the previously infected individuals had ***Gompertzian*** and ***Pasteurian Reductions*** from the infection that endowed them with ***Gompertzian*** and ***Pasteurian Reductions*** in future infections that were as good as, and perhaps even better than, the benefit of vaccination seen in individuals who had never been infected.

| | Cohort 0: unvaccinated and not previously infected. | Cohort 1A: followed from day 1 to day 14 after the first vaccine dose. | Cohort 1B: followed from 15 days after the first dose to 13 days after the second dose. | Cohort 2: followed from 14 days after the second dose onwards. | Recovered cohort: previously infected individuals. |
|---|---|---|---|---|---|
| **Gompertzian Reduction**: 1- (Actual # of Deaths/Expected # of Deaths [for Age-Matched, Unvaccinated, Individuals]) | 4% | 36% | 39% | 50% | 83% |
| **X-Fold** Reduction in Lethality by Vaccination or Previous Disease | 1.04 | 1.55 | 1.64 | 2.01 | 5.99 |

APPENDIX-TABLE III

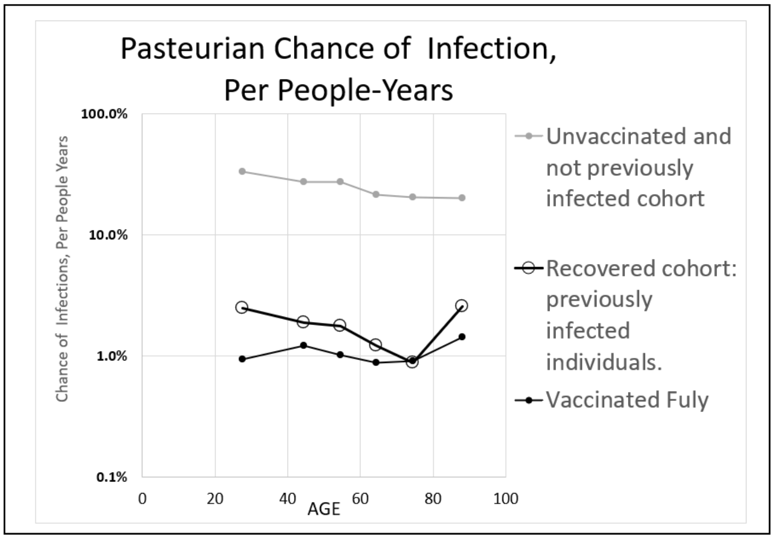


APPENDIX-FIGURE 16

***Gompertzian Analysis*** of COVID-19 Vaccination: POPULATION COMPARISIONS

A comparison of the results of the ***Gompertzian Analysis*** of data on COVID-19 vaccination from each of the four studies described above (Israel, 12/20/21-3/20/21; Hungary, 1/22/21-6/10/21; USA(CDC), 3/1/22-6/20/22; USA(VA), 12/11/20-6/30/21) gives us our first look at how ***Gompertzian Lethality*** (Deaths/Cases) ***Pasteurian Infectivity*** (Cases/Population) and ***Malthusian Lethality***, (Deaths/Population) may change over time (APPENDIX-FIGURE 17, APPENDIX-TABLE IV). In the Israel population, the reduction in COVID-19 ***infectivity***, measured by ***Pasteurian Reduction*** (Cases/Population) was much stronger than the reduction in COVID-19 lethality, measured by ***Gompertzian Reduction*** (Deaths/Cases). In contrast, in the USA(CDC), population, the reverse was seen, with the reduction in COVID-19 lethality, the ***Gompertzian Reduction*** (Deaths/Cases) being slightly stronger than the reduction in COVID-19 ***infectivity***, the ***Pasteurian Reduction*** (Cases/Population). The Hungary and USA(VA) populations fell in between. These values suggest, again, that change in COVID-19 ***Gompertzian Lethality*** measured by the ***Gompertzian Reduction*** of Deaths/Cases, can be independent of change in COVID-19 ***Pasteurian Infectivity*** measured by the ***Pasteurian Reduction*** of Cases/Population, even though vaccination is at work on both effects.

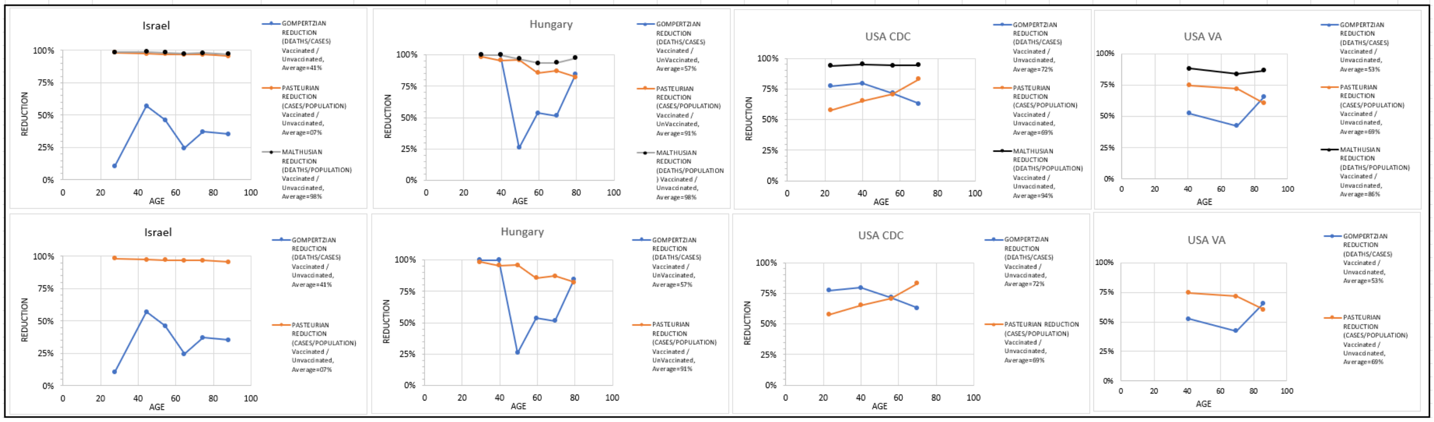


APPENDIX-FIGURE 17

| STUDY | GOMPERTZIAN REDUCTION (CASES / DEATHS) "1-(Vaccinated / Unvaccinated)" | PASTEURIAN REDUCTION (CASES / POPULATION) "1-(Vaccinated / Unvaccinated)" | MALTHUSIAN REDUCTION (DEATHS / POPULATION) "1-(Vaccinated / Unvaccinated)" | GOMPERTZIAN / PASTEURIAN |
|---|---|---|---|---|
| Israel, 12/20/21-3/20/21 | 41% | 97% | 98% | 0.43 |
| Hungary, 1/22/21-6/10/21 | 58% | 91% | 96% | 0.63 |
| USA(CDC), 3/1/22-6/20/22 | 53% | 69% | 86% | 0.77 |
| USA(VA), 12/11/20-6/30/21 | 73% | 69% | 94% | 1.05 |

APPENDIX-TABLE IV

***Gompertzian***, ***Pasteurian***, and ***Malthusian Reductions***, of four studies, each an average of the age groups.

***Gompertzian Analysis*** of COVID-19 Vaccination: COVID-19 Examined Over Time

USA(CDC): April 2021 to June 2022

Let us now move on from examining COVID-19 datasets as single chunks of data, to examining data assembled to capture how COVID-19's impact has changed over time. We begin by returning to the rather extraordinary CDC data on approximately 70% of all COVID-19 cases in the USA. The first dataset, from April 2021 to June 2022, contains data on patients sorted by vaccination status, but no booster information. Thus, we have data over the period from three months after the first vaccines became available (late December 2021), beginning when the alpha variant was predominant, and then followed by Delta (~August 2021), and then Omicron (~December 2022+). During this period, the first boosters were approved (late September 2021), then the second boosters (end of March 2022), and the third boosters were approved September 2022. This dataset didn't contain booster data, but a second dataset, discussed below, does (see "***Gompertzian Analysis*** of COVID-19 Vaccination: COVID-19 Examined by Dose" below).

At the beginning of the first CDC dataset, April 2021, about 25% of Americans were vaccinated, and thus about 75% unvaccinated. By the end of this dataset, June 2022, the reverse was the case, with about 75% of Americans being vaccinated and about 25% unvaccinated (APPENDIX-FIGURE 18).

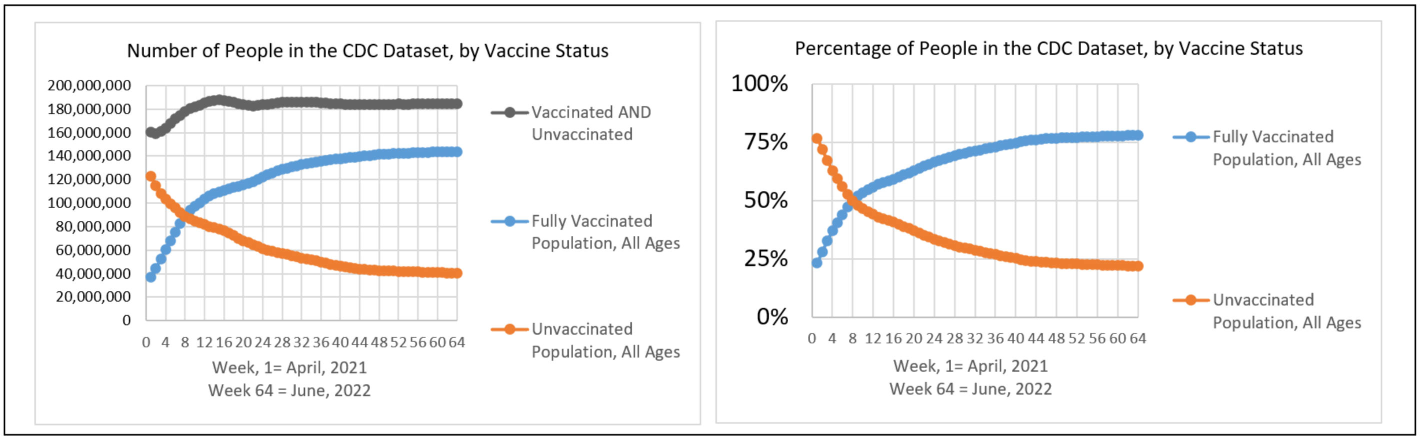


APPENDIX-FIGURE 18

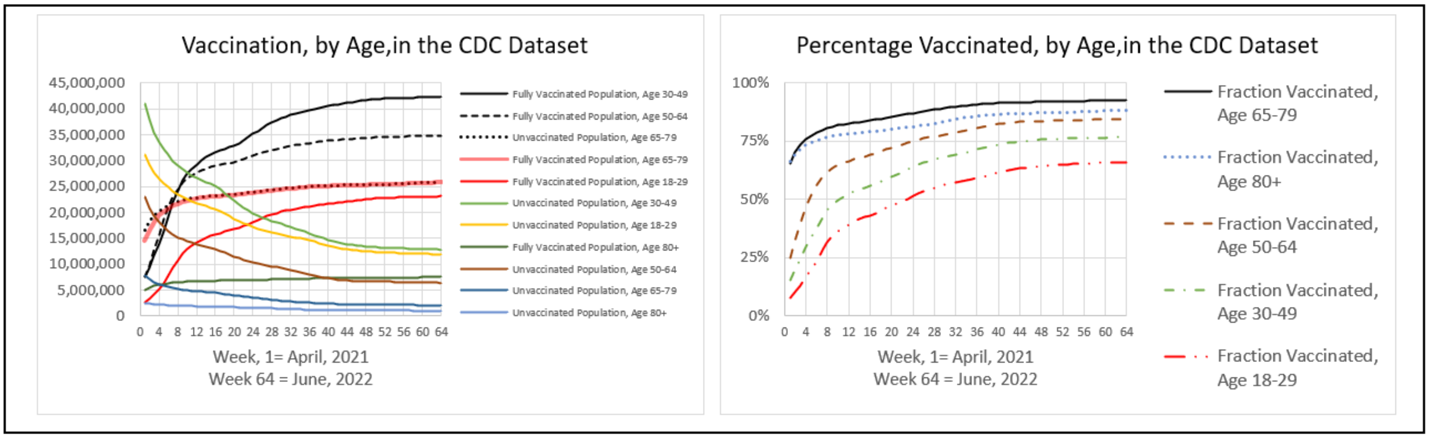


APPENDIX-FIGURE 19

In parallel, in the first dataset, from April 2021 the percentage of COVID-19 cases occurring among the vaccinated rose from about 25% of all cases to about 75% by June 2022 (APPENDIX-FIGURE 20).

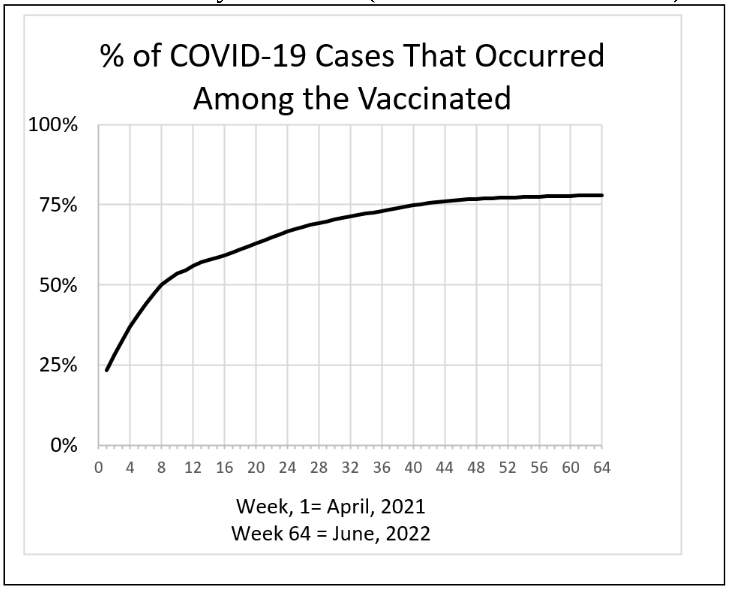


APPENDIX-FIGURE 20

Vaccinated patients had a markedly lower rate of death, with about 70,000 lives lost among the vaccinated from April 2021 to June 2022, and about 180,00 lives lost among unvaccinated during this time period (APPENDIX-FIGURE 21). In terms of the populations of the vaccinated and unvaccinated individuals, the unvaccinated had about six times the frequency of death (APPENDIX-FIGURE 21).

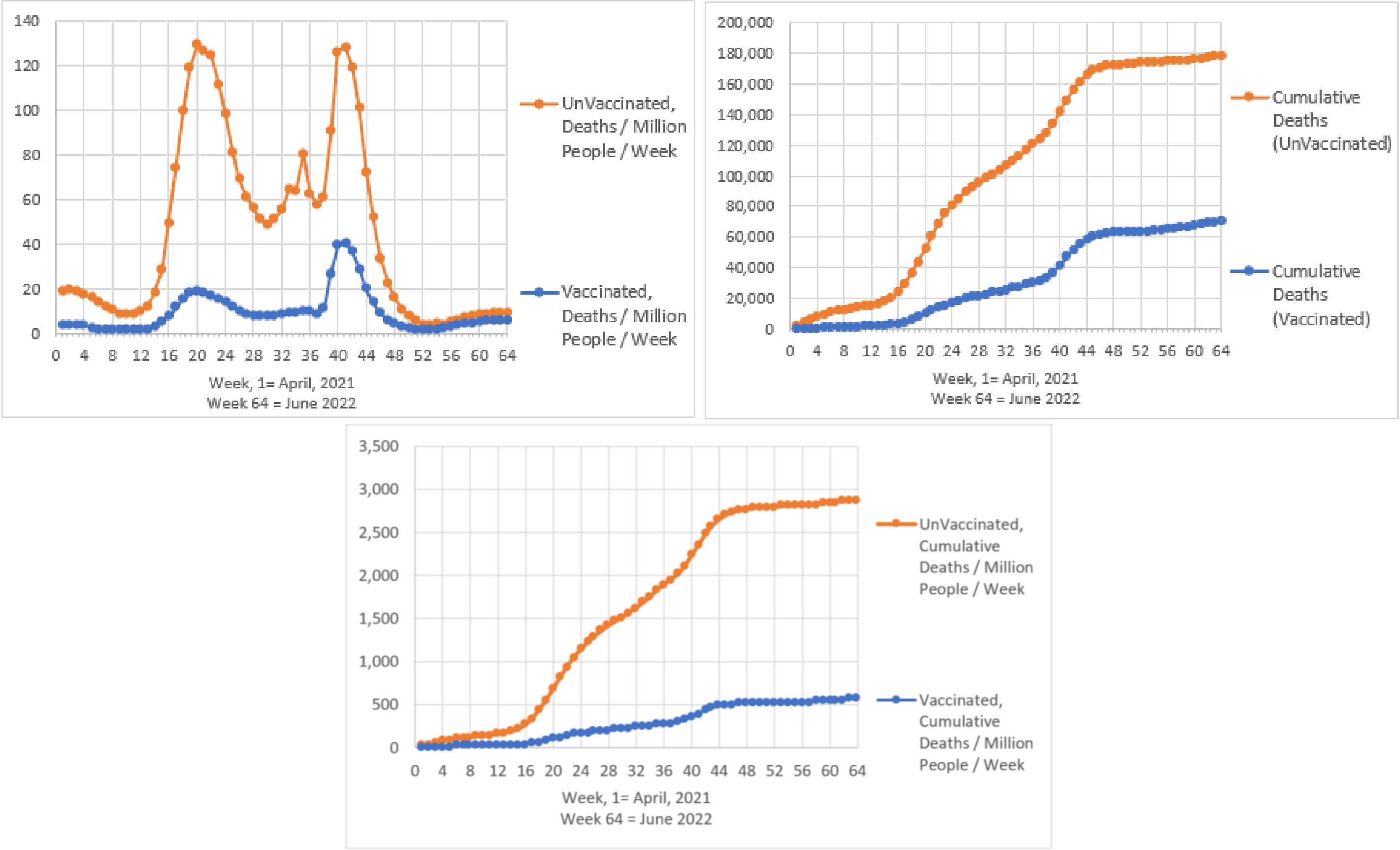


APPENDIX-FIGURE 21

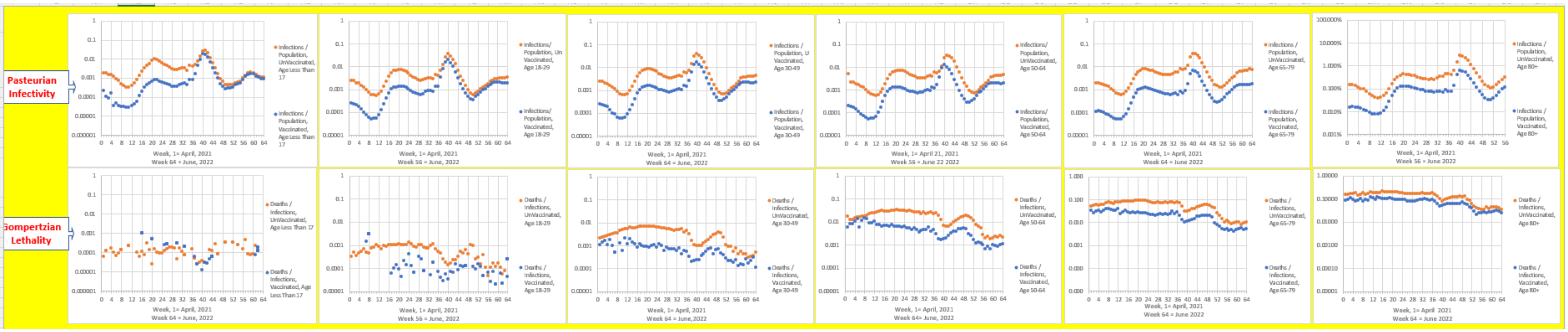


APPENDIX-FIGURE 22

USA(CDC): ***Gompertzian Lethality*** and ***Pasteurian Infectivity*** April 21-June 22, by age.

In APPENDIX-FIGURE 22 are shown values for ***Pasteurian Infectivity*** (Cases/Population) and ***Gompertzian Lethality*** (Deaths/Cases), for the vaccinated and unvaccinated, for each week, from April 2021 to June 2022. The Y-Axis for these figures is on a logarithmic scale, in units of 10, so a dot that is twice as high as another dot captures a 10-fold higher value, and a dot that's three time as high captures a 100-fold higher value, etc. The series of dots form wave-like images, from April 2021 on the left of each graph, to June 2022 on the right, visualizing how ***Pasteurian Infectivity*** (Cases/Population) and ***Gompertzian Lethality*** (Deaths/Cases) change over time, for the vaccinated, and the unvaccinated. There is a great deal of information in these figures, which we shall unpack, item by item:

Age has little impact on ***Pasteurian Infectivity***:

Age had no perceptible impact on ***Pasteurian Infectivity*** (Cases/Population) for both vaccinated and unvaccinated individuals. That is, every week, the ratio of Cases/Population was almost the same for every age group. 85-year-olds were about as likely to get COVID-19 as teenagers (APPENDIX-FIGURES 22-25).

Age has massive impact on ***Gompertzian Lethality***:

Age had a dramatic exponential impact on ***Gompertzian Lethality*** (Deaths/Cases) for both vaccinated and unvaccinated individuals. 85-year-olds were a thousand-fold more likely to die of COVID-19, once infected, than teenagers. Thus, the CDC data showed the very same frightening exponential impact of case fatality as age increases, taking the form of the ***Gompertz Mortality Equation*** (APPENDIX-FIGURES 1 and 2 and many other figures throughout this APPENDIX). This can, perhaps, be more easily envisaged when we display this information on ***Gompertz Plots***, that is to say, on log graphs, where the chance of death, ***D***, ***Gompertzian Lethality*** (Deaths/Cases), is compared with age, ***t***, as we shall see below.

Vaccinated people had lower rates of ***Gompertzian Lethality*** than unvaccinated people:

Vaccinated people always had lower rates of ***Gompertzian Lethality*** (Deaths/Cases) than unvaccinated people. The virus never defeated this benefit of vaccines, the ***Gompertzian Reduction*** in the risk of death once infected, which grew stronger as the pandemic proceeded (APPENDIX-FIGURES 22-25).

***Pasteurian Infectivity*** went up and down wildly, ending 10-fold higher than at the beginning:

For both vaccinated and unvaccinated individuals, ***Pasteurian Infectivity*** (Cases/Population) went up and down wildly with time, with the huge waves, varying over a hundred+-fold difference in height, and displaying, over the long term, a roughly a 10-fold increase in the chance of infection from April 2021 to June 2022 (APPENDIX-FIGURES 22-25). Vaccinated people had lower rates of ***Pasteurian Infectivity*** (Cases/Population) than unvaccinated people, but the virus kept defeating the vaccines.

***Gompertzian Lethality*** declined progressively over time:

For both vaccinated and unvaccinated individuals, ***Gompertzian Lethality*** (Deaths/Cases) went up and down mildly with time, with small waves that varied several-fold in height. However, over the long term, ***Gompertzian Lethality*** declined roughly 10-fold from April 2021 to June 2022 (APPENDIX-FIGURES 22-25).

***Gompertzian Lethality*** declined progressively over time, heading toward zero death for the vaccinated:

Graphing ***Gompertzian Lethality*** (Deaths/Cases), sorted by age, among the vaccinated, over time, on a linear scale, reveals all age groups pointing down to zero over the next few months (APPENDIX-FIGURE 23).

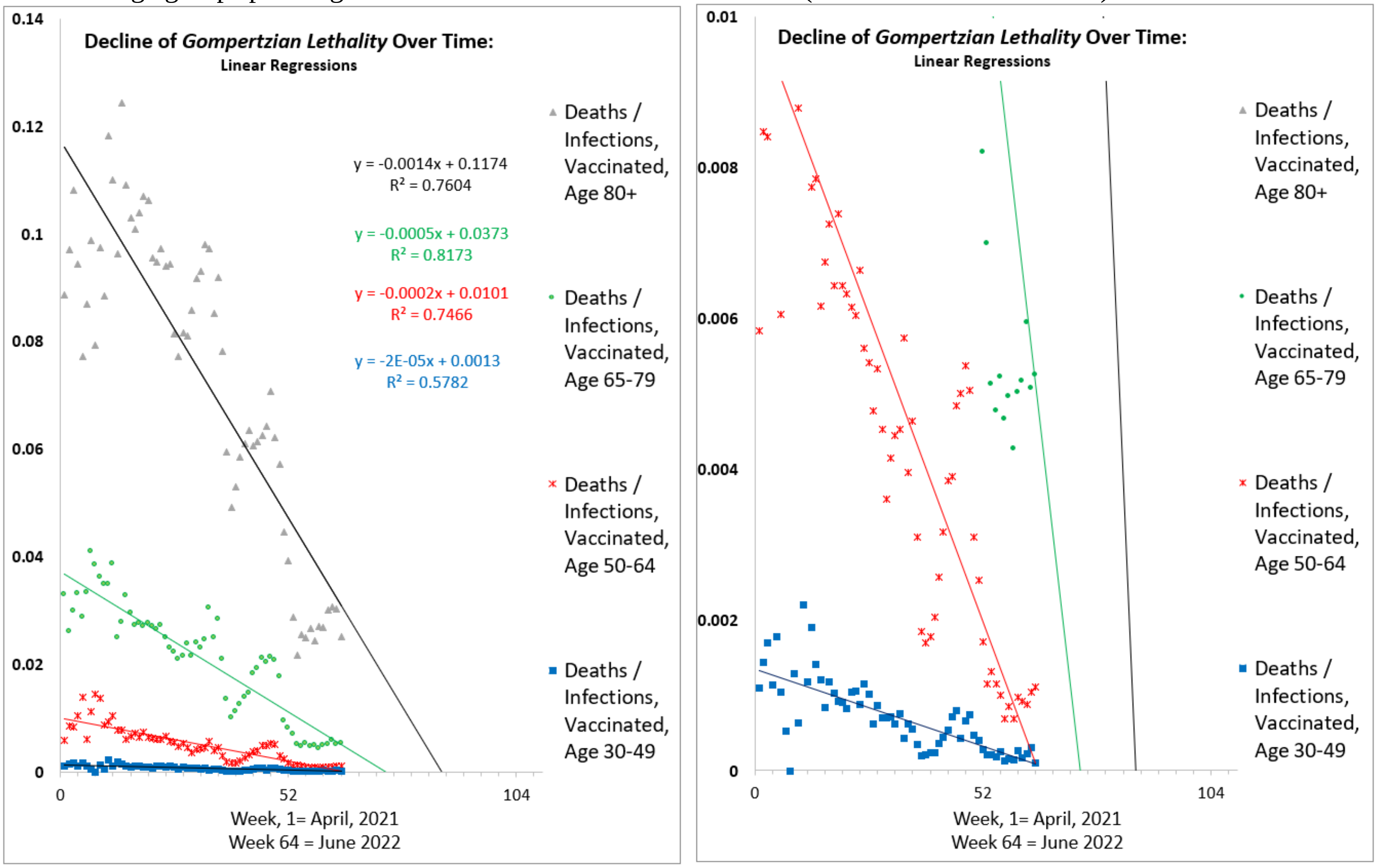


(APPENDIX-FIGURE 23)

Precisely why ***Gompertzian Lethality*** (Deaths/Cases) among the vaccinated declined over this 15-month period is not contained in these data. Boosters, infections, new variants, or other forces could be behind these changes. However, as we shall see below, using a second dataset with booster information, it was found that boosters are likely to be a major force in lessening ***Gompertzian Lethality*** (Deaths/Cases).

***Gompertzian Lethality***, declined linearly, heading toward zero; ***Pasteurian Infectivity*** exploded erratically, heading upward.

Comparison of ***Gompertzian Lethality*** with ***Pasteurian Infectivity*** reveals how much progress has occurred in reducing COVID-19 lethality, and how little in progress has occurred in reducing COVID-19 infection. This is illustrated in APPENDIX-FIGURE 24 for Age 65-79 individuals, but it is seen in all age groups.

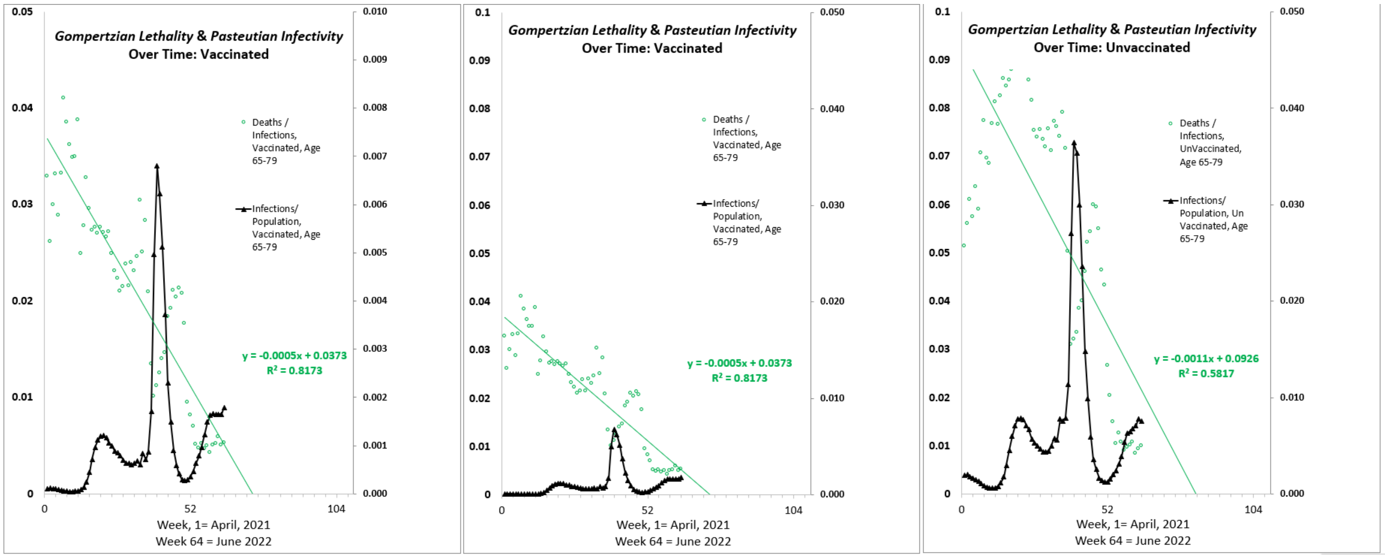


APPENDIX-FIGURE 24

COVID-19, by age, on ***Gompertz Plots*** Show ***Gompertzian Lethality*** Declined by ***Gompertzian Height***:

Graphs of the log of ***Gompertzian Lethality*** (Deaths/Cases) vs (age), for monthly groups of vaccinated COVID-19 patients revealed ***Gompertz Lines*** of ***Gompertzian Lethality***, declining progressively in ***Gompertzian Height***, with little evident change in the ***Gompertzian Slope*** (APPENDIX-FIGURE 25). That is to say, ***Gompertzian Lethality*** declines by roughly equivalently improvement across all ages. In these ***Gompertz Plots*** of COVID-19 lethality, as we had seen in the raw data weekly shown in FIGURE 22, we again see the roughly a 10-fold decline in ***Gompertzian Lethality*** that occurred among vaccinated individuals between April 2021 to June 2022.

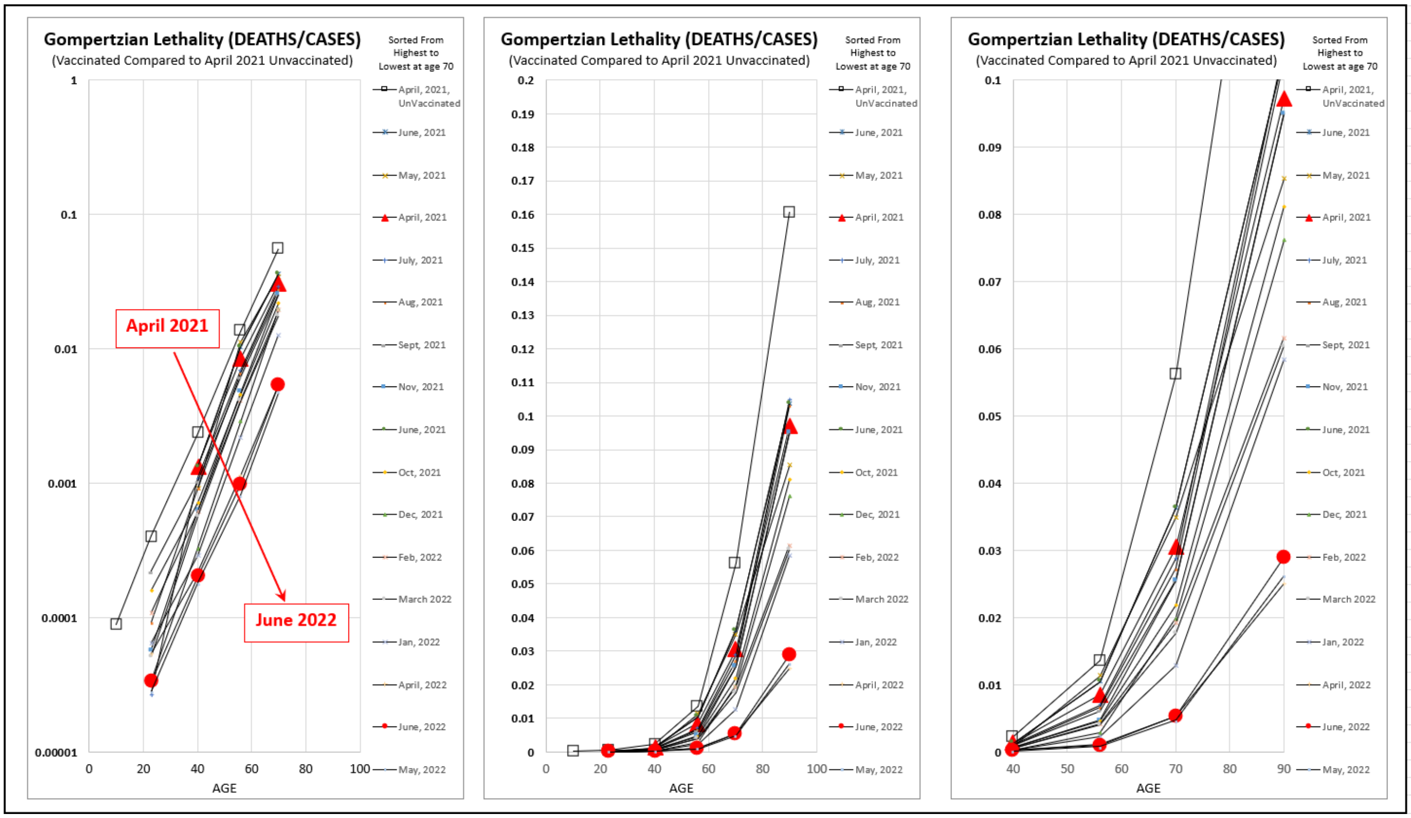


APPENDIX-FIGURE 25

Curiously, the unvaccinated groups also underwent such a decline in ***Gompertzian Lethality***, but never catching up with the vaccinated (not shown). Perhaps this decline ***Gompertzian Lethality*** among the unvaccinated was created by COVID-19 infection, as we saw above from the Israel cohort of recovered COVID-19 individuals (APPENDIX-TABLE III). If so, the sad price for the reduction in the ***Gompertzian Lethality*** among the unvaccinated was the loss of life that occurred after some of these infections; by June 2022, unvaccinated Americans were ¼th of the population in this dataset (APPENDIX-FIGURE 18, APPENDIX-FIGURE 19) but had 2½ times more deaths than the vaccinated (APPENDIX-FIGURE 21).

***Gompertzian Analysis*** of COVID-19 Vaccination: COVID-19 Examined by Dose

Let us now examine how COVID-19 changes with sequential vaccination.

I. USA(CDC): March 2022 to July 2022, Boosters.

The second CDC dataset, from March 2022 through July 2022, thus one month beyond the first dataset described above, contains additional information on booster status[20] (APPENDIX-FIGURE 26). The first boosters were approved in late September 2021, the second boosters at the end of March 2022, and the third boosters in September 2022. The CDC dataset of roughly 65 million Americans shows that as of June 2022, about 10 million had been vaccinated to 2 boosters, ~20 million had been vaccinated to 1 booster, ~57 million vaccinated but no boosters yet, and ~10 million were unvaccinated (APPENDIX-FIGURE 26).Thus, about 87% of the population had any vaccination, 39% had one booster, 17% had two boosters, 31% had vaccination but no boosters, and 13% had no vaccination. This dataset has two age groups (50-64 and 65+).

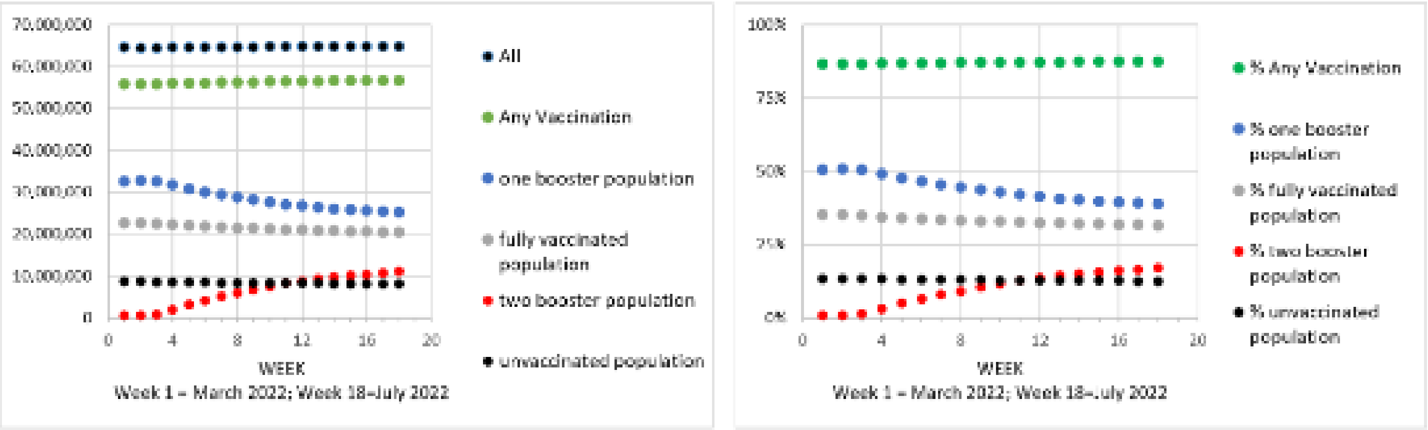


APPENDIX-FIGURE 26

#### 1. Boosters: ***Gompertzian Lethality*** (Deaths/Cases)

The graphs in APPENDIX-FIGURE 28 below shown ***Gompertzian Lethality*** (Deaths/Cases), for each of the 18 weeks, covered between March 2022 and July 2022. These values show that the chance of death after infection, ***Gompertzian Lethality*** (Deaths/Cases), was highest for those who are unvaccinated, lower for those vaccinated without boosters, even lower for those vaccinated, followed by 1 booster, and lowest yet for those with 2 boosters. The magnitude of these beneficial effects of boosters are measured by their ***Gompertzian Reductions*** (**1**-[vaccinated/unvaccinated], Deaths/Cases), averaged for the two age groups (50-64 and 80+). shown in (APPENDIX-FIGURE 27). For those vaccinated without boosters, the ***Gompertzian Reduction*** was 23%, a 1.3-fold reduction in (Deaths/Cases). For those vaccinated followed by 1 booster ***Gompertzian Reduction*** was 59%, a 2.5-fold reduction in (Deaths/Cases). For those vaccinated followed by 2 boosters ***Gompertzian Reduction*** was 67%, a 3-fold reduction in (Deaths/Cases).

These data also give some hints, however indirect, for how long the ***Gompertzian Reduction*** persist after boosters are given. The "1" booster group is perhaps most informative, as its numbers decline over time, no doubt because some of them become "2" boosters (APPENDIX-FIGURE 26). Thus, we suspect that few individuals are added to the "1" booster group as the study proceeded. In this regard, it is striking that there was no evident waning of ***Gompertzian Lethality*** for this "1" booster group over the 18 weeks in the dataset, from March 2022 to July 2022 (APPENDIX-FIGURE 27, and other FIGURES shown here). Of course, this persistence of ***Gompertzian Lethality*** deserves more complete attention, using data on the time since injection.

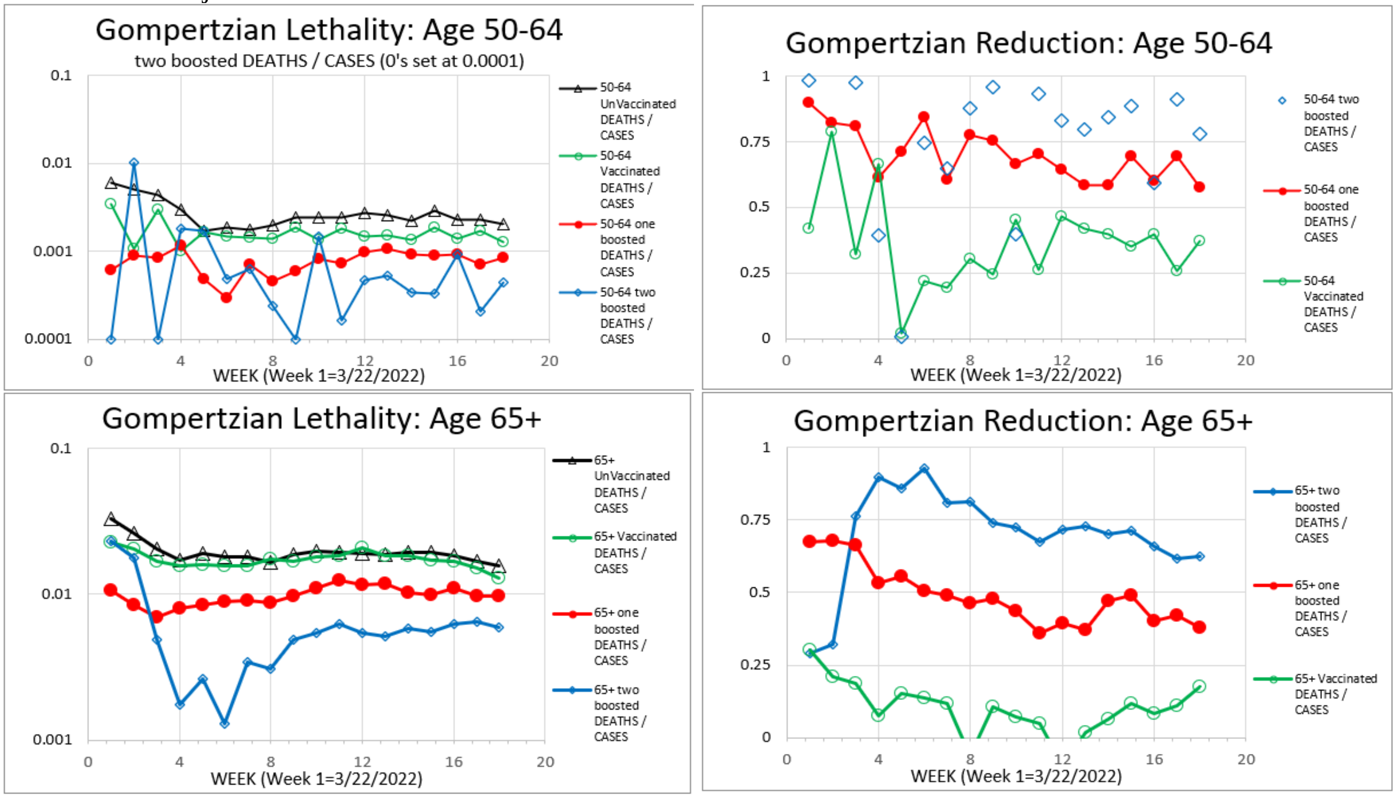


APPENDIX-FIGURE 27

2. Boosters: ***Pasteurian Infectivity*** (Cases/Population)

The graphs in APPENDIX-FIGURE 28 below show ***Pasteurian Infectivity*** (Cases/Population) for each of the 18 weeks, covered between March 2022 and July 2022. These values reveal that the chance of infection is highest for those who are unvaccinated and lower for those vaccinated without boosters. Boosters did not appear to have added any obvious additional benefit, and, in fact, those with 1 or 2 boosters showed slightly poorer reduction in ***Pasteurian Infectivity*** than those that have simply been vaccinated, although the differences are so small that they may be irrelevant.

The magnitude of these beneficial effects of vaccination and boosters are measured by their ***Pasteurian Reductions*** (1-[vaccinated/unvaccinated]; Cases/Population), shown in (APPENDIX-FIGURE 28), averaged for the two age groups (50-64 and 80+). For those vaccinated without boosters, the ***Pasteurian Reduction*** was 73%, a 3.6-fold reduction in (Cases/Population). For those vaccinated followed by 1 booster ***Pasteurian Reduction*** was 53%, a 2.1-fold reduction in (Cases/Population). For those vaccinated followed by 2 boosters ***Pasteurian Reduction*** was 63%, a 2.7-fold reduction in (Cases/Population).

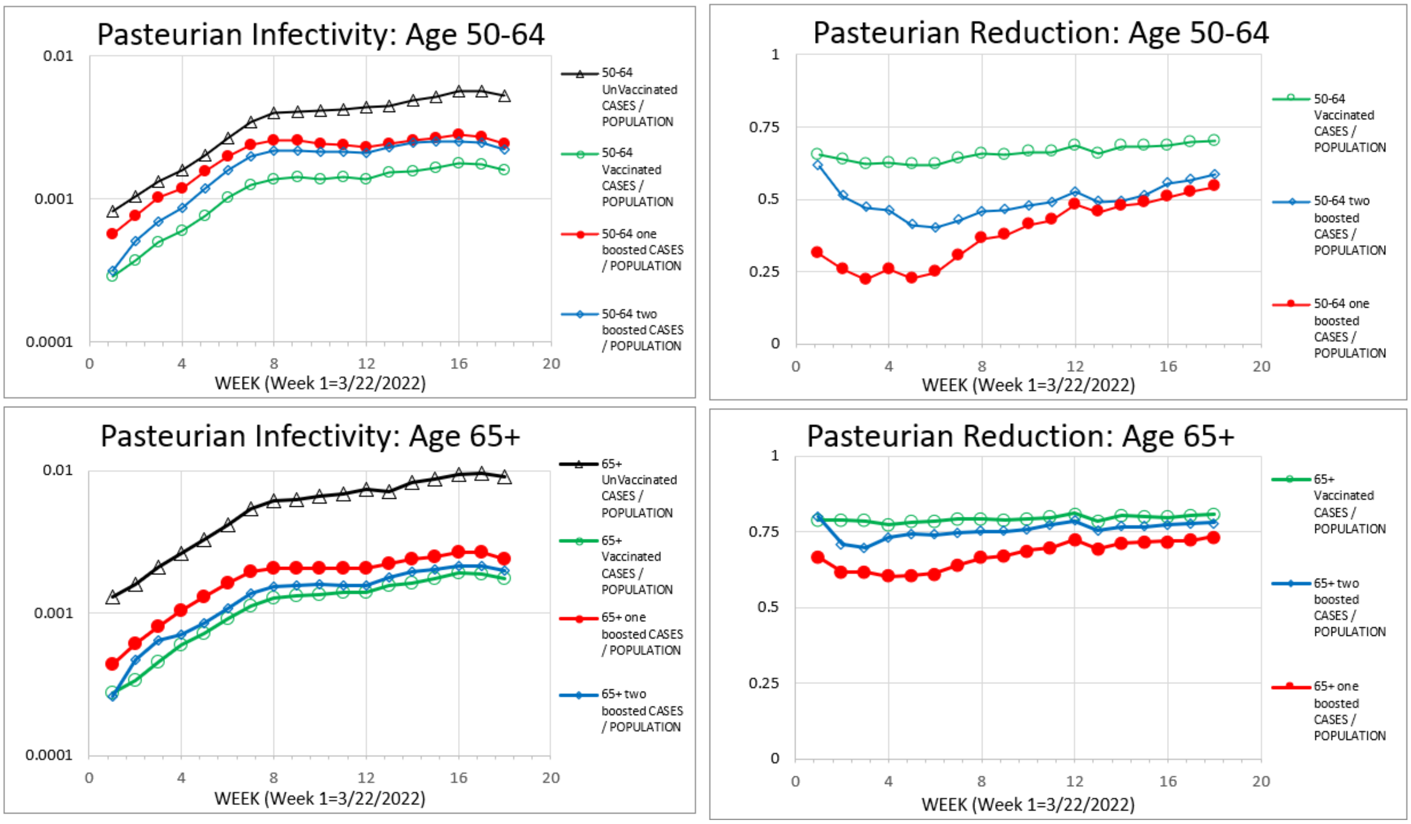


(APPENDIX-FIGURE 28)

Note that while ***Gompertzian Lethality*** (Deaths/Cases) remained relatively constant across the 18 weeks, as indeed across the previous year+, in aggregate, showing a 10-fold decline, ***Pasteurian Infectivity*** (Cases/Population) increased progressively over the first 8 weeks of this dataset, for both the vaccinated, and the unvaccinated, and then showed a milder increase over the next 10 weeks. This is in agreement with the data from the previous year+ noted above, when ***Pasteurian Infectivity*** (Cases/Population) went up and down wildly, varying with over a hundred+-fold differences in height, displaying, over the long term, the roughly a 10-fold increase in the chance of infection from April 2021 to June 2022. Again, vaccination appears to give a lower chance of infection, but not control of the pandemic. Fortunately, though, vaccination does lower the burden of death, seen in the ***Gompertzian*** and ***Malthusian Lethalities***, as we shall examine next.

3. Boosters: ***Gompertzian Lethality***, ***Pasteurian Infectivity***, ***Malthusian Lethality*** compared

In APPENDIX-FIGURE 29, the ***Gompertzian Lethality*** (Deaths/Cases), averaged over all 18 weeks, is displayed against vaccination. On the graph, “0” identifies no vaccination, “1” identifies vaccination without boosters, “2” identifies vaccination followed by 1 booster, “3” identifies vaccination followed by 2 boosters. Remarkably, each dose decreased the risk of death, as seen by linear regression, pointing towards zero lethality at ~3 or 4 boosters.

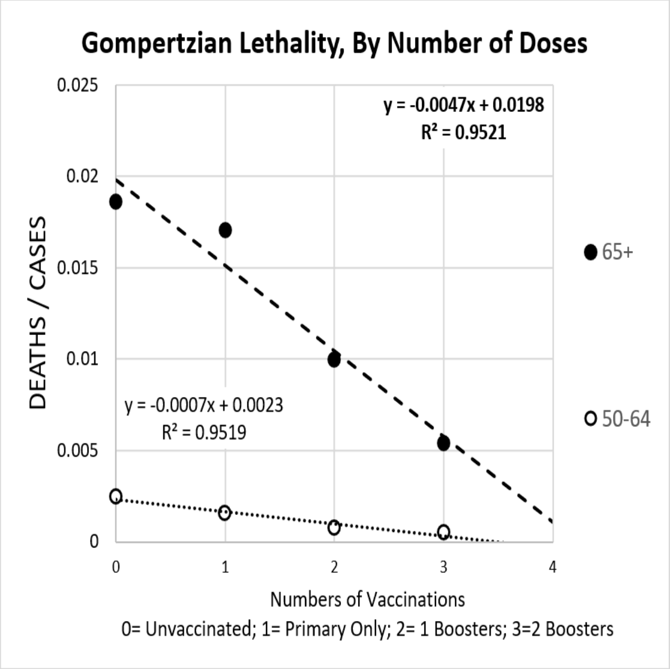


(APPENDIX-FIGURE 29)

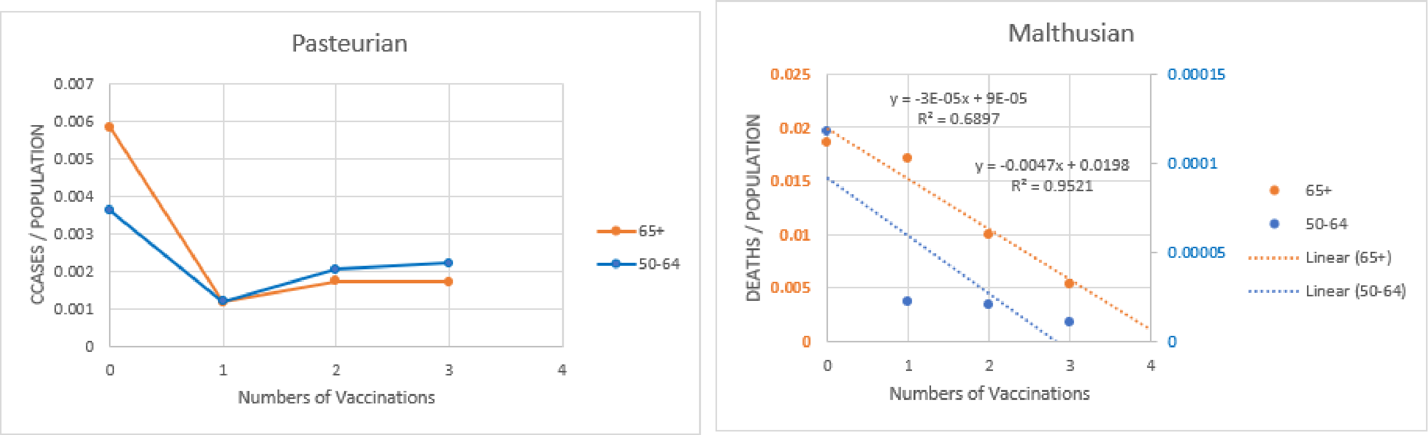


(APPENDIX-FIGURE 30)

In APPENDIX-FIGURE 30, left, the average ***Pasteurian Infectivity*** (Cases/Population) is shown. This graph illustrates vaccination reducing the chance of infection. See also reference 21.

In APPENDIX-FIGURE 30, right, the average ***Malthusian Lethality*** (Deaths/Population) is displayed. These data illustrate the combined benefit of ***Gompertzian*** and ***Pasteurian Reduction***, becoming progressively more beneficial with each booster.

For ***Pasteurian Infectivity*** (Cases/Population), the chance of infection, relative to the unvaccinated, was reduced by the initial vaccination, but this initial benefit became less after the first booster, and even less after the second booster. Whether these changes to ***Pasteurian Infectivity*** were due to weakening action of vaccines among the vaccinated, changed variants, or improved resistance among the unvaccinated due to infection, is not known from these data, although resolving this would be doable, and of considerable value.

The ***Gompertzian Reductions*** achieved by each sequential vaccine dose was sufficient to push the chance of death towards zero. On the other hand, ***Pasteurian Reduction*** was achieved by the initial vaccination, but this was not followed by improvement for the sequential booster doses, thus failing to defeat the virus.

4. Boosters: ***Gompertzian Lethality*** (Deaths/Cases) on ***Gompertz Plots***

Having only two age groups (50-64 and 65+) makes it impossible to examine fit to the ***Gompertz Mortality Equation***. However, the two data points can be placed on a ***Gompertz Plots***, displaying a reduction in the ***Gompertzian Height***, ***$G_H$***, in the presence of a roughly constant ***Gompertzian Slope***, ***$G_s$***, as we have seen in so many examples above (APPENDIX-FIGURE 31).

We can also project these values down to what might be hope for the current third booster (APPENDIX-FIGURES 31 and 32). Again, this exercise in statistical wishful thinking, at least gives an encouraging prediction, against which the benefit of ongoing 3rd booster campaigns can be compared. Thus, the linearization of these ***Gompertz Lines***, shown in APPENDIX-FIGURE 31, and APPENDIX-FIGURE 32, visualize the remarkable progress we have made, and the projections can give us an idea of what we hope for. Indeed, as we shall outline below, results of 3rd booster studies are now coming in, and generally confirming the predictions of such projections.

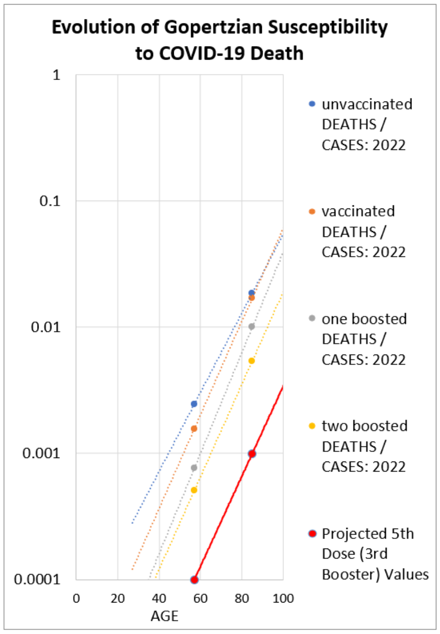


(APPENDIX-FIGURE 31)

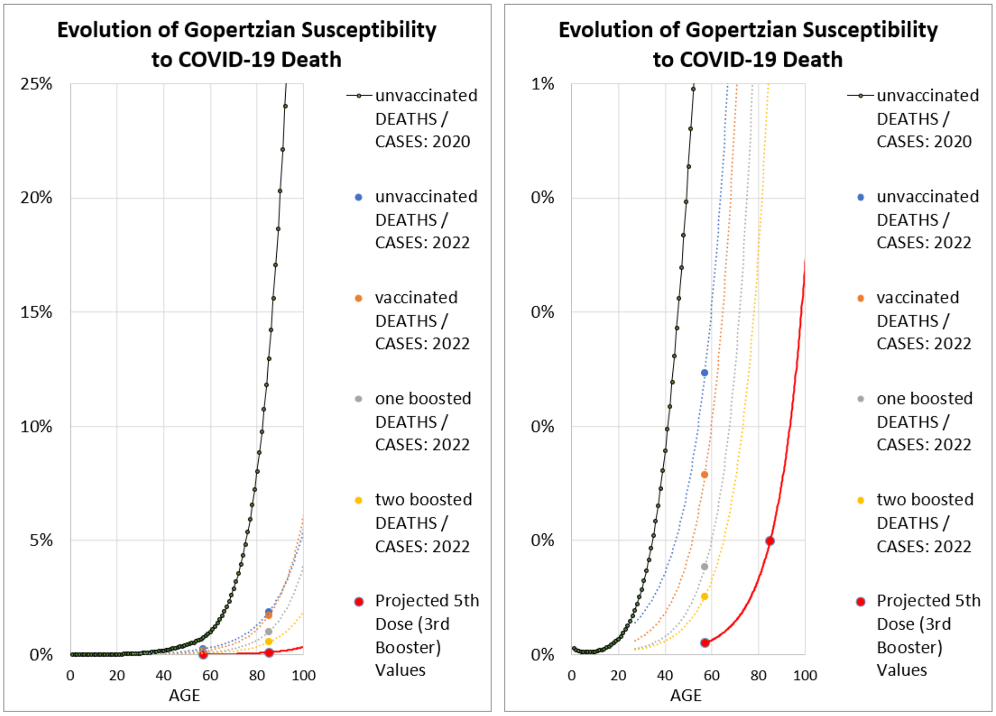


(APPENDIX-FIGURE 32)

#### II. HUNGARY, Boosters

The Hungary group followed the HUN-VE study, analyzed above, with two more studies, HUN-VE 2[22] and HUN-VE 3[23], the first of which is of most relevance here, as it examined the impact of the first and second boosters that followed the primary vaccinations. Values of deaths and cases were provided for patients who had receive no vaccine (0 in APPENDIX-FIGURE 33), Primary vaccination (1), primary vaccination plus just one booster (2), or two boosters (3) in the Omicron Wave, January 1, 2021-February 23, 2021. With their data on cases and deaths, sorted by age, I again could calculate the ***Gompertzian Reductions***: 56% (a 1.80-fold reduction in the lethality) after the Primary Vaccination ("1" on the x-axis, APPENDIX-FIGURE 33, Left); 92%, (an 11.81-fold reduction in the chance of death after infection, ("2" on the x-axis) after the First Booster; 98% (a 55.16-fold reduction in the chance of death after infection) after the Second Booster ("3" on the x-axis).

The practical implications of these reductions in COVID-19 lethality can be seen by graphing the ***Gompertzian*** chance of death, relative to the unvaccinated (APPENDIX-FIGURE 33, Right). These numbers show the events from vaccination ("1" on the x-axis) to first booster ("2" on the x-axis) point to a negligible risk of death, as realized by the second booster ("3" on the x-axis), which forms a straight line on APPENDIX-FIGURE 33, as seen by linear regression. These data reveal that boosters elicit a linearly cumulative, progressively greater, and lasting, resistance to lethality with each sequential dose, pointing to zero mortality after the second booster; that projection seems largely fulfilled by the ***Gompertzian Lethality*** seen after the second booster ("3" on the x-axis, APPENDIX-FIGURE 33). In short, it appears that with boosters, infection may continue, but death may become imperceptible. Above, we have seen precisely this same encouraging impression emerges from USA data, and below we shall see this again in Israeli data. Below, we shall also see data from 2nd booster data that seems to be supporting such a possibility.

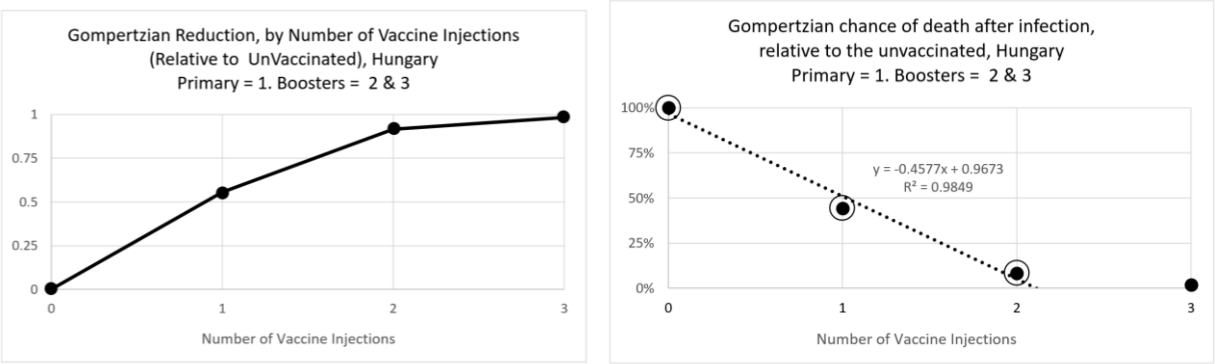


APPENDIX-FIGURE 33

| Gompertzian Reduction | 1 Booster | 2 Boosters | Primary | BOTH |
|---|---|---|---|---|
| Observed CFR/Expected CFR-> | 0.08468 | 0.01813 | 0.55573 | 0.08418 |
| +/- 95% -> | 0.0014 | 0.0126 | 0.0123 | 0.0014 |
| Reduction, as a % (1-v/un) | 92% | 98% | 56% | 92% |
| #-Fold Reduction in Death, once infected -> | 11.81 | 55.16 | 1.80 | 11.88 |

These ***Gompertzian Reductions*** translate into massive saving in life, that becomes progressively more dramatic as we age. As we can see on the ***Gompertz Plots***, and their linear visualizations, shown above, while an unvaccinated 80-year-old at the beginning on the epidemic would have had a 40% chance of death if infected, that number declined to 3% after vaccination and one booster, and 1% after vaccination and two boosters (APPENDIX-FIGURE 34).

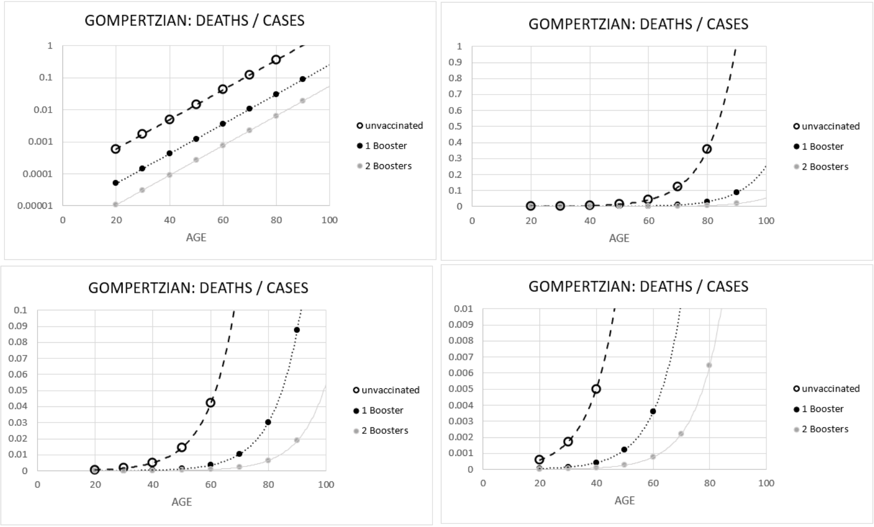


APPENDIX-FIGURE 34yyy

(APPENDIX-FIGURE 34)

Regrettably, for the earlier phase, where primary only individuals, and unvaccinated individuals, were also studied, case numbers were not published, so calculations for the impact of sequential vaccinations and boosters cannot be yet calculated, which, as we shall see later, may well be informative.

##### III. ISRAEL, Boosters

Studies from Israel provides with yet further valuable data in the impact of 1st and 2nd boosters on COVID-19 ***Gompertzian Lethality*** (Deaths/Cases) and ***Malthusian Lethality*** (Deaths/Population).

The first study captured data on deaths and cases by age, making ***Gompertzian Analysis*** of ***Gompertzian Lethality*** (Deaths/Cases) possible.[24] ***Gompertzian Lethality*** (Deaths/Cases), by age, on ***Gompertz Plots***, can be seen in APPENDIX-FIGURE 35. ***Gompertzian Reductions***, relative to the unvaccinated, for patients with just primary vaccination (2 Doses), 1 Booster (3 Doses) or 2 Boosters (4 Doses), can be seen in APPENDIX-FIGURE 36. ***Gompertzian Lethality*** (Deaths/Cases), for each age group, for the unvaccinated ("0" on the X-axis), vaccinated with just the primary ("1" on the X-axis), vaccinated with 1 Booster ("2" on the X-axis), and vaccinated with 2 Boosters ("3" on the X-axis) can be seen in APPENDIX-FIGURE 37.

Most striking of all of these calculations can be seen in APPENDIX-FIGURE 37, where the data show that with each booster, ***Gompertzian Lethality*** declines, again pointing to zero lethality at ~3 or 4 boosters, as we saw above for the USA CDC and Hungarian booster data. We shall see additional data below, again showing this encouraging image.

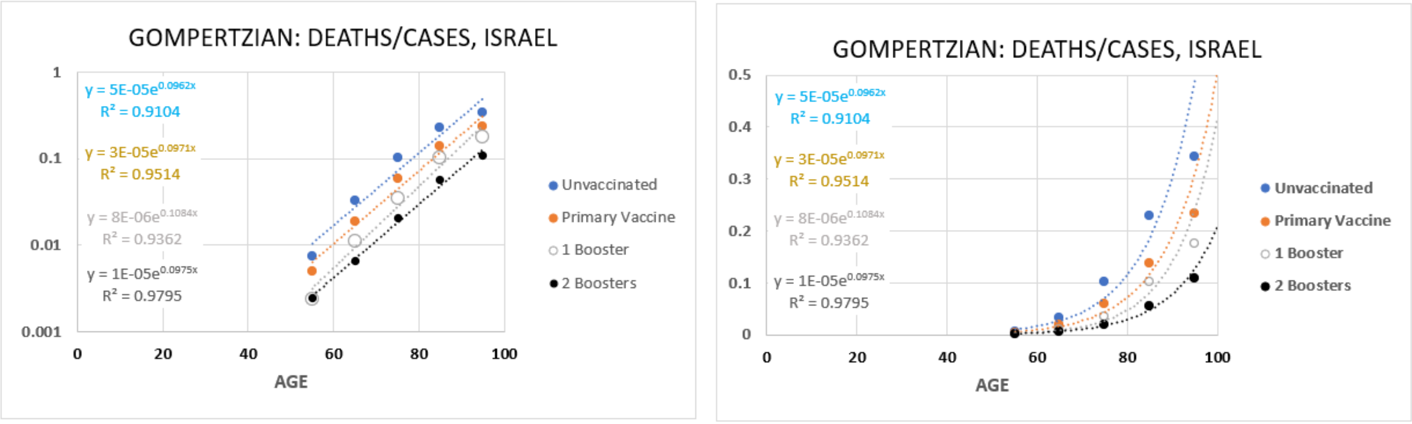


APPENDIX-FIGURE 35

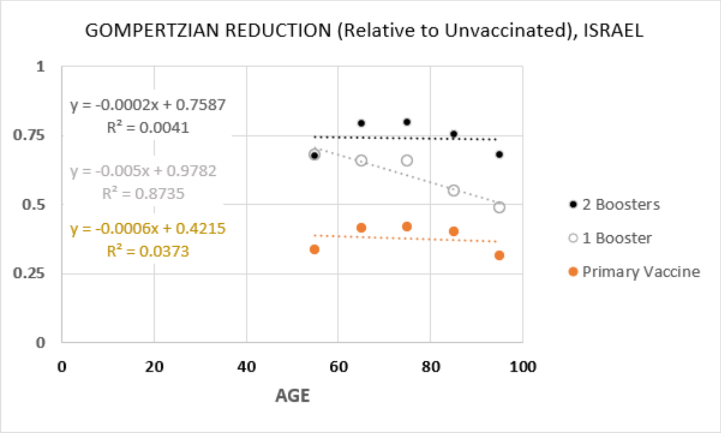


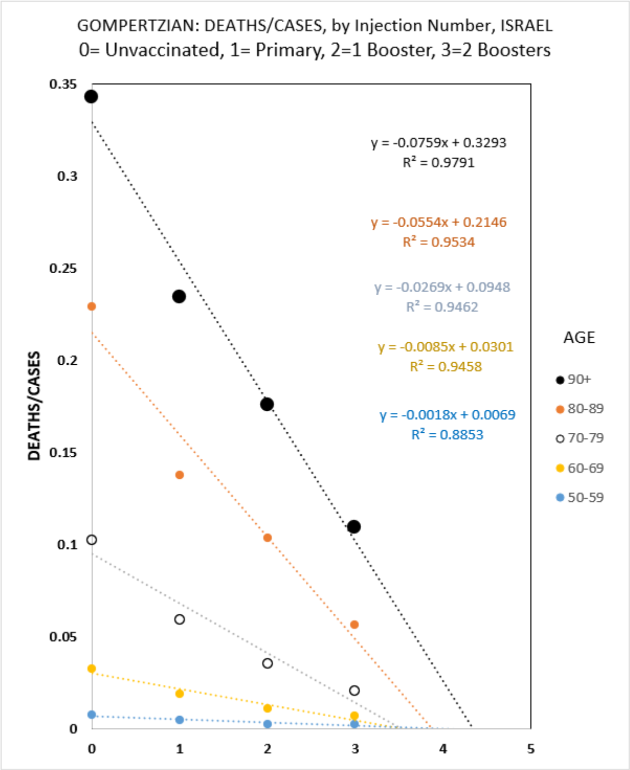


APPENDIX-FIGURE 36

APPENDIX-FIGURE 37

A second study from Israel data captured the number of COVID-19 Deaths, per populations, in the Pre-Vaccination era (March 23 2020-28 March 2021),[25] while the third study examined COVID-19 deaths, per population, among people over age 60 who had had a primary vaccination with one booster (3 doses) or two boosters (4 doses) (APPENDIX-FIGURE 38).[26] Combing these on ***Gompertz Plots*** of ***Malthusian Lethality***, reveals that an unvaccinated 80-year-old at the beginning on the epidemic would have had a 0.34% chance of death, declining to 0.16% after vaccination and 1 booster, and declining yet further to 0.014% after vaccination and two boosters, a twelve-fold reduction in the chance of death (APPENDIX-FIGURE 38)

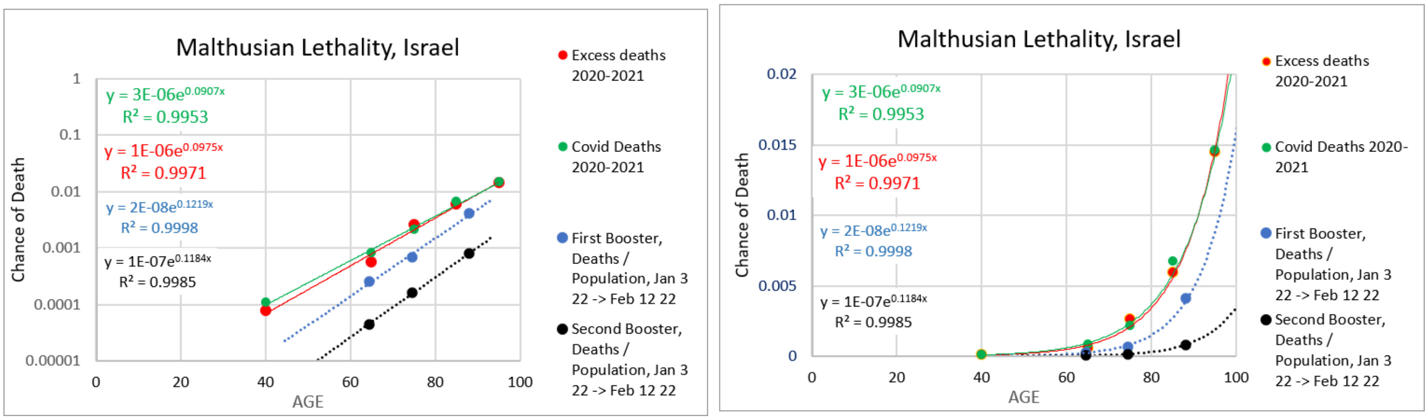


APPENDIX-FIGURE 38

Yet another study from Israel, provided data which revealed lower death rates for the those with 2 boosters (4 doses), than those with only 1 Booster (3 doses).[27] There were 35 deaths in among the 9,021 cases among those who had had 3 doses (1st booster) (0.38%), and 9 deaths among the 5,040 cases among those who had 4 doses (2 boosters) (0.18%). These deaths occurred among three age groups, and using the risk of death values that would have been expected for each age group, as calculated from the USA CDC data, for patients with a 4th dose (2 boosters), yielded an expect 0.09% risk of death, resulting in an expected of 5 deaths, which is in the same ballpark as the 9 deaths that actually occurred in this study group. Furthermore, patients who had had a previous COVID infections were excluded from analysis, so this study reveals the impact of sequential vaccination without the confounding immunizing effect of infection.

Finally, yet another study from Israel collected data on severe COVID-19. The data from this study also revealed a better outcome for those with 4 doses (2 boosters) than those who had only 3 doses (1 booster) (APPENDIX-FIGURE 39).[28]

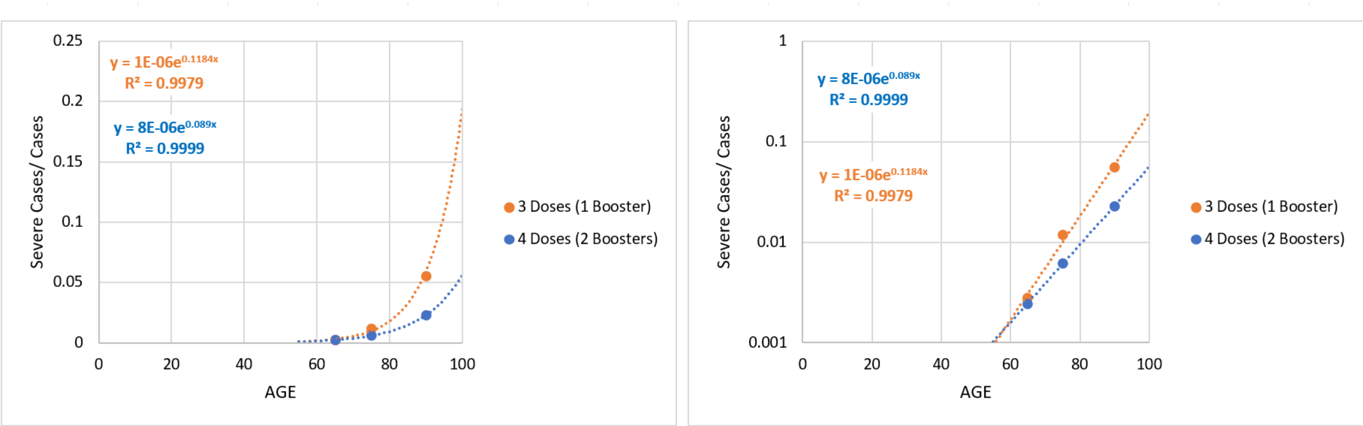


APPENDIX-FIGURE 39

IV. <u>More recent 2nd Booster Studies</u>

A number of more recent studies have examined the relative rate of COVID death in those with 4 doses (2 boosters) vs those who had had 3 doses (1 booster), and while these studies have not provided fine-grained enough datapoints for ***Gompertzian Analysis***, all of these studies have shown the superior death-reducing potential of the additional dose. Two studies were of COVID-19 deaths among patients in in old age homes,[29,30] another of death in a long-term scare facility, [31] another of rates of admission to intensive care or death among elderly (isolated death values not provided)[32], and another of infection, hospitalization, or death in nursing home residents[33] All of these studies have found a superior reduction in death for 2 boosters (4 doses) over 1 booster (3 doses).

WHO'S DYING NOW?

The data reviewed above make clear that people who have had 2 boosters have a better chance of surviving after infection than people with 1 booster, while people with 1 booster have a better chance than people with just primary vaccination, and people with primary vaccination have a better chance than the unvaccinated. The data also show that the vaccinated have a lower chance of getting infected than the unvaccinated.

So how does all this translate into who is dying now? The CDC vaccination and booster status data on people with COVID-19 from March 2022 through July 2022[20] give us a picture of this matter (APPENDIX-TABLE V). 38% of the deaths occurred among the unvaccinated, even though they made up only 12% of the populations. Only 5% of the deaths occurred among those who have had two boosters, even though they made up 17% of the populations.

Those with only one booster, and those with only primary vaccination, lowered chance of dying of COVID-19, by ~1/4$^{th}$, when compared with those without vaccination. Those with two boosters had only ~1/10$^{th}$ of the chance of dying of COVID-19, in comparison to those without vaccination.

| | deaths | % of all deaths | cases | population 7/22/22 | % | CASES/ Population (7/22) | DEATHS/ Population (7/22) | deaths/ million/ week | % |
|---|---|---|---|---|---|---|---|---|---|
| unvaccinated | 6,559 | 38% | 707,447 | 8,902,210.65 | 12% | 7.9% | 0.074% | 41 | 63% |
| vaccinated | 3,906 | 23% | 511,859 | 22,898,297 | 31% | 2.2% | 0.017% | 9 | 15% |
| one booster | 5,755 | 34% | 1,129,984 | 29,289,472 | 40% | 3.9% | 0.012% | 11 | 17% |
| two boosters | 892 | 5% | 244,979 | 12,667,677 | 17% | 1.9% | 0.007% | 4 | 6% |
| Total | 17,112 | 100% | 2,594,269 | 73,757,657 | 100% | 3.5% | 0.117% | 65 | 100% |

APPENDIX-TABLE V[20]

There were 41 deaths per million Americans per week among the unvaccinated, but only 4 deaths per million Americans per week among those who had had 2 boosters. Of course, the additional boosters now being offered rase the possibility of further savings in life, as suggested by the progressive linear reductions in ***Gompertzian Lethality*** (Deaths/Cases) that can be seen in the data (APPENDIX-FIGURE 29) (APPENDIX-FIGURE 32). The fact that this latest booster also contains antigens for ***Omicron*** offers the potential for additional reduction in ***Pasteurian Infectivity*** (Cases/Population). Clearly, we shall be watching the data to see whether these hopeful possibilities are realized. However, the lesson is clear: It's the unvaccinated who are dying now, as are those who have not had all of the available boosters. For those who are most fully boosted, COVID-19 death is becoming a memory.

Unfortunately, it is not such an encouraging story for COVID-19 infection. As can be seen in APPENDIX-FIGURE 27, and others throughout this APPENDIX, for those who are fully boosted, the chance of death, ***Gompertzian Lethality***, may be drifting into the past, but the chance of becoming infected, ***Pasteurian Infectivity***, is not.

***Gompertzian Analysis*** of LONG COVID

Long COVID manifests in multiple symptoms, which appear and last after infection,[34,35] as does pandemic associated excess mortality.[36,37,38,39] Unlike ***Gompertzian Lethality***, long COVID appears to occur at relatively equivalent rates across ages, as reported in studies from Turkey[40] and UK[41] (APPENDIX-FIGURE 40), and elsewhere. Whether this age-equivalence occurs across each of the many Long-COVID symptoms is not clear and would be most worthy of a closer look.

Fortunately, vaccination has been found to protect against Long COVID,[42,4344,45], including several studies that have found progressive greater protected as the numbers of doses are accumulated, as can be seen in one study whose values are shown APPENDIX-FIGURE 41 [46] A large study of Long COVID in Scotland found vaccination reduced some Long COVID cases associated with 7 symptoms, although all were in fewer than 5% of Long COVID patients, and vaccine reduction was not reported of the most common symptoms: tiredness, headache, muscle aches/weakness, joint pain, breathless.[47] Al-Aly, Bowe, and Xie,[48] using USA VA data, found that vaccination (dose numbers not noted) reduced the risk of Long COVID, including increased risk death, 30 days after infection, but only partially. As they stated: "the findings suggest that vaccination before infection confers only partial protection in the post-acute phase of the disease; hence, reliance on it as a sole mitigation strategy may not optimally reduce long-term health consequences of SARS-CoV-2 infection." Clearly, whether additional boosters continue to reduce Long COVID, and precisely which symptoms/illnesses are reduced by vaccination, are topics most worthy of continued analysis.

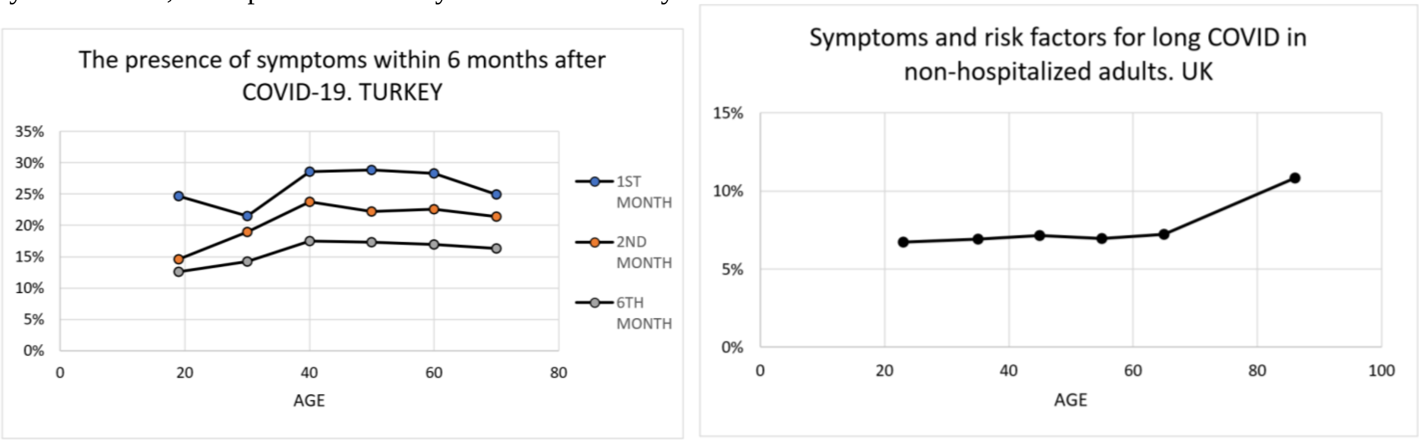


APPENDIX-FIGURE 40

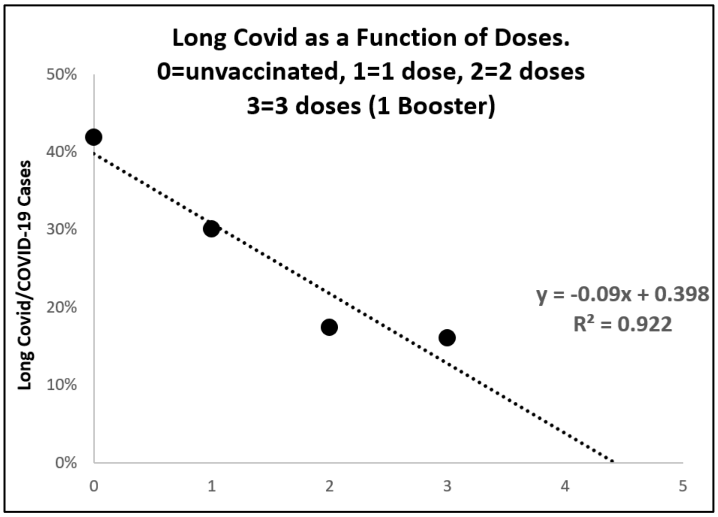


APPENDIX-FIGURE 41

<u>***Gompertzian Analysis*** of the Impact of Non-Immunological Factors On COVID-19 Outcome</u>

Many comorbidity factors have been associated with mortality generally, and COVID-19 lethality specifically. ***Gompertzian Analysis*** gives us some insights, and a single mathematical framework, for assessing, and integrating, the impact of such factors.

<u>The impact of Sex on Mortality</u>

Women live longer than men. Census Bureau data, displayed on the ***Gompertz Plot*** shown in APPENDIX-FIGURE 42 reveals that this sex-associated survival is roughly captured by a difference in ***Gompertzian Height***, $\boldsymbol{G_H}$, with a somewhat similar ***Gompertzian Slope***, $\boldsymbol{G_S}$. That is, at every age, men have a greater chance of death than women. This gives the appearance of roughly parallel ***Gompertz Lines***. (The subtle departure and curviness from absolute linearity and parallelism that the observant reader will note in APPENDIX-FIGURE 42 is a topic of continued analysis by my colleagues and I, but, fortunately, this quality does not affect the rough generalizations we shall discuss here.)

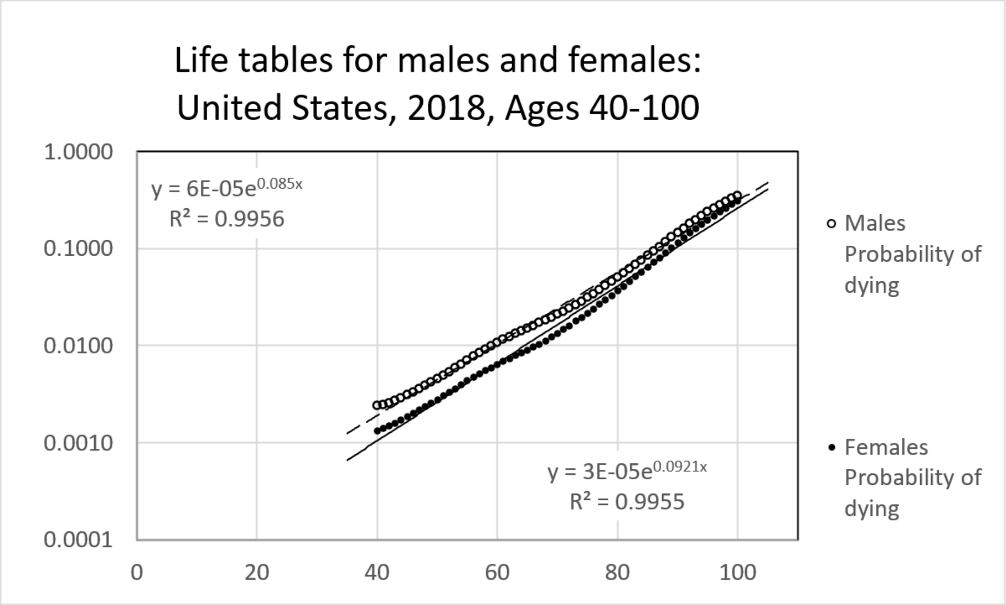


APPENDIX-FIGURE 42

Women also have a lower chance of dying of COVID-19 than men. A study from patients in Mexico provided age-sorted data on cases and death, with which it was possible to carry out ***Gompertzian Analysis*** of the impact of sex on COVID 19 outcome (APPENDIX-FIGURE 43)[49]. These data revealed lower ***Gompertzian Lethality*** for women in comparison to men. The difference is captured, yet again, by a difference in ***Gompertzian Height***, $\boldsymbol{G_H}$, with both sexes expressing the somewhat similar ***Gompertzian Slope***, $\boldsymbol{G_S}$, as we have seen in some many instances above. That is, ***Gompertzian Analysis*** reveals that sex-associated differences in COVID-19 ***Gompertzian Lethality*** (Deaths/Cases) occurs similarly across all ages. There was no evident male/female difference in ***Pasteurian Infectivity*** (Cases/Population).

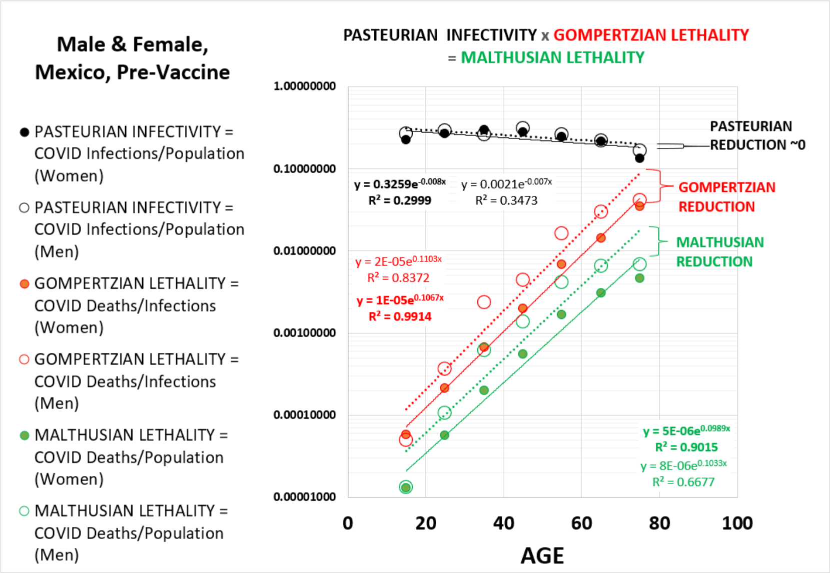


APPENDIX-FIGURE 43

The impact of Diabetes on COVID-19 Infection and Mortality

Diabetes has also been known to increase the lethal burden of COVID-10. A study from patients in Sweden provided age-sorted data on cases and death, with which it was possible to carry out ***Gompertzian Analysis*** of the impact of diabetes on COVID-19 outcome (APPENDIX-FIGURE 44).[50] These data revealed higher ***Gompertzian Lethality*** for diabetics in comparison to controls. The difference is captured, yet again, by a difference in ***Gompertzian Height***, $G_H$, with both diabetics and controls expressing the somewhat similar ***Gompertzian Slope***, $G_S$, as we have seen in some many instances above. There was also a strikingly evident diabetic/control difference in ***Pasteurian Infectivity*** (Cases/Population), with the two lines on the ***Gompertzian Plot*** being of a similar (but not identical) slope. That is, ***Gompertzian Analysis*** reveals that diabetes-associated differences in COVID-19 ***Gompertzian Lethality*** (Deaths/Cases) and ***Pasteurian Infectivity*** (Cases/Population), both occurring across all ages, leading to a disturbing negative ***Malthusian Reduction*** in ***Malthusian Lethality*** (Deaths//Population).

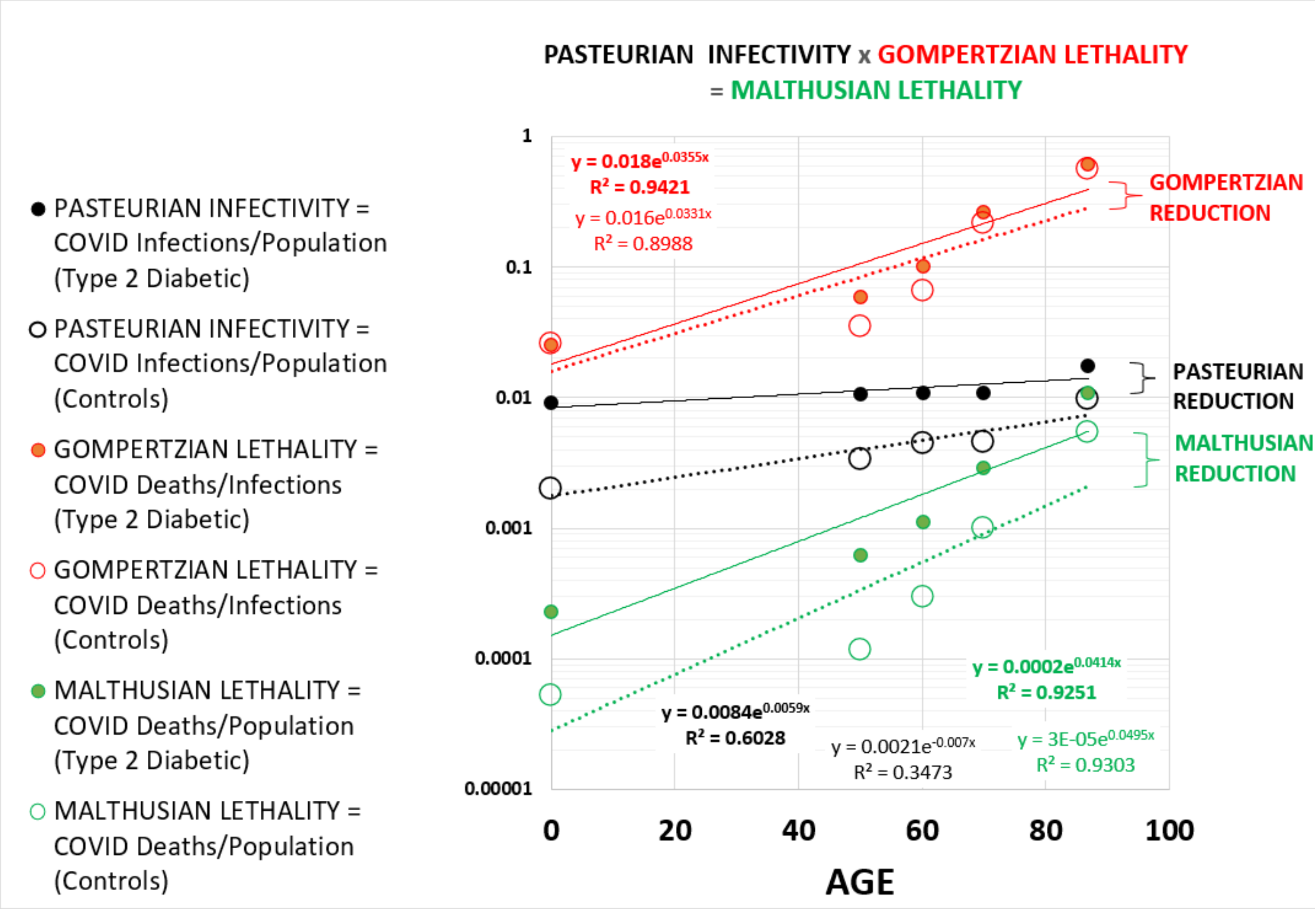

APPENDIX-FIGURE 44

The impact of Variants on COVID-19 Infection and Mortality.

Two studies provided age-sorted data on cases and death, one in the UK,[51] and the other in the USA[52], with which it was possible to carry out ***Gompertzian Analysis*** of COVID-19 variants. In both cases, ***Delta*** was associated with lower ***Gompertzian Lethality*** than the initial variant, and ***Omicron*** with and even lower level of ***Gompertzian Lethality*** (APPENDIX-FIGURES 45 AND 46). These differences are captured, yet again, by differences in ***Gompertzian Height***, ***$G_H$***, with each expressing the somewhat similar ***Gompertzian Slope***, ***$G_S$***, as we have seen in some many instances above. That is, ***Gompertzian Analysis*** reveals that the same variant-associated differences in COVID-19 ***Gompertzian Lethality*** (Deaths/Cases) occurs similarly across all ages.

Of course, the measures of variant-associated differences in COVID-19 ***Gompertzian Lethality*** (Deaths/Cases) come from separate points in time, when different levels vaccination, infection-acquire immunity, and other forces are likely to have been at work, in addition to the variant's properties themselves. Thus, the data shown here don't tell us which of these forces are at work, although this would certainly be a worthwhile exercise, and a perfectly achievable goal, should the underlying data be available. At the very least, however, we can see that the emergence of these new variants was not correlated with an increased chance of death after infection.

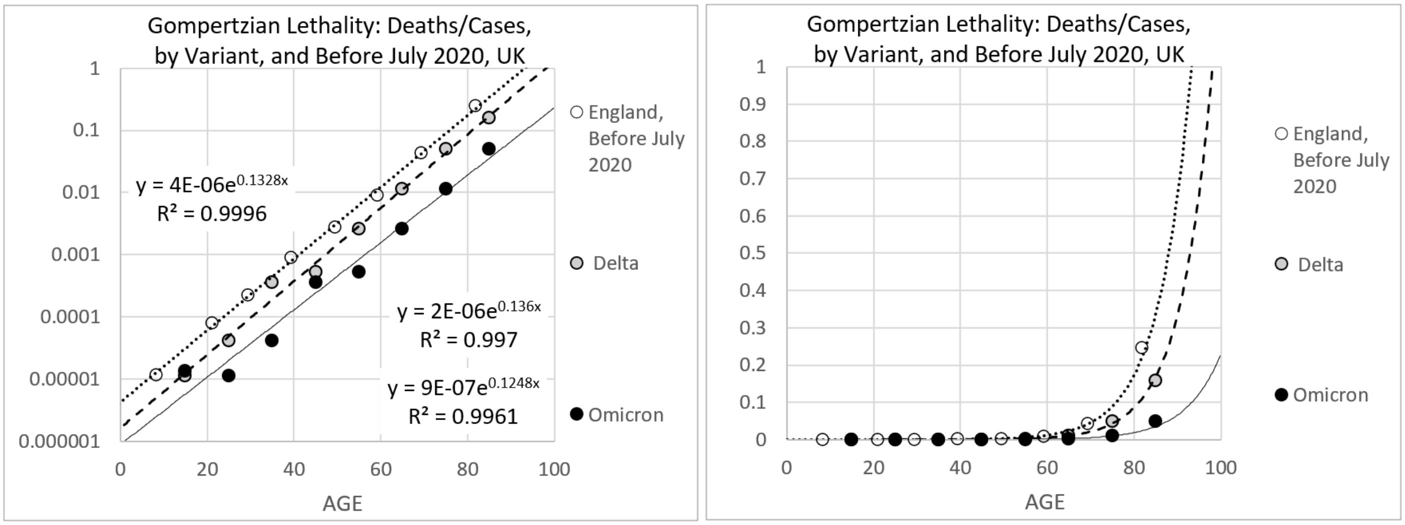


APPENDIX-FIGURE 45

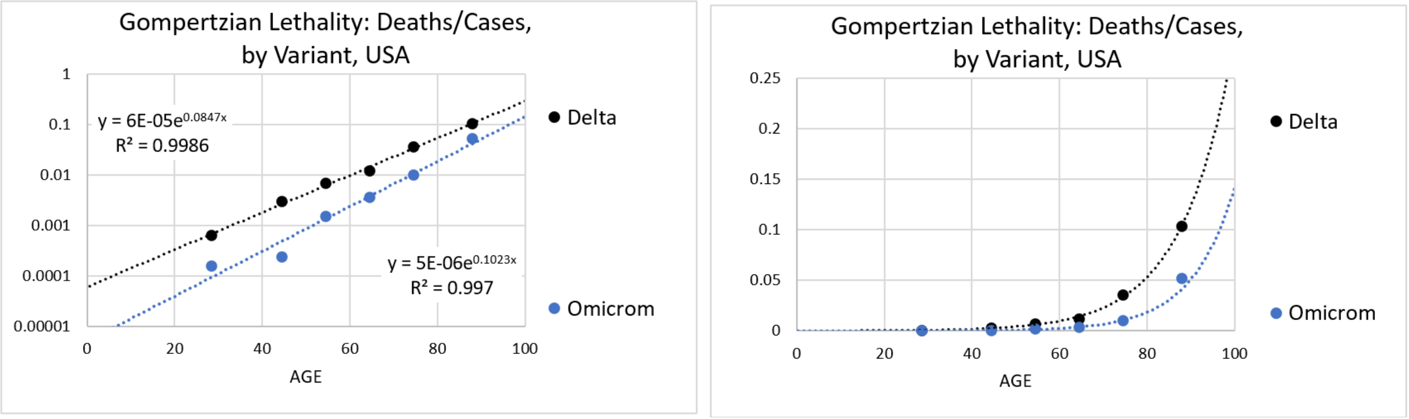


APPENDIX-FIGURE 46

REFERENCES

[1] Levin AT, Hanage WP, Owusu-Boaitey N, Cochran KB, Walsh SP, Meyerowitz-Katz G. Assessing the age specificity of infection fatality rates for COVID-19: systematic review, meta-analysis, and public policy implications. Eur J Epidemiol. 2020 Dec;35(12):1123-1138. doi: 10.1007/s10654-020-00698-1. Epub 2020 Dec 8. PMID: 33289900; PMCID: PMC7721859.

[2] Brazeau NF, Verity R, Jenks S, Fu H, Whittaker C, Winskill P, Dorigatti I, Walker PGT, Riley S, Schnekenberg RP, Hoeltgebaum H, Mellan TA, Mishra S, Unwin HJT, Watson OJ, Cucunubá ZM, Baguelin M, Whittles L, Bhatt S, Ghani AC, Ferguson NM, Okell LC. Estimating the COVID-19 infection fatality ratio accounting for seroreversion using statistical modelling. Commun Med (Lond). 2022 May 19;2:54. doi: 10.1038/s43856-022-00106-7. PMID: 35603270; PMCID: PMC9120146.

[3] O'Driscoll M, Ribeiro Dos Santos G, Wang L, Cummings DAT, Azman AS, Paireau J, Fontanet A, Cauchemez S, Salje H. Age-specific mortality and immunity patterns of SARS-CoV-2. Nature. 2021 Feb;590(7844):140-145. doi: 10.1038/s41586-020-2918-0. Epub 2020 Nov 2. PMID: 33137809.

[4] Verity R, Okell LC, Dorigatti I, Winskill P, Whittaker C, Imai N, Cuomo-Dannenburg G, Thompson H, Walker PGT, Fu H, Dighe A, Griffin JT, Baguelin M, Bhatia S, Boonyasiri A, Cori A, Cucunubá Z, FitzJohn R, Gaythorpe K, Green W, Hamlet A, Hinsley W, Laydon D, Nedjati-Gilani G, Riley S, van Elsland S, Volz E, Wang H, Wang Y, Xi X, Donnelly CA, Ghani AC, Ferguson NM. Estimates of the severity of coronavirus disease 2019: a model-based analysis. Lancet Infect Dis. 2020 Jun;20(6):669-677. doi: 10.1016/S1473-3099(20)30243-7. Epub 2020 Mar 30. Erratum in: Lancet Infect Dis. 2020 Apr 15;: Erratum in: Lancet Infect Dis. 2020 May 4;: PMID: 32240634; PMCID: PMC7158570.

[5] Wood SN, Wit EC, Fasiolo M, Green PJ. COVID-19 and the difficulty of inferring epidemiological parameters from clinical data. Lancet Infect Dis. 2021 Jan;21(1):27-28. doi: 10.1016/S1473-3099(20)30437-0. Epub 2020 May 28. PMID: 32473661; PMCID: PMC7255708.

[6] Levin AT, Owusu-Boaitey N, Pugh S, Fosdick BK, Zwi AB, Malani A, Soman S, Besançon L, Kashnitsky I, Ganesh S, McLaughlin A, Song G, Uhm R, Herrera-Esposito D, de Los Campos G, Peçanha Antonio ACP, Tadese EB, Meyerowitz-Katz G. Assessing the burden of COVID-19 in developing countries: systematic review, meta-analysis and public policy implications. BMJ Glob Health. 2022 May;7(5):e008477. doi: 10.1136/bmjgh-2022-008477. PMID: 35618305; PMCID: PMC9136695.

[7] COVID-19 Forecasting Team. Variation in the COVID-19 infection-fatality ratio by age, time, and geography during the pre-vaccine era: a systematic analysis. Lancet. 2022 Apr 16;399(10334):1469-1488. doi: 10.1016/S0140-6736(21)02867-1. Epub 2022 Feb 24. Erratum in: Lancet. 2022 Apr 16;399(10334):1468. PMID: 35219376; PMCID: PMC8871594.

[8] Glynn JR, Moss PAH. Systematic analysis of infectious disease outcomes by age shows lowest severity in school-age children. Sci Data. 2020 Oct 15;7(1):329. doi: 10.1038/s41597-020-00668-y. PMID: 33057040; PMCID: PMC7566589.

[9] Jones HB, A SPECIAL CONSIDERATION OF THE AGING PROCESS, DISEASE, AND LIFE EXPECTANCY November 1, 1955 CLEVELAND PUBLIC LIBRARY TECHNOLOGY DIVISION MAR 2 1956 SERIAL DOCS OARDS Printed for the U. S. Atomic Energy Commission UNIVERSITY OF CALIFORNIA Radiation Laboratory Berkeley, California Contract No. W - 7405 - eng – 48. https://www.google.com/books/edition/A_Special_Consideration_of_the_Aging_Pro/xLL4pko-4hoC?hl=en

[10] Citi L, Su, J, Huang, L, Michaelson, J. Counting Cells By Age Tells Us How, and Why, and When, We Grow, and Become Old and Ill. 2022. In Preparation

[11] Tartof SY, Qian L, Hong V, Wei R, Nadjafi RF, Fischer H, Li Z, Shaw SF, Caparosa SL, Nau CL, Saxena T, Rieg GK, Ackerson BK, Sharp AL, Skarbinski J, Naik TK, Murali SB. Obesity and Mortality Among Patients Diagnosed With COVID-19: Results From an Integrated Health Care Organization. Ann Intern Med. 2020 Nov 17;173(10):773-781. doi: 10.7326/M20-3742. Epub 2020 Aug 12. PMID: 32783686; PMCID: PMC7429998.

[12] CDC COVID Data Tracker: Rates of COVID-19 Cases and Deaths by Vaccination Status
https://covid.cdc.gov/covid-data-tracker/#rates-by-vaccine-status
https://data.cdc.gov/Public-Health-Surveillance/Rates-of-COVID-19-Cases-or-Deaths-by-Age-Group-and/ukww-au2k

[13] Ioannou GN, Locke ER, O'Hare AM, Bohnert ASB, Boyko EJ, Hynes DM, Berry K. COVID-19 Vaccination Effectiveness Against Infection or Death in a National U.S. Health Care System : A Target Trial Emulation Study. Ann Intern Med. 2022 Mar;175(3):352-361. doi: 10.7326/M21-3256. Epub 2021 Dec 21. PMID: 34928700; PMCID: PMC8697485.

[14] Ioannou GN, Locke ER, Green PK, Berry K. Comparison of Moderna versus Pfizer-BioNTech COVID-19 vaccine outcomes: A target trial emulation study in the U.S. Veterans Affairs healthcare system. EClinicalMedicine. 2022 Mar 5;45:101326. doi: 10.1016/j.eclinm.2022.101326. PMID: 35261970; PMCID: PMC8896984.

[15] Vokó Z, Kiss Z, Surján G, Surján O, Barcza Z, Pályi B, Formanek-Balku E, Molnár GA, Herczeg R, Gyenesei A, Miseta A, Kollár L, Wittmann I, Müller C, Kásler M. Nationwide effectiveness of five SARS-CoV-2 vaccines in Hungary-the HUN-VE study. Clin Microbiol Infect. 2022 Mar;28(3):398-404. doi: 10.1016/j.cmi.2021.11.011. Epub 2021 Nov 25. PMID: 34838783; PMCID: PMC8612758.

[16] Kiss Z, Wittmann I, Polivka L, Surján G, Surján O, Barcza Z, Molnár GA, Nagy D, Müller V, Bogos K, Nagy P, Kenessey I, Wéber A, Pálosi M, Szlávik J, Schaff Z, Szekanecz Z, Müller C, Kásler M, Vokó Z. Nationwide Effectiveness of First and Second SARS-CoV2 Booster Vaccines During the Delta and Omicron Pandemic Waves in Hungary (HUN-VE 2 Study). Front Immunol. 2022 Jun 23;13:905585. doi: 10.3389/fimmu.2022.905585. PMID: 35812442; PMCID: PMC9260843.

[17] Kiss Z, Wittmann I, Polivka L, Surján G, Surján O, Barcza Z, Molnár GA, Nagy D, Müller V, Bogos K, Nagy P, Kenessey I, Wéber A, Pálosi M, Szlávik J, Schaff Z, Szekanecz Z, Müller C, Kásler M, Vokó Z. Nationwide Effectiveness of First and Second SARS-CoV2 Booster Vaccines During the Delta and Omicron Pandemic Waves in Hungary (HUN-VE 2 Study). Front Immunol. 2022 Jun 23;13:905585. doi: 10.3389/fimmu.2022.905585. PMID: 35812442; PMCID: PMC9260843.

[18] Rearte A, Castelli JM, Rearte R, Fuentes N, Pennini V, Pesce M, Barbeira PB, Iummato LE, Laurora M, Bartolomeu ML, Galligani G, Del Valle Juarez M, Giovacchini CM, Santoro A, Esperatti M, Tarragona S, Vizzotti C. Effectiveness of rAd26-rAd5, ChAdOx1 nCoV-19, and BBIBP-CorV vaccines for risk of infection with SARS-CoV-2 and death due to COVID-19 in people older than 60 years in Argentina: a test-negative, case-control, and retrospective longitudinal study. Lancet. 2022 Mar 26;399(10331):1254-1264. doi: 10.1016/S0140-6736(22)00011-3. Epub 2022 Mar 15. Erratum in: Lancet. 2022 Jun 11;399(10342):2190. PMID: 35303473; PMCID: PMC8923678.

[19] Goldberg Y, Mandel M, Woodbridge Y, Fluss R, Novikov I, Yaari R, Ziv A, Freedman L, Huppert A. Similarity of Protection Conferred by Previous SARS-CoV-2 Infection and by BNT162b2 Vaccine: A 3-Month Nationwide Experience From Israel. Am J Epidemiol. 2022 Jul 23;191(8):1420-1428. doi: 10.1093/aje/kwac060. PMID: 35355048; PMCID: PMC8992290.

[20] CDC COVID Data Tracker: Rates of COVID-19 Cases and Deaths by Vaccination Status
https://covid.cdc.gov/covid-data-tracker/#rates-by-vaccine-status
https://data.cdc.gov/Public-Health-Surveillance/Rates-of-COVID-19-Cases-or-Deaths-by-Age-Group-and/ukww-au2k

[21] Eythorsson E, Runolfsdottir HL, Ingvarsson RF, Sigurdsson MI, Palsson R. Rate of SARS-CoV-2 Reinfection During an Omicron Wave in Iceland. JAMA Netw Open. 2022 Aug 1;5(8):e2225320. doi: 10.1001/jamanetworkopen.2022.25320. PMID: 35921113; PMCID: PMC9350711.

[22] Kiss Z, Wittmann I, Polivka L, Surján G, Surján O, Barcza Z, Molnár GA, Nagy D, Müller V, Bogos K, Nagy P, Kenessey I, Wéber A, Pálosi M, Szlávik J, Schaff Z, Szekanecz Z, Müller C, Kásler M, Vokó Z. Nationwide Effectiveness of First and Second SARS-CoV2 Booster Vaccines During the Delta and Omicron Pandemic Waves in Hungary (HUN-VE 2 Study). Front Immunol. 2022 Jun 23;13:905585. doi: 10.3389/fimmu.2022.905585. PMID: 35812442; PMCID: PMC9260843.

[23] Vokó Z, Kiss Z, Surján G, Surján O, Barcza Z, Wittmann I, Molnár GA, Nagy D, Müller V, Bogos K, Nagy P, Kenessey I, Wéber A, Polivka L, Pálosi M, Szlávik J, Rokszin G, Müller C, Szekanecz Z, Kásler M. Effectiveness and Waning of Protection With Different SARS-CoV-2 Primary and Booster Vaccines During the Delta Pandemic Wave in 2021 in Hungary (HUN-VE 3 Study). Front Immunol. 2022 Jul 22;13:919408. doi: 10.3389/fimmu.2022.919408. PMID: 35935993; PMCID: PMC9353007.

[24] Saban M, Myers V, Wilf-Miron R. Changes in infectivity, severity and vaccine effectiveness against delta COVID-19 variant ten months into the vaccination program: The Israeli case. Prev Med. 2022 Jan;154:106890. doi: 10.1016/j.ypmed.2021.106890. Epub 2021 Nov 17. PMID: 34800471; PMCID: PMC8596646..

[25] Chava Peretz 1, Naama Rotem 2, Lital Keinan-Boker 3 4, Avner Furshpan 5, Manfred Green 3, Michal Bitan 6, David M Steinberg 6
Excess mortality in Israel associated with COVID-19 in 2020-2021 by age group and with estimates based on daily mortality patterns in 2000-2019 Int J Epidemiol. 2022 Jun 13;51(3):727-736. doi: 10.1093/ije/dyac047.

[26] Ronen Arbel, Ruslan Sergienko, Michael Friger, Alon Peretz, Tanya Beckenstein, Shlomit Yaron, Doron Netzer, Ariel Hammerman
Effectiveness of a second BNT162b2 booster vaccine against hospitalization and death from COVID-19 in adults aged over 60 years
Nat Med . 2022 Jul;28(7):1486-1490. doi: 10.1038/s41591-022-01832-0. Epub 2022 Apr 25.

[27] Magen O, Waxman JG, Makov-Assif M, et al. Fourth dose of BNT162b2 mRNA Covid-19 vaccine in a nationwide setting. N Engl J Med 2022; 386: 1603-14 Epub 2022 Apr 13.

[28] Yinon M Bar-On, Yair Goldberg, Micha Mandel, Omri Bodenheimer, Ofra Amir, Laurence Freedman, Sharon Alroy-Preis, Nachman Ash, Amit Huppert, Ron Milo Protection by a Fourth Dose of BNT162b2 against Omicron in Israel N Engl J Med . 2022 May 5;386(18):1712-1720. doi: 10.1056/NEJMoa2201570. Epub 2022 Apr 5.

[29] Muhsen K, Maimon N, Mizrahi AY, Boltyansky B, Bodenheimer O, Diamant ZH, Gaon L, Cohen D, Dagan R. Association of Receipt of the Fourth BNT162b2 Dose With Omicron Infection and COVID-19 Hospitalizations Among Residents of Long-term Care Facilities. JAMA Intern Med. 2022 Aug 1;182(8):859-867. doi: 10.1001/jamainternmed.2022.2658. PMID: 35737368; PMCID: PMC9227688

[30] Nordström P, Ballin M, Nordström A. Effectiveness of a fourth dose of mRNA COVID-19 vaccine against all-cause mortality in long-term care facility residents and in the oldest old: A nationwide, retrospective cohort study in Sweden. Lancet Reg Health Eur. 2022 Oct;21:100466. doi: 10.1016/j.lanepe.2022.100466. Epub 2022 Jul 13. PMID: 35855494; PMCID: PMC9277096.

[31] Grewal R, Kitchen SA, Nguyen L, Buchan SA, Wilson SE, Costa AP, Kwong JC. Effectiveness of a fourth dose of covid-19 mRNA vaccine against the omicron variant among long term care residents in Ontario, Canada: test negative design study. BMJ. 2022 Jul 6;378:e071502. doi: 10.1136/bmj-2022-071502. PMID: 35793826; PMCID: PMC9257064.

[32] Tan CY, Chiew CJ, Lee VJ, Ong B, Lye DC, Tan KB. Effectiveness of a Fourth Dose of COVID-19 mRNA Vaccine Against Omicron Variant Among Elderly People in Singapore. Ann Intern Med. 2022 Nov;175(11):1622-1623. doi: 10.7326/M22-2042. Epub 2022 Sep 13. PMID: 36095316; PMCID: PMC9578545.

[33] McConeghy KW, White EM, Blackman C, Santostefano CM, Lee Y, Rudolph JL, Canaday D, Zullo AR, Jernigan JA, Pilishvili T, Mor V, Gravenstein S. Effectiveness of a Second COVID-19 Vaccine Booster Dose Against Infection, Hospitalization, or Death Among Nursing Home Residents - 19 States, March 29-July 25, 2022. MMWR Morb Mortal Wkly Rep. 2022 Sep 30;71(39):1235-1238. doi: 10.15585/mmwr.mm7139a2. PMID: 36173757; PMCID: PMC9533729.

[34] Global Burden of Disease Long COVID Collaborators, Wulf Hanson S, Abbafati C, Aerts JG, Al-Aly Z, Ashbaugh C, Ballouz T, Blyuss O, Bobkova P, Bonsel G, Borzakova S, Buonsenso D, Butnaru D, Carter A, Chu H, De Rose C, Diab MM, Ekbom E, El Tantawi M, Fomin V, Frithiof R, Gamirova A, Glybochko PV, Haagsma JA, Haghjooy Javanmard S, Hamilton EB, Harris G, Heijenbrok-Kal MH, Helbok R, Hellemons ME, Hillus D, Huijts SM, Hultström M, Jassat W, Kurth F, Larsson IM, Lipcsey M, Liu C, Loflin CD, Malinovschi A, Mao W, Mazankova L, McCulloch D, Menges D, Mohammadifard N, Munblit D, Nekliudov NA, Ogbuoji O, Osmanov IM, Peñalvo JL, Petersen MS, Puhan MA, Rahman M, Rass V, Reinig N, Ribbers GM, Ricchiuto A, Rubertsson S, Samitova E, Sarrafzadegan N, Shikhaleva A, Simpson KE, Sinatti D, Soriano JB, Spiridonova E, Steinbeis F, Svistunov AA, Valentini P, van de Water BJ, van den Berg-Emons R, Wallin E, Witzenrath M, Wu Y, Xu H, Zoller T, Adolph C, Albright J, Amlag JO, Aravkin AY, Bang-Jensen BL, Bisignano C, Castellano R, Castro E, Chakrabarti S, Collins JK, Dai X, Daoud F, Dapper C, Deen A, Duncan BB, Erickson M, Ewald SB, Ferrari AJ, Flaxman AD, Fullman N, Gamkrelidze A, Giles JR, Guo G, Hay SI, He J, Helak M, Hulland EN, Kereselidze M, Krohn KJ, Lazzar-Atwood A, Lindstrom A, Lozano R, Malta DC, Månsson J, Mantilla Herrera AM, Mokdad AH, Monasta L, Nomura S, Pasovic M, Pigott DM, Reiner RC Jr, Reinke G, Ribeiro ALP, Santomauro DF, Sholokhov A, Spurlock EE, Walcott R, Walker A, Wiysonge CS, Zheng P, Bettger JP, Murray CJL, Vos T. Estimated Global Proportions of Individuals With Persistent Fatigue, Cognitive, and Respiratory Symptom Clusters Following Symptomatic COVID-19 in 2020 and 2021. JAMA. 2022 Oct 25;328(16):1604-1615. doi: 10.1001/jama.2022.18931. PMID: 36215063; PMCID: PMC9552043..

[35] Al-Aly Z, Bowe B, Xie Y. Long COVID after breakthrough SARS-CoV-2 infection. Nat Med. 2022 Jul;28(7):1461-1467. doi: 10.1038/s41591-022-01840-0. Epub 2022 May 25. PMID: 35614233; PMCID: PMC9307472..

[36] https://data.gov.scot/coronavirus-covid-19/detail.html#excess_deaths

[37] Paglino E, Lundberg DJ, Cho A, Wasserman JA, Raquib R, Luck AN, Hempstead K, Bor J, Elo IT, Preston SH, Stokes AC. Excess all-cause mortality across counties in the United States, March 2020 to December 2021. medRxiv [Preprint]. 2022 May 17:2022.04.23.22274192. doi: 10.1101/2022.04.23.22274192. PMID: 35547848; PMCID: PMC9094106.
https://www.ncbi.nlm.nih.gov/pmc/articles/PMC9094106/

[38] Excess Deaths Associated with COVID-19 https://www.cdc.gov/nchs/nvss/vsrr/covid19/excess_deaths.htm

[39] Excess mortality in England analysis
https://app.powerbi.com/view?r=eyJrIjoiYmUwNmFhMjYtNGZhYS00NDk2LWFlMTAtOTg0OGNhNmFiNGM0IiwidCI6ImVlNGUxNDk5LTRhMzUtNGIyZS1hZDQ3LTVmM2NmOWRlODY2NiIsImMiOjh9

[40] Emecen AN, Keskin S, Turunc O, Suner AF, Siyve N, Basoglu Sensoy E, Dinc F, Kilinc O, Avkan Oguz V, Bayrak S, Unal B. The presence of symptoms within 6 months after COVID-19: a single-center longitudinal study. Ir J Med Sci. 2022 Jun 17:1–10. doi: 10.1007/s11845-022-03072-0. Epub ahead of print. PMID: 35715663; PMCID: PMC9205653.

[41] Subramanian A, Nirantharakumar K, Hughes S, Myles P, Williams T, Gokhale KM, Taverner T, Chandan JS, Brown K, Simms-Williams N, Shah AD, Singh M, Kidy F, Okoth K, Hotham R, Bashir N, Cockburn N, Lee SI, Turner GM, Gkoutos GV, Aiyegbusi OL, McMullan C, Denniston AK, Sapey E, Lord JM, Wraith DC, Leggett E, Iles C, Marshall T, Price MJ, Marwaha S, Davies EH, Jackson LJ, Matthews KL, Camaradou J, Calvert M, Haroon S. Symptoms and risk factors for long COVID in non-hospitalized adults. Nat Med. 2022 Aug;28(8):1706-1714. doi: 10.1038/s41591-022-01909-w. Epub 2022 Jul 25. PMID: 35879616; PMCID: PMC9388369.

[42] Gao P, Liu J, Liu M. Effect of COVID-19 Vaccines on Reducing the Risk of Long COVID in the Real World: A Systematic Review and Meta-Analysis. Int J Environ Res Public Health. 2022 Sep 29;19(19):12422. doi: 10.3390/ijerph191912422. PMID: 36231717; PMCID: PMC9566528.

[43] Kuodi P, Gorelik Y, Zayyad H, Wertheim O, Wiegler KB, Abu Jabal K, Dror AA, Nazzal S, Glikman D, Edelstein M. Association between BNT162b2 vaccination and reported incidence of post-COVID-19 symptoms: cross-sectional study 2020-21, Israel. NPJ Vaccines. 2022 Aug 26;7(1):101. doi: 10.1038/s41541-022-00526-5. PMID: 36028498; PMCID: PMC9411827.

[44] Ayoubkhani D, Bosworth ML, King S, Pouwels KB, Glickman M, Nafilyan V, Zaccardi F, Khunti K, Alwan NA, Walker AS. Risk of Long COVID in People Infected With Severe Acute Respiratory Syndrome Coronavirus 2 After 2 Doses of a Coronavirus Disease 2019 Vaccine: Community-Based, Matched Cohort Study. Open Forum Infect Dis. 2022 Sep 12;9(9):ofac464. doi: 10.1093/ofid/ofac464. PMID: 36168555; PMCID: PMC9494414.

[45] Brannock MD, Chew RF, Preiss AJ, Hadley EC, McMurry JA, Leese PJ, Girvin AT, Crosskey M, Zhou AG, Moffitt RA, Funk MJ, Pfaff ER, Haendel MA, Chute CG; N3C and RECOVER Consortia. Long COVID Risk and Pre-COVID Vaccination: An EHR-Based Cohort Study from the RECOVER Program. medRxiv [Preprint]. 2022 Oct 7:2022.10.06.22280795. doi: 10.1101/2022.10.06.22280795. PMID: 36238713; PMCID: PMC9558440.

[46] Azzolini E, Levi R, Sarti R, Pozzi C, Mollura M, Mantovani A, Rescigno M. Association Between BNT162b2 Vaccination and Long COVID After Infections Not Requiring Hospitalization in Health Care Workers. JAMA. 2022 Aug 16;328(7):676-678. doi: 10.1001/jama.2022.11691. PMID: 35796131; PMCID: PMC9250078.

[47] Hastie CE, Lowe DJ, McAuley A, Winter AJ, Mills NL, Black C, Scott JT, O'Donnell CA, Blane DN, Browne S, Ibbotson TR, Pell JP. Outcomes among confirmed cases and a matched comparison group in the Long-COVID in Scotland study. Nat Commun. 2022 Oct 12;13(1):5663. doi: 10.1038/s41467-022-33415-5. Erratum in: Nat Commun. 2022 Nov 1;13(1):6540. PMID: 36224173; PMCID: PMC9556711.

[48] Al-Aly Z, Bowe B, Xie Y. Long COVID after breakthrough SARS-CoV-2 infection. Nat Med. 2022 Jul;28(7):1461-1467. doi: 10.1038/s41591-022-01840-0. Epub 2022 May 25. PMID: 35614233; PMCID: PMC9307472.

[49] Torres-Ibarra L, Basto-Abreu A, Carnalla M, Torres-Alvarez R, Reyes-Sanchez F, Hernández-Ávila JE, Palacio-Mejia LS, Alpuche-Aranda C, Shamah-Levy T, Rivera JA, Barrientos-Gutierrez T. SARS-CoV-2 infection fatality rate after the first epidemic wave in Mexico. Int J Epidemiol. 2022 May 9;51(2):429-439. doi: 10.1093/ije/dyac015. PMID: 35157072; PMCID: PMC8903396.

[50] Rawshani A, Kjölhede EA, Rawshani A, Sattar N, Eeg-Olofsson K, Adiels M, Ludvigsson J, Lindh M, Gisslén M, Hagberg E, Lappas G, Eliasson B, Rosengren A. Severe COVID-19 in people with type 1 and type 2 diabetes in Sweden: A nationwide retrospective cohort study. Lancet Reg Health Eur. 2021 May;4:100105. doi: 10.1016/j.lanepe.2021.100105. Epub 2021 Apr 30. PMID: 33969336; PMCID: PMC8086507.

[51] Nyberg T, Ferguson NM, Nash SG, Webster HH, Flaxman S, Andrews N, Hinsley W, Bernal JL, Kall M, Bhatt S, Blomquist P, Zaidi A, Volz E, Aziz NA, Harman K, Funk S, Abbott S; COVID-19 Genomics UK (COG-UK) consortium, Hope R, Charlett A, Chand M, Ghani AC, Seaman SR, Dabrera G, De Angelis D, Presanis AM, Thelwall S. Comparative analysis of the risks of hospitalisation and death associated with SARS-CoV-2 omicron (B.1.1.529) and delta (B.1.617.2) variants in England: a cohort study. Lancet. 2022 Apr 2;399(10332):1303-1312. doi: 10.1016/S0140-6736(22)00462-7. Epub 2022 Mar 16. PMID: 35305296; PMCID: PMC8926413.

[52] Skarbinski J, Wood MS, Chervo TC, Schapiro JM, Elkin EP, Valice E, Amsden LB, Hsiao C, Quesenberry C, Corley DA, Kushi LH. Risk of severe clinical outcomes among persons with SARS-CoV-2 infection with differing levels of vaccination during widespread Omicron (B.1.1.529) and Delta (B.1.617.2) variant circulation in Northern California: A retrospective cohort study. Lancet Reg Health Am. 2022 Aug;12:100297. doi: 10.1016/j.lana.2022.100297. Epub 2022 Jun 16. PMID: 35756977; PMCID: PMC9212563.
